# Projecting individualized probabilities of lifetime all-cancer risk

**DOI:** 10.1101/2025.07.23.25326097

**Authors:** Neel M. Butala, Noor Al-Hammadi, Asiri Ediriwickrema, Jaime L. Schneider, Aaron Fullerton, Jeya Balasubramanian, Parichoy Pal Choudhury, Nilanjan Chatterjee

## Abstract

**Introduction:** Technological advances and direct-to-consumer marketing have unearthed significant organic demand from patients for cancer screening and prevention. However, in the absence of strong data or guidelines, physicians have minimal support on how to approach patients in clinical practice.

**Methods:** We projected individualized probabilities of 10-year and lifetime cancer risk across a population as well as potential improvement with healthy behaviors in the UK Biobank.

**Results:** A total of 118 distinct variables were included across 38 cancer-specific models. The distribution of lifetime cancer risk had a rightward skew and wide variation for both men and women. The median lifetime cancer risk was 29.5% for men (interquartile range (IQR) 8.4%) and 21.0% for women (IQR 8.8%). If all modifiable risk factors were set to the ideal state, this decreased to 20.5% for men (IQR 3.9%) and 16.5% for women (IQR 4.9%). There was considerable overlap between age groups, with men aged 50-59 at the 90^th^ percentile having greater risk (11.9%) than men aged 60-70 at the 25^th^ percentile (11.8%), and women aged 40-49 at the 90^th^ percentile having greater risk (7.4%) than women aged 50-59 at the 60^th^ percentile (6.8%) and women aged 60-70 at the 20^th^ percentile (7.3%).

**Conclusions:** Lifetime cancer risk varies widely across the UK Biobank cohort, but this risk decreases substantially with healthy behaviors. There was considerable overlap in 10-year cancer risk between age groups, suggesting that future multicancer screening guidelines should account for more than age and sex as more evidence becomes available in the future.

## Introduction

Recent technological advances have unearthed significant organic demand from patients for novel cancer screening and prevention strategies.^1–3^ Multi-cancer early detection (MCED) tests and whole-body MRI screening have become commercially available and marketed directly to consumers.^4,5^ However, in the absence of strong data or guidelines, physicians have minimal support on how to counsel patients in clinical practice.^6^

Cancer risk varies substantially among individuals due to numerous cancer-specific risk factors. Multi-cancer screening strategies based solely on age or sex may overlook high-risk individuals while promoting low-value testing in low-risk individuals. Moreover, given that certain risk factors, such as smoking and obesity, influence multiple cancer types, preventive counseling can be more effective if done based on all-cancer risk. We created a model to project individual lifetime all-cancer risk and applied it to a population to evaluate (1) the distribution of risk by age and sex and (2) the potential impact of healthy behaviors.

## Methods

### Population

We included all participants in the UK Biobank (UKB) without a history of cancer at time of enrollment. UKB recruited approximately 500,000 individuals aged 40-70 across the UK between 2006-2010. ^7^ UKB has ethical approval from the North West Multi-centre Research Ethics Committee.

### Data Sources and Study Variables

For model development, we conducted a comprehensive literature review of meta-analyses, large-scale cohort studies, and existing cancer-specific models to identify established risk factors contributing to the risk of developing 21 common cancers. We used billing codes to ascertain diagnosis of these cancers for UKB participants.

For calibration and projection of lifetime risk, we obtained age-specific cancer incidence for 21 cancer types from the Cancer Research UK database and mortality risk from the UK Office for National Statistics.^8,9^

### Statistical analysis

We developed cancer-specific models separately for men and women in the UKB cohort to simultaneously select and quantify the impact of risk factors identified in the literature to account for collinearity (**Supplemental Table 1**). Additional variables strongly associated with cancer but not found in the UKB were appended to these models with risk associations based on literature review. For appended variables, participants were randomly assigned values simulated based on population-level prevalence data, as has previously been done.^10^ Full modeling details are described in the **Supplemental Methods**.

Individualized absolute probabilities of lifetime all-cancer risk in the UKB were projected using the iCARE package.^11,12^ Cancer models were calibrated to the broader UK population using UK cancer incidence files while accounting for competing mortality risk.

For each participant, we estimated individual cancer risk until age 85 (lifetime risk) based on age at recruitment. For 29 modifiable risk factors (e.g. diet, lifestyle, physical activity, oral health, etc.), we then substituted the ideal value for participants and recomputed lifetime risk to project the distribution of lifetime cancer risk if the UKB population had ideal values for risk factors that could still be modified (**Supplemental Table 2**). We calculated all-cancer risk for each participant by taking the inverse of the risk of not getting any individual cancer.

We additionally estimated 10-year individual all-cancer risk based on age at recruitment. We stratified this by decade of age at recruitment to project the distributions of 10-year all-cancer risk in the UKB by decade.

All analyses were stratified by sex and conducted using SAS v9.4, R v4.3.2, and Python v3.11.11.

## Results

The final study sample included 446,795 patients (**Supplemental Figure 1**). A total of 118 distinct variables were included across 38 cancer-specific models (**Supplemental Table 1**). The distribution of lifetime cancer risk had a rightward skew and wide variation for both men and women (**Figure 1, Supplemental Table 3**). The median lifetime cancer risk was 29.47% for men (interquartile range (IQR) 8.39%), whereas the median lifetime cancer risk was 21.00% for women (IQR 8.78%).

**Figure 1.**
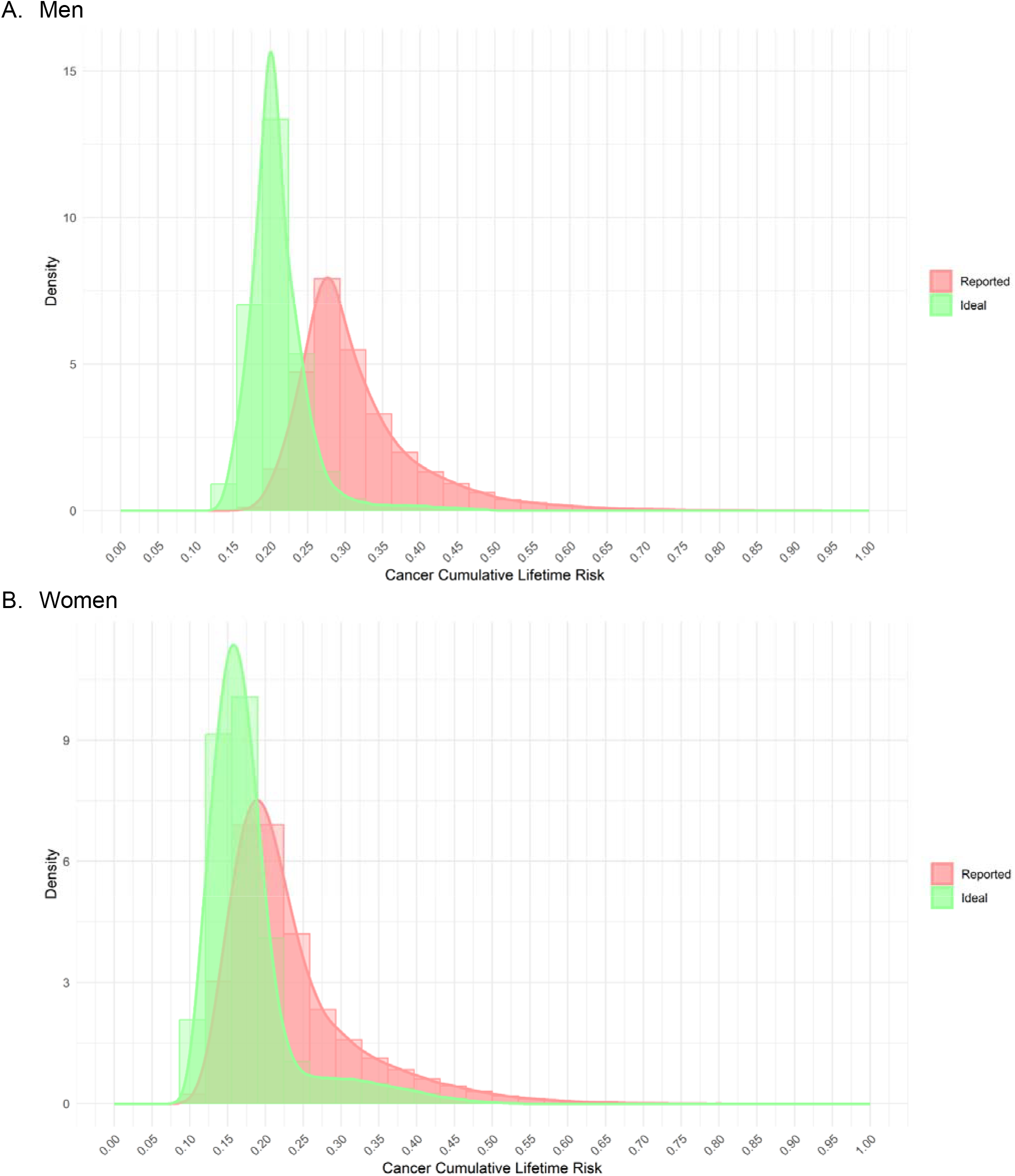
Distribution of lifetime all-cancer risk in the UK Biobank cohort based on reported risk factors and with all modifiable risk factors set to ideal values

The lifetime risk of cancer at the 90^th^ percentile (42.61%) was 1.4x higher compared to the median and 1.8x higher than the 10^th^ percentile (23.78%) for men. Similarly, the lifetime risk of cancer at the 90^th^ percentile (35.46%) was 1.7x higher compared to the median and 2.3x higher than the 10^th^ percentile (15.28%) for women.

If all modifiable risk factors were set to the ideal state, the overall risk distribution had a lower average risk and decreased spread. The median risk decreased to 20.45% for men (IQR 3.85%) and 16.51% for women (IQR 4.93%).

For men, median 10-year all-cancer risk increased as age increased from 40-49 (2.55%, IQR 1.54%), to 50-59 (7.47%, IQR 3.68%), to 60-70 (13.88%, IQR 5.02%, **Figure 2; Supplemental Table 4**). A similar pattern was observed for women (age 40-49: median 3.95%, IQR 1.78%; 50-59 median 6.27%, IQR 2.78%; 60-70 median 8.88%, IQR 3.75%). There was considerable overlap between age groups, with men aged 50-59 at the 90^th^ percentile having greater risk (11.91%) than men aged 60-70 at the 25^th^ percentile (11.80%), and women aged 40-49 at the 90^th^ percentile having greater risk (7.41%) than women aged 50-59 at the 60^th^ percentile (6.79%) and women aged 60-70 at the 20^th^ percentile (7.29%).

**Figure 2.**
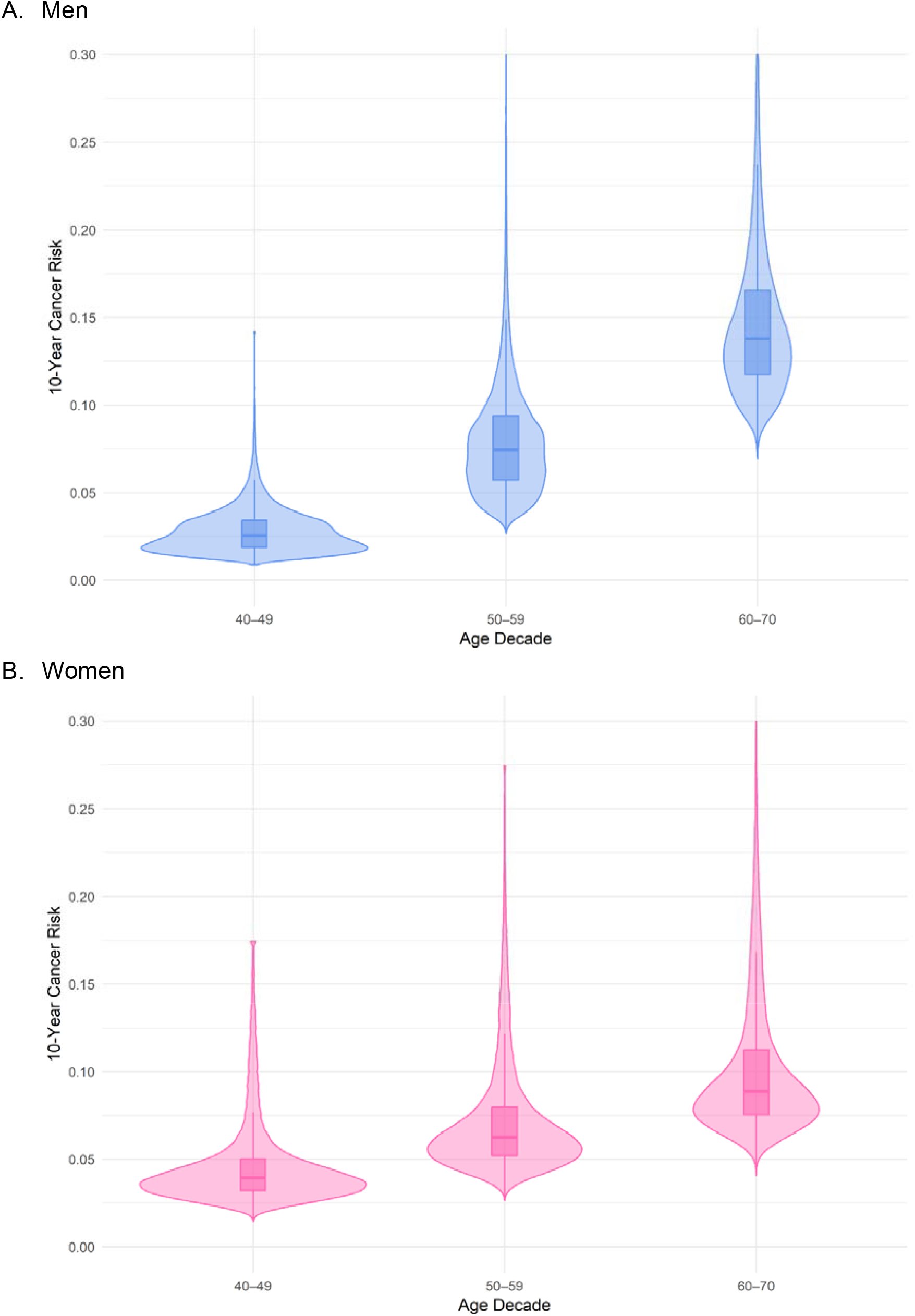
Distribution of 10-year all-cancer risk in the UK Biobank cohort by decade

## Discussion

We project lifetime risk of cancer across the UKB cohort and demonstrate for the first time how a multitude of factors other than age and sex create substantial heterogeneity in cancer risk. These results build on prior work estimating population-wide absolute breast cancer risk by examining all cancer.^13^ We find that the wide variability in lifetime cancer risk narrows, and median cancer risk is substantially lower if all individuals have ideal values for modifiable risk factors. Importantly, median 10-year cancer risk increases by decade of life, but the distribution of cancer risk overlaps considerably across decades.

Our findings have implications for multicancer screening guidelines as data on MCEDs and novel imaging emerge. Current population-wide screening recommendations rely largely on age, sex, and smoking history for lung cancer, neglecting other risk factors such as UV exposure, obesity, physical inactivity, alcohol use, and diet.^10^ Our findings demonstrate that the utility of multicancer screening may vary widely according to a multitude of factors. Future guidelines should evaluate performance of multicancer testing in subgroups, such as those with family history, certain genetic mutations, or environmental exposures.

Our results also inform clinicians counseling patients today. Prior work suggests approximately 40% of cancer risk is attributable to modifiable factors, though this did not account for overlap among risk factors at the population level.^10^ Our study accounts for the nature and magnitude of overlap for risk factors found in the UKB and extends this work by demonstrating how healthy behaviors modify the distribution of lifetime risk at the population level. In our study, median lifetime risk drops by 21% for women and 31% for men if ideal values are substituted. This suggests that a substantial number of cancers may be prevented if behavior change is undertaken over a lifetime. While individual near-term cancer risks may be low—especially at younger ages—quantifying lifetime all-cancer risk may help patients better understand the impact of behavior change and motivate prevention efforts.

The major strengths of our study over other multi-cancer risk studies are the inclusion of a larger number of cancers and risk factors and the ability to project lifetime cancer risk.^14–16^ However, several limitations exist. First, our model requires validation in more diverse populations. Second, we could not account for collinearity among risk factors absent from the UKB, though others have taken a similar approach.^10^ Finally, we assumed that risk factors remained constant within individuals over time.

## Conclusion

We find that lifetime cancer risk varies widely across the UKB cohort, but that this risk can be meaningfully lower with healthy behaviors. Furthermore, we find considerable overlap in 10-year risk between age groups, suggesting that future multicancer screening guidelines should consider factors beyond age and sex as more evidence emerges.

## Data Availability

Study data cannot be shared due to restrictions on data sharing in the data use agreement with the UK Biobank.

## Glossary

MCED: multi-cancer early detection

UKB: UK Biobank

## Acknowledgment

None

## References

1. Gelhorn H, Ross MM, Kansal AR, Fung ET, Seiden MV, Krucien N, Chung KC. Patient Preferences for Multi-Cancer Early Detection (MCED) Screening Tests. Patient. Jan 2023;16(1):43–56. doi:10.1007/s40271-022-00589-5

2. Gram EG, Copp T, Ransohoff DF, Plüddemann A, Kramer BS, Woloshin S, Shih P. Direct-to-consumer tests: emerging trends are cause for concern. BMJ. 2024;387:e080460. doi:10.1136/bmj-2024-080460

3. Wan JCM, Sasieni P, Rosenfeld N. Promises and pitfalls of multi-cancer early detection using liquid biopsy tests. Nat Rev Clin Oncol. Jun 13 2025;doi:10.1038/s41571-025-01033-x

4. Rubinstein WS, Patriotis C, Dickherber A, et al. Cancer screening with multicancer detection tests: A translational science review. CA Cancer J Clin. Jul-Aug 2024;74(4):368–382. doi:10.3322/caac.21833

5. Krainak DM. 21 CFR 892.2050. Medical image management and processing system. April 6, 2023. https://www.accessdata.fda.gov/cdrh_docs/pdf23/K230264.pdf. Accessed December 12, 2024.

6. Rubin R. Questions Swirl Around Screening for Multiple Cancers With a Single Blood Test. Jama. Apr 2 2024;331(13):1077–1080. doi:10.1001/jama.2024.1018

7. Bycroft C, Freeman C, Petkova D, et al. The UK Biobank resource with deep phenotyping and genomic data. Nature. Oct 2018;562(7726):203–209. doi:10.1038/s41586-018-0579-z

8. Office for National Statistics, “Deaths broken down by age, sex, area and cause of death.” https://www.ons.gov.uk/peoplepopulationandcommunity/birthsdeathsandmarriages/deaths. Accessed 3/20/2025.

9. Cancer Research UK, “Cancer Statistics for the UK.” https://www.cancerresearchuk.org/health-professional/cancer-statistics-for-the-uk, Accessed 3/20/2025.

10. Islami F, Marlow EC, Thomson B, et al. Proportion and number of cancer cases and deaths attributable to potentially modifiable risk factors in the United States, 2019. CA Cancer J Clin. Sep-Oct 2024;74(5):405–432. doi:10.3322/caac.21858

11. Pal Choudhury P, Maas P, Wilcox A, et al. iCARE: An R package to build, validate and apply absolute risk models. PLoS One. 2020;15(2):e0228198. doi:10.1371/journal.pone.0228198

12. Balasubramanian JB, Choudhury PP, Mukhopadhyay S, Ahearn T, Chatterjee N, García-Closas M, Almeida JS. Wasm-iCARE: a portable and privacy-preserving web module to build, validate, and apply absolute risk models. JAMIA Open. Jul 2024;7(2):ooae055. doi:10.1093/jamiaopen/ooae055

13. Maas P, Barrdahl M, Joshi AD, et al. Breast Cancer Risk From Modifiable and Nonmodifiable Risk Factors Among White Women in the United States. JAMA Oncol. Oct 1 2016;2(10):1295–1302. doi:10.1001/jamaoncol.2016.1025

14. Hippisley-Cox J, Coupland C. Development and validation of risk prediction algorithms to estimate future risk of common cancers in men and women: prospective cohort study. BMJ Open. 2015;5(3):e007825. doi:10.1136/bmjopen-2015-007825

15. Kachuri L, Graff RE, Smith-Byrne K, et al. Pan-cancer analysis demonstrates that integrating polygenic risk scores with modifiable risk factors improves risk prediction. Nature Communications. 2020/11/27 2020;11(1):6084. doi:10.1038/s41467-020-19600-4

16. Kim ES, Scharpf RB, Garcia-Closas M, Visvanathan K, Velculescu VE, Chatterjee N. Potential utility of risk stratification for multicancer screening with liquid biopsy tests. npj Precision Oncology. 2023/04/22 2023;7(1):39. doi:10.1038/s41698-023-00377-w

