## Supplemental Material for "Projecting individualized probabilities of lifetime all-cancer risk"

**Supplemental Materials**

**eMethods. Supplemental Methods**

*Overall Approach*

Our all-cancer model is comprised of 38 individual cancer and sex-specific models. We first conducted a comprehensive literature review of meta-analyses, large-scale cohort studies, and existing cancer-specific models to identify established risk factors contributing to the risk of developing 21 common cancers. We then developed cancer-specific models to simultaneously select and quantify the impact of risk factors identified in the literature to account for collinearity. Finally, we used the Python version of the iCARE package to quantify and project absolute cancer risks for individual participants.^1^

*Model Development*

The purpose of developing the cancer risk model was not to identify novel cancer risk factors or develop and validate novel cancer risk models, but rather to account for collinearity among established risk factors identified in the literature. For cancer-specific model derivation, separate models were developed for each cancer type by sex using the UK Biobank cohort who did not have cancer at time of enrollment (eFigure 1). Individuals under age 40 or over age 70 at time of recruitment or with inconsistent information regarding sex (e.g. listed as male, but had ovaries removed) were excluded due to suspected data quality issues.

First, we identified cancer diagnoses, which would serve as the outcomes for each of our cancer-specific models, based on the presence of any established International Classification of Diseases, 10^th^ Edition codes recorded during any hospitalization for the patient since recruitment. Next, we next identified variables in the UK biobank corresponding to established literature review-based risk factors as potential exposures. We then checked the distribution of continuous variables and transformed them as needed (e.g. square-root function, log function, etc.) to approximate a normal distribution. All variables had <5% missing (except for pack-years among current/former smokers, duration of moderate and vigorous activity, age at first live birth for females, and water intake), and missing values were imputed to the mean (single imputation).

To address multicollinearity while preserving coefficient estimates, we used a hybrid approach. Regularization methods (Lasso and Elastic Net) were applied to identify key predictors but were not used for coefficient estimation to avoid shrinkage.^2-4^ Selected variables were further evaluated for collinearity using Variance Inflation Factor (VIF), with a threshold of VIF > 10 guiding the exclusion or aggregation of variables.^5,6^ Backward stepwise selection was subsequently used to refine the final variable set included in the model.^7,8^ While stepwise selection can exacerbate multicollinearity by retaining one variable and discarding another highly correlated variable,^9^ we mitigated this by evaluating excluded variables for their conceptual importance and potential proxy effects, ensuring that critical risk factors were retained.^10^ We also retained coefficients derived from published studies for variables with strong literature-based evidence to maintain consistency with established risk factor estimates. Notably, we did not include genetic cancer syndromes (e.g. BRCA1, PALB2) or rare occupational (e.g. nickel) or toxin exposures (e.g. dioxin TCDD) given that this information was not consistently available for UK Biobank participants, correlation with other included variables (e.g. family history or smoking status), and their relatively infrequent occurrence such that inclusion would not affect population-level distributions significantly. Final coefficient estimation was conducted using logistic regression, ensuring the interpretability and stability of the parameter estimates.^11,12^

Separate models for lung cancer were developed for current smokers and never smokers due to the substantial impact of smoking on modifying lung cancer risk and the potential for distinct risk factors between smokers and nonsmokers. Based on the proportion of the population and breakdown of lung cancer cases by smoking status,^13^ we multiplied the aggregate UK lung cancer incidence files by 2.5 to reflect risk for current smokers (which account for 36.7% of lung cancers and 14.7% of the population) and by 0.21 to reflect risk for never smokers (which account for 12.5% of lung cancers and 60% of the population). Lung cancer risk for former smokers was modeled similarly to that of current smokers, though risk was linearly attenuated based on the number of years since quitting smoking until it matched that of never smokers at 15 years, in line with guidelines that no longer recommend screening among former smokers who quit greater than 15 years ago.^14^ Former smokers who quit more than 15 years ago or who had missing information with regards to age at quit were modeled as nonsmokers.

*Absolute risk computation*

To feed risk model parameters into the iCARE framework, a compilation of risk model inputs was prepared as JSON and text files to generate cancer-specific log-odds estimates and formula inputs, respectively. Age was included as a covariate in all risk models during development but was subsequently removed given that iCARE incorporates age separately in computing absolute risk. Reference data were prepared by appending the final UK Biobank sample with simulated risk factors not available in the UK Biobank generated using a fixed random seed to ensure reproducibility.^15^ To ensure using the whole sample as a reference, missing data were imputed with sex-specific means. Cancer-specific incidence by age was obtained from the Cancer Research UK database.^16^ All-cause mortality data by age was sourced from the UK Office for National Statistics and included as competing risks.^17^ Although age-specific mortality data was only available for England and Wales, 93% of the UK Biobank population came from these areas.

We computed the 10-year risk and the lifetime risk of developing each individual cancer up to age 85. For each participant, we used the age at recruitment to determine the starting age in the iCARE framework. Cancer-specific query risk factor inputs were further stratified by recruitment age into distinct files for faster iCARE processing. The starting age and interval length was computed from the age indicator in the file name of each of the age-and-cancer-specific stratified query files.

Cancer risk was computed through an iterative loop that processed the computation of participants’ absolute risk scores for each individual cancer type, accounting for the unique subgroups in lung cancers. The results were systematically aggregated in the lifetime and 10-year risk profiles. Each computation was stratified by cancer-subgroup with outputs stored in structured data dictionaries, enabling robust risk assessment and ensuring consistency in computations across all cancer types.

All-cancer risk was calculated as the complement of the product of risk of not getting any cancer (Ci) across all cancer types according to the formula:

Risk_All_cancer_ = (1 – (1-Risk_C1_) x (1-Risk_C2_) x (1-Risk_C3_) … (1-Risk_Ci_))

This calculation assumes statistical independence across different cancers, conditional on the inclusion of the comprehensive set of established risk factors included in the disparate models. To explore age-related risk trends, data were stratified into age decades: 40–49, 50–59, and 60–70 years.

Lastly, modifiable risk factors were adjusted to their ideal values (eTable2), and lifetime risks were then recomputed to project the population impact of adopting ideal risk profiles. This allowed for population-level insights into the impact of potential lifestyle and environmental changes.

Separate summary statistics were computed for all cancer risk within the female and male groups, including means, medians, standard deviations, minima, maxima, interquartile ranges (Q1 and Q3), and deciles (10th to 90th percentiles). Separate datasets were generated for both lifetime risk (up to age 85) and 10-year risk horizons.

Throughout the analysis, we applied quality control measures to maintain data integrity and computation accuracy. This included automated checks for the presence and format of required input files, as well as alignment checks to ensure consistency among iCARE inputs.

*Visualizations*

We created overlaid violin and box plots to visualize the distribution and peak of risk scores. For comparisons between reported query input values for participants and healthy risk profiles for modifiable risk factors, we generated overlaid density-scaled histograms and smoothed density plots. For improved visual interpretability, we removed outliers within each health group if the value exceeded Q3 +7 x IQR.^18^

*Software and Tools*

All analyses were completed using SAS 9.4, R v4.3.2, and Python v3.11.11. Pandas in Python facilitated data management and integration. Cloud-based workflows within Google Drive were used for the structured analysis pipeline.

**References**

1.	Balasubramanian JB, Choudhury PP, Mukhopadhyay S, Ahearn T, Chatterjee N, García-Closas M, Almeida JS. Wasm-iCARE: a portable and privacy-preserving web module to build, validate, and apply absolute risk models. *JAMIA Open*. Jul 2024;7(2):ooae055. doi:10.1093/jamiaopen/ooae055

2.	Tibshirani R. Regression shrinkage and selection via the lasso. *Journal of the Royal Statistical Society Series B: Statistical Methodology*. 1996;58(1):267-288.

3.	Zou H, Hastie T. Regularization and variable selection via the elastic net. *Journal of the Royal Statistical Society Series B: Statistical Methodology*. 2005;67(2):301-320.

4.	Vatcheva KP, Lee M, McCormick JB, Rahbar MH. Multicollinearity in regression analyses conducted in epidemiologic studies. *Epidemiology (Sunnyvale, Calif)*. 2016;6(2):227.

5.	O’brien RM. A caution regarding rules of thumb for variance inflation factors. *Quality & quantity*. 2007;41:673-690.

6.	Myers RH. *Classical and modern regression with applications*. vol 2. Duxbury press Belmont, CA; 1990.

7.	Harrell FE. *Regression modeling strategies: with applications to linear models, logistic regression, and survival analysis*. vol 608. Springer; 2001.

8.	Johnson KD, Lin D, Ungar LH, Foster DP, Stine RA. A Risk Ratio Comparison of $ l_0 $ and $ l_1 $ Penalized Regression. *arXiv preprint arXiv:151006319*. 2015;

9.	Lydersen S. Statistical review: frequently given comments updated. *Annals of the Rheumatic Diseases*. 2025;

10.	Mickey RM, Greenland S. The impact of confounder selection criteria on effect estimation. *American journal of epidemiology*. 1989;129(1):125-137.

11.	Hosmer Jr DW, Lemeshow S, Sturdivant RX. *Applied logistic regression*. John Wiley & Sons; 2013.

12.	Long JS, Freese J. *Regression models for categorical dependent variables using Stata*. vol 7. Stata press; 2006.

13.	Siegel DA, Fedewa SA, Henley SJ, Pollack LA, Jemal A. Proportion of Never Smokers Among Men and Women With Lung Cancer in 7 US States. *JAMA Oncol*. Feb 1 2021;7(2):302-304. doi:10.1001/jamaoncol.2020.6362

14.	Force UPST. Screening for Lung Cancer: US Preventive Services Task Force Recommendation Statement. *JAMA*. 2021;325(10):962-970. doi:10.1001/jama.2021.1117

15.	Gentleman R, Temple Lang D. Statistical analyses and reproducible research. *Journal of Computational and Graphical Statistics*. 2007;16(1):1-23.

16.	Cancer Research UK, "Cancer Statistics for the UK." <https://www.cancerresearchuk.org/health-professional/cancer-statistics-for-the-uk>, Accessed 3/20/2025.

17.	Office for National Statistics, "Deaths broken down by age, sex, area and cause of death." <https://www.ons.gov.uk/peoplepopulationandcommunity/birthsdeathsandmarriages/deaths>. Accessed 3/20/2025.

18.	Tukey JW. *Exploratory data analysis*. vol 2. Springer; 1977.

**eFigure 1. Study Consort**

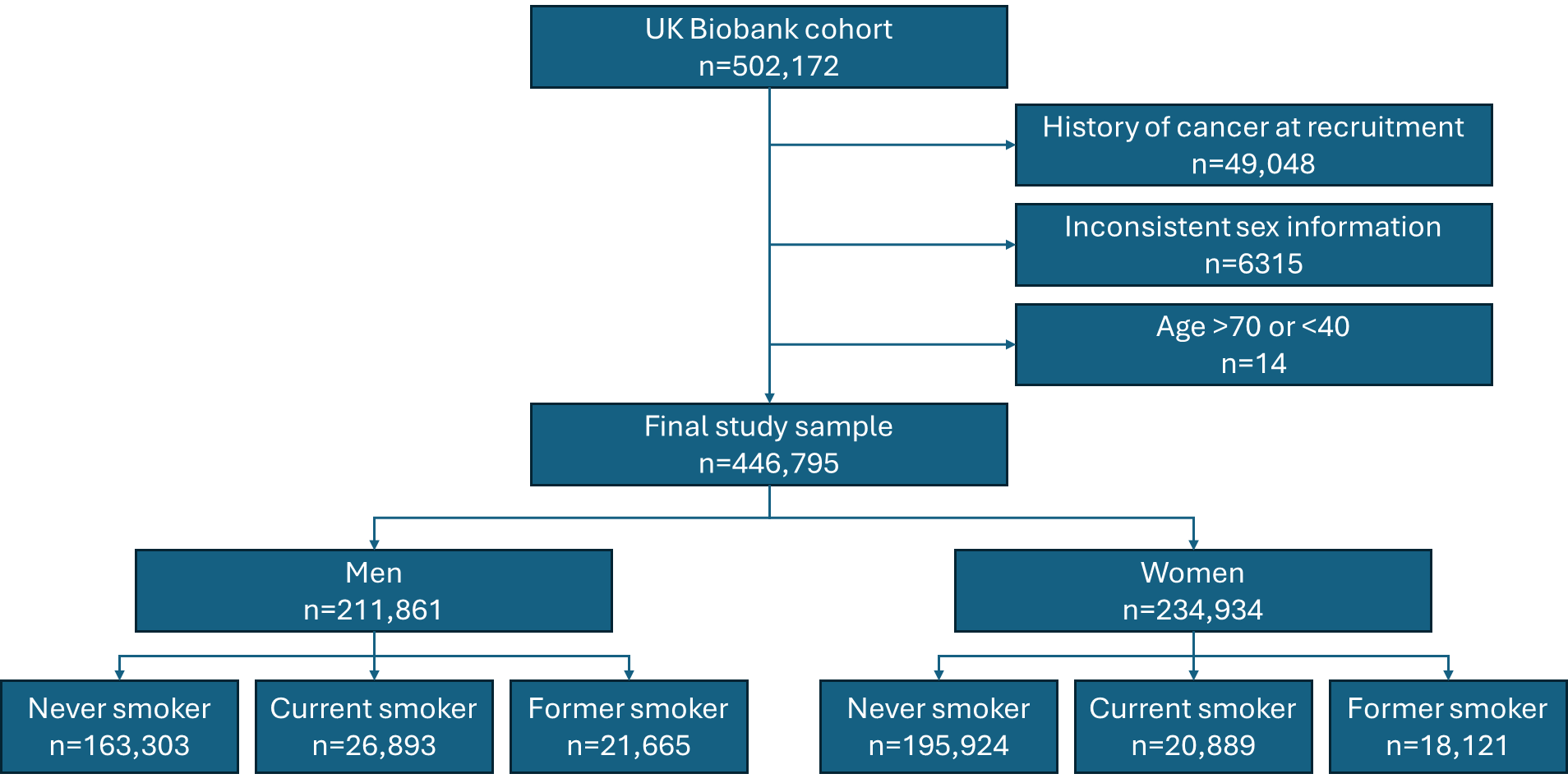

**eTable 1. Model details and sources**

| **Cancer model** | **Variable** | **Variable type** | **Odds ratio** | **Odds ratio source** | **Mean** | **Prevalence** | **Reference** |
| --- | --- | --- | --- | --- | --- | --- | --- |
| Bladder Female | Body mass index | Continuous | 1.00 | UK Biobank | 29.80 |  | Bhaskaran K, Douglas I, Forbes H, dos-Santos-Silva I, Leon DA, Smeeth L. Body-mass index and risk of 22 specific cancers: a population-based cohort study of 5·24 million UK adults. Lancet. Aug 30 2014;384(9945):755-65. doi:10.1016/s0140-6736(14)60892-8 |
| Bladder Female | Chronic cystitis | Binary | 4.90 | Literature | 0.07 | 0.03 | Groah SL, Weitzenkamp DA, Lammertse DP, Whiteneck GG, Lezotte DC, Hamman RF. Excess risk of bladder cancer in spinal cord injury: evidence for an association between indwelling catheter use and bladder cancer. Arch Phys Med Rehabil. Mar 2002;83(3):346-51. doi:10.1053/apmr.2002.29653 |
| Bladder Female | Chronic kidney disease | Binary | 2.63 | UK Biobank | 0.04 |  | Stengel B. Chronic kidney disease and cancer: a troubling connection. J Nephrol. May-Jun 2010;23(3):253-62. |
| Bladder Female | Diabetes mellitus type 2 | Binary | 1.28 | UK Biobank | 0.04 |  | Xu Y, Huo R, Chen X, Yu X. Diabetes mellitus and the risk of bladder cancer: A PRISMA-compliant meta-analysis of cohort studies. Medicine (Baltimore). Nov 2017;96(46):e8588. doi:10.1097/md.0000000000008588 Peng XF, Meng XY, Wei C, et al. The association between metabolic syndrome and bladder cancer susceptibility and prognosis: an updated comprehensive evidence synthesis of 95 observational studies involving 97,795,299 subjects. Cancer Manag Res. 2018;10:6263-6274. doi:10.2147/cmar.S181178 |
| Bladder Female | Glasses of water per day (cube-root transformation) | Continuous | 0.88 | UK Biobank | 1.42 |  | Michaud DS, Spiegelman D, Clinton SK, Rimm EB, Curhan GC, Willett WC, Giovannucci EL. Fluid intake and the risk of bladder cancer in men. N Engl J Med. May 6 1999;340(18):1390-7. doi:10.1056/nejm199905063401803 |
| Bladder Female | Human papilloma virus infection history | Binary | 2.84 | Literature | 0.43 | 0.43 | Li N, Yang L, Zhang Y, Zhao P, Zheng T, Dai M. Human papillomavirus infection and bladder cancer risk: a meta-analysis. J Infect Dis. Jul 15 2011;204(2):217-23. doi:10.1093/infdis/jir248 |
| Bladder Female | Maternal smoking around birth | Binary | 1.14 | UK Biobank | 0.25 |  | Jiang X, Yuan JM, Skipper PL, Tannenbaum SR, Yu MC. Environmental tobacco smoke and bladder cancer risk in never smokers of Los Angeles County. Cancer Res. Aug 1 2007;67(15):7540-5. doi:10.1158/0008-5472.Can-07-0048 |
| Bladder Female | Pack-years smoking history (fifth-root transformation) | Continuous | 1.34 | UK Biobank | 0.00 |  | Freedman ND, Silverman DT, Hollenbeck AR, Schatzkin A, Abnet CC. Association between smoking and risk of bladder cancer among men and women. Jama. Aug 17 2011;306(7):737-45. doi:10.1001/jama.2011.1142 Cumberbatch MG, Rota M, Catto JW, La Vecchia C. The Role of Tobacco Smoke in Bladder and Kidney Carcinogenesis: A Comparison of Exposures and Meta-analysis of Incidence and Mortality Risks. Eur Urol. Sep 2016;70(3):458-66. doi:10.1016/j.eururo.2015.06.042 Jubber I, Ong S, Bukavina L, et al. Epidemiology of Bladder Cancer in 2023: A Systematic Review of Risk Factors. Eur Urol. Aug 2023;84(2):176-190. doi:10.1016/j.eururo.2023.03.029 |
| Bladder Female | Secondhand smoke at home or work > 10 years | Binary | 1.97 | Literature | 0.20 | 0.20 | Jiang X, Yuan JM, Skipper PL, Tannenbaum SR, Yu MC. Environmental tobacco smoke and bladder cancer risk in never smokers of Los Angeles County. Cancer Res. Aug 1 2007;67(15):7540-5. doi:10.1158/0008-5472.Can-07-0048 |
| Bladder Female | Smoking (current) | Binary | 1.65 | UK Biobank | 0.09 |  | Freedman ND, Silverman DT, Hollenbeck AR, Schatzkin A, Abnet CC. Association between smoking and risk of bladder cancer among men and women. Jama. Aug 17 2011;306(7):737-45. doi:10.1001/jama.2011.1142 Cumberbatch MG, Rota M, Catto JW, La Vecchia C. The Role of Tobacco Smoke in Bladder and Kidney Carcinogenesis: A Comparison of Exposures and Meta-analysis of Incidence and Mortality Risks. Eur Urol. Sep 2016;70(3):458-66. doi:10.1016/j.eururo.2015.06.042 Jubber I, Ong S, Bukavina L, et al. Epidemiology of Bladder Cancer in 2023: A Systematic Review of Risk Factors. Eur Urol. Aug 2023;84(2):176-190. doi:10.1016/j.eururo.2023.03.029 |
| Bladder Male | Body mass index | Continuous | 1.01 | UK Biobank | 29.40 |  | Bhaskaran K, Douglas I, Forbes H, dos-Santos-Silva I, Leon DA, Smeeth L. Body-mass index and risk of 22 specific cancers: a population-based cohort study of 5·24 million UK adults. Lancet. Aug 30 2014;384(9945):755-65. doi:10.1016/s0140-6736(14)60892-8 |
| Bladder Male | Chronic cystitis | Binary | 4.90 | Literature | 0.07 | 0.03 | Groah SL, Weitzenkamp DA, Lammertse DP, Whiteneck GG, Lezotte DC, Hamman RF. Excess risk of bladder cancer in spinal cord injury: evidence for an association between indwelling catheter use and bladder cancer. Arch Phys Med Rehabil. Mar 2002;83(3):346-51. doi:10.1053/apmr.2002.29653 |
| Bladder Male | Chronic kidney disease | Binary | 2.46 | UK Biobank | 0.05 |  | Stengel B. Chronic kidney disease and cancer: a troubling connection. J Nephrol. May-Jun 2010;23(3):253-62. |
| Bladder Male | Glasses of water per day (cube-root transformation) | Continuous | 0.98 | UK Biobank | 1.42 |  | Michaud DS, Spiegelman D, Clinton SK, Rimm EB, Curhan GC, Willett WC, Giovannucci EL. Fluid intake and the risk of bladder cancer in men. N Engl J Med. May 6 1999;340(18):1390-7. doi:10.1056/nejm199905063401803 |
| Bladder Male | Human papilloma virus infection history | Binary | 2.84 | Literature | 0.43 | 0.43 | Li N, Yang L, Zhang Y, Zhao P, Zheng T, Dai M. Human papillomavirus infection and bladder cancer risk: a meta-analysis. J Infect Dis. Jul 15 2011;204(2):217-23. doi:10.1093/infdis/jir248 |
| Bladder Male | Maternal smoking around birth | Binary | 1.04 | UK Biobank | 0.26 |  | Jiang X, Yuan JM, Skipper PL, Tannenbaum SR, Yu MC. Environmental tobacco smoke and bladder cancer risk in never smokers of Los Angeles County. Cancer Res. Aug 1 2007;67(15):7540-5. doi:10.1158/0008-5472.Can-07-0048 |
| Bladder Male | Nonsteroidal anti-inflammatory drug use | Binary | 0.92 | UK Biobank | 0.12 |  | Daugherty SE, Pfeiffer RM, Sigurdson AJ, et al. Nonsteroidal antiinflammatory drugs and bladder cancer: a pooled analysis. Am J Epidemiol. Apr 1 2011;173(7):721-30. doi:10.1093/aje/kwq437 |
| Bladder Male | Pack-years smoking history (fifth-root transformation) | Continuous | 1.30 | UK Biobank | 0.00 |  | Freedman ND, Silverman DT, Hollenbeck AR, Schatzkin A, Abnet CC. Association between smoking and risk of bladder cancer among men and women. Jama. Aug 17 2011;306(7):737-45. doi:10.1001/jama.2011.1142 Cumberbatch MG, Rota M, Catto JW, La Vecchia C. The Role of Tobacco Smoke in Bladder and Kidney Carcinogenesis: A Comparison of Exposures and Meta-analysis of Incidence and Mortality Risks. Eur Urol. Sep 2016;70(3):458-66. doi:10.1016/j.eururo.2015.06.042 Jubber I, Ong S, Bukavina L, et al. Epidemiology of Bladder Cancer in 2023: A Systematic Review of Risk Factors. Eur Urol. Aug 2023;84(2):176-190. doi:10.1016/j.eururo.2023.03.029 |
| Bladder Male | Smoking (current) | Binary | 1.51 | UK Biobank | 0.13 |  | Freedman ND, Silverman DT, Hollenbeck AR, Schatzkin A, Abnet CC. Association between smoking and risk of bladder cancer among men and women. Jama. Aug 17 2011;306(7):737-45. doi:10.1001/jama.2011.1142 Cumberbatch MG, Rota M, Catto JW, La Vecchia C. The Role of Tobacco Smoke in Bladder and Kidney Carcinogenesis: A Comparison of Exposures and Meta-analysis of Incidence and Mortality Risks. Eur Urol. Sep 2016;70(3):458-66. doi:10.1016/j.eururo.2015.06.042 Jubber I, Ong S, Bukavina L, et al. Epidemiology of Bladder Cancer in 2023: A Systematic Review of Risk Factors. Eur Urol. Aug 2023;84(2):176-190. doi:10.1016/j.eururo.2023.03.029 |
| Brain Female | Aspirin use | Binary | 1.00 | UK Biobank | 0.10 |  | Amirian ES, Ostrom QT, Armstrong GN, et al. Aspirin, NSAIDs, and Glioma Risk: Original Data from the Glioma International Case-Control Study and a Meta-analysis. Cancer Epidemiol Biomarkers Prev. Mar 2019;28(3):555-562. doi:10.1158/1055-9965.Epi-18-0702 |
| Brain Female | Asthma | Binary | 0.68 | Literature | 0.08 | 0.08 | Linos E, Raine T, Alonso A, Michaud D. Atopy and risk of brain tumors: a meta-analysis. J Natl Cancer Inst. Oct 17 2007;99(20):1544-50. doi:10.1093/jnci/djm170 |
| Brain Female | CT scan of head as a child | Binary | 1.50 | Literature | 0.02 | 0.02 | Pearce MS, Salotti JA, Little MP, et al. Radiation exposure from CT scans in childhood and subsequent risk of leukaemia and brain tumours: a retrospective cohort study. Lancet. Aug 4 2012;380(9840):499-505. doi:10.1016/s0140-6736(12)60815-0 |
| Brain Female | Cups of tea daily (fifth-root transformation) | Continuous | 0.92 | UK Biobank | 0.97 |  | Song Y, Wang Z, Jin Y, Guo J. Association between tea and coffee consumption and brain cancer risk: an updated meta-analysis. World J Surg Oncol. Mar 15 2019;17(1):51. doi:10.1186/s12957-019-1591-y |
| Brain Female | Eczema | Binary | 0.69 | Literature | 0.07 | 0.07 | Linos E, Raine T, Alonso A, Michaud D. Atopy and risk of brain tumors: a meta-analysis. J Natl Cancer Inst. Oct 17 2007;99(20):1544-50. doi:10.1093/jnci/djm170 |
| Brain Female | Seasonal allergies | Binary | 0.61 | Literature | 0.30 | 0.30 | Linos E, Raine T, Alonso A, Michaud D. Atopy and risk of brain tumors: a meta-analysis. J Natl Cancer Inst. Oct 17 2007;99(20):1544-50. doi:10.1093/jnci/djm170 |
| Brain Female | Servings of vegetables per day (square-root transformation) | Continuous | 0.91 | UK Biobank | 1.22 |  | Chen H, Ward MH, Tucker KL, et al. Diet and risk of adult glioma in eastern Nebraska, United States. Cancer Causes Control. Sep 2002;13(7):647-55. doi:10.1023/a:1019527225197 Giles GG, McNeil JJ, Donnan G, et al. Dietary factors and the risk of glioma in adults: results of a case-control study in Melbourne, Australia. Int J Cancer. Nov 1 1994;59(3):357-62. doi:10.1002/ijc.2910590311 |
| Brain Male | Aspirin use | Binary | 0.86 | UK Biobank | 0.19 |  | Amirian ES, Ostrom QT, Armstrong GN, et al. Aspirin, NSAIDs, and Glioma Risk: Original Data from the Glioma International Case-Control Study and a Meta-analysis. Cancer Epidemiol Biomarkers Prev. Mar 2019;28(3):555-562. doi:10.1158/1055-9965.Epi-18-0702 |
| Brain Male | Asthma | Binary | 0.68 | Literature | 0.08 | 0.08 | Linos E, Raine T, Alonso A, Michaud D. Atopy and risk of brain tumors: a meta-analysis. J Natl Cancer Inst. Oct 17 2007;99(20):1544-50. doi:10.1093/jnci/djm170 |
| Brain Male | CT scan of head as a child | Binary | 1.50 | Literature | 0.02 | 0.02 | Pearce MS, Salotti JA, Little MP, et al. Radiation exposure from CT scans in childhood and subsequent risk of leukaemia and brain tumours: a retrospective cohort study. Lancet. Aug 4 2012;380(9840):499-505. doi:10.1016/s0140-6736(12)60815-0 |
| Brain Male | Cups of coffee daily (fifth-root transformation) | Continuous | 0.84 | UK Biobank | 0.90 |  | Song Y, Wang Z, Jin Y, Guo J. Association between tea and coffee consumption and brain cancer risk: an updated meta-analysis. World J Surg Oncol. Mar 15 2019;17(1):51. doi:10.1186/s12957-019-1591-y |
| Brain Male | Cups of tea daily (fifth-root transformation) | Continuous | 0.92 | UK Biobank | 0.97 |  | Song Y, Wang Z, Jin Y, Guo J. Association between tea and coffee consumption and brain cancer risk: an updated meta-analysis. World J Surg Oncol. Mar 15 2019;17(1):51. doi:10.1186/s12957-019-1591-y |
| Brain Male | Eczema | Binary | 0.69 | Literature | 0.07 | 0.07 | Linos E, Raine T, Alonso A, Michaud D. Atopy and risk of brain tumors: a meta-analysis. J Natl Cancer Inst. Oct 17 2007;99(20):1544-50. doi:10.1093/jnci/djm170 |
| Brain Male | Seasonal allergies | Binary | 0.61 | Literature | 0.30 | 0.30 | Linos E, Raine T, Alonso A, Michaud D. Atopy and risk of brain tumors: a meta-analysis. J Natl Cancer Inst. Oct 17 2007;99(20):1544-50. doi:10.1093/jnci/djm170 |
| Brain Male | Servings of fatty fish per week (square-root transformation) | Continuous | 0.90 | UK Biobank | 1.16 |  | Lee KH, Seong HJ, Kim G, et al. Consumption of Fish and ω-3 Fatty Acids and Cancer Risk: An Umbrella Review of Meta-Analyses of Observational Studies. Adv Nutr. Sep 1 2020;11(5):1134-1149. doi:10.1093/advances/nmaa055 |
| Brain Male | Servings of processed meat (bacon, ham, sausages, burgers, nuggets, etc.) per week (square-root transformation) | Continuous | 1.01 | UK Biobank | 1.64 |  | Giles GG, McNeil JJ, Donnan G, et al. Dietary factors and the risk of glioma in adults: results of a case-control study in Melbourne, Australia. Int J Cancer. Nov 1 1994;59(3):357-62. doi:10.1002/ijc.2910590311 |
| Brain Male | Servings of red meat (beef, lamb, or pork) per week (square-root transformation) | Binary | 1.07 | UK Biobank | 2.12 |  | Giles GG, McNeil JJ, Donnan G, et al. Dietary factors and the risk of glioma in adults: results of a case-control study in Melbourne, Australia. Int J Cancer. Nov 1 1994;59(3):357-62. doi:10.1002/ijc.2910590311 |
| Brain Male | Servings of vegetables per day (square-root transformation) | Continuous | 0.99 | UK Biobank | 1.22 |  | Chen H, Ward MH, Tucker KL, et al. Diet and risk of adult glioma in eastern Nebraska, United States. Cancer Causes Control. Sep 2002;13(7):647-55. doi:10.1023/a:1019527225197 Giles GG, McNeil JJ, Donnan G, et al. Dietary factors and the risk of glioma in adults: results of a case-control study in Melbourne, Australia. Int J Cancer. Nov 1 1994;59(3):357-62. doi:10.1002/ijc.2910590311 |
| Brain Male | Statin use | Binary | 0.91 | UK Biobank | 0.23 |  | Gaist D, Andersen L, Hallas J, Toft Sørensen H, Schrøder HD, Friis S. Use of statins and risk of glioma: a nationwide case–control study in Denmark. British Journal of Cancer. 2013/02/01 2013;108(3):715-720. doi:10.1038/bjc.2012.536 |
| Breast Female | Age first live birth <25 | Binary | 0.90 | UK Biobank | 0.00 |  | Menarche, menopause, and breast cancer risk: individual participant meta-analysis, including 118 964 women with breast cancer from 117 epidemiological studies. Lancet Oncol. Nov 2012;13(11):1141-51. doi:10.1016/s1470-2045(12)70425-4 |
| Breast Female | Age of menarche (<=11, 12-13, >13) | Ordinal | 0.95 | UK Biobank | 1.00 |  | Menarche, menopause, and breast cancer risk: individual participant meta-analysis, including 118 964 women with breast cancer from 117 epidemiological studies. Lancet Oncol. Nov 2012;13(11):1141-51. doi:10.1016/s1470-2045(12)70425-4 |
| Breast Female | Age of menopause > 55 | Binary | 1.02 | UK Biobank | 0.00 |  | Collaborative Group on Hormonal Factors in Breast Cancer. Breast cancer and hormone replacement therapy: collaborative reanalysis of data from 51 epidemiological studies of 52,705 women with breast cancer and 108,411 women without breast cancer. Lancet. 11;350(9084):1047-59, 1997 |
| Breast Female | Alcohol consumption (2-3 drinks/day) | Binary | 1.31 | Literature | 0.12 | 0.12 | Bagnardi V, Blangiardo M, La Vecchia C, Corrao G. Alcohol consumption and the risk of cancer: a meta-analysis. Alcohol Res Health. 2001;25(4):263-70. https://www.ncbi.nlm.nih.gov/pmc/articles/PMC6705703/ |
| Breast Female | Alcohol consumption (4+ drinks/day) | Binary | 1.67 | Literature | 0.05 | 0.05 | Bagnardi V, Blangiardo M, La Vecchia C, Corrao G. Alcohol consumption and the risk of cancer: a meta-analysis. Alcohol Res Health. 2001;25(4):263-70. https://www.ncbi.nlm.nih.gov/pmc/articles/PMC6705703/ |
| Breast Female | Benign breast disease | Binary | 1.57 | Literature | 0.61 | 0.61 | Colditz GA, Rosner B. Cumulative risk of breast cancer to age 70 years according to risk factor status: data from the Nurses' Health Study. Am J Epidemiol. Nov 15 2000;152(10):950-64. doi:10.1093/aje/152.10.950 |
| Breast Female | Breastfeeding 12-17 months | Binary | 0.96 | Literature | 0.20 | 0.20 | Breast cancer and breastfeeding: collaborative reanalysis of individual data from 47 epidemiological studies in 30 countries, including 50302 women with breast cancer and 96973 women without the disease. Lancet. Jul 20 2002;360(9328):187-95. doi:10.1016/s0140-6736(02)09454-0 |
| Breast Female | Breastfeeding 18-23 months | Binary | 0.94 | Literature | 0.09 | 0.09 | Breast cancer and breastfeeding: collaborative reanalysis of individual data from 47 epidemiological studies in 30 countries, including 50302 women with breast cancer and 96973 women without the disease. Lancet. Jul 20 2002;360(9328):187-95. doi:10.1016/s0140-6736(02)09454-0 |
| Breast Female | Breastfeeding 24-29 months | Binary | 0.91 | Literature | 0.04 | 0.04 | Breast cancer and breastfeeding: collaborative reanalysis of individual data from 47 epidemiological studies in 30 countries, including 50302 women with breast cancer and 96973 women without the disease. Lancet. Jul 20 2002;360(9328):187-95. doi:10.1016/s0140-6736(02)09454-0 |
| Breast Female | Breastfeeding 30-35 months | Binary | 0.89 | Literature | 0.04 | 0.04 | Breast cancer and breastfeeding: collaborative reanalysis of individual data from 47 epidemiological studies in 30 countries, including 50302 women with breast cancer and 96973 women without the disease. Lancet. Jul 20 2002;360(9328):187-95. doi:10.1016/s0140-6736(02)09454-0 |
| Breast Female | Breastfeeding 36+ months | Binary | 0.87 | Literature | 0.07 | 0.07 | Breast cancer and breastfeeding: collaborative reanalysis of individual data from 47 epidemiological studies in 30 countries, including 50302 women with breast cancer and 96973 women without the disease. Lancet. Jul 20 2002;360(9328):187-95. doi:10.1016/s0140-6736(02)09454-0 |
| Breast Female | Breastfeeding 6-11 months | Binary | 0.98 | Literature | 0.28 | 0.09 | Breast cancer and breastfeeding: collaborative reanalysis of individual data from 47 epidemiological studies in 30 countries, including 50302 women with breast cancer and 96973 women without the disease. Lancet. Jul 20 2002;360(9328):187-95. doi:10.1016/s0140-6736(02)09454-0 |
| Breast Female | Extremely dense breasts (>75%) | Binary | 4.70 | Literature | 0.03 | 0.10 | Boyd NF, Guo H, Martin LJ, et al. Mammographic density and the risk and detection of breast cancer. N Engl J Med. Jan 18 2007;356(3):227-36. doi:10.1056/NEJMoa062790 |
| Breast Female | Family history of breast cancer in first degree relative | Binary | 1.12 | UK Biobank | 0.07 |  | Familial breast cancer: collaborative reanalysis of individual data from 52 epidemiological studies including 58,209 women with breast cancer and 101,986 women without the disease. Lancet. Oct 27 2001;358(9291):1389-99. doi:10.1016/s0140-6736(01)06524-2 |
| Breast Female | Height (in cm) | Continuous | 1.02 | UK Biobank | 162.36 |  | Ahlgren M, Melbye M, Wohlfahrt J, Sørensen TI. Growth patterns and the risk of breast cancer in women. N Engl J Med. Oct 14 2004;351(16):1619-26. doi:10.1056/NEJMoa040576 Green J, Cairns BJ, Casabonne D, Wright FL, Reeves G, Beral V. Height and cancer incidence in the Million Women Study: prospective cohort, and meta-analysis of prospective studies of height and total cancer risk. Lancet Oncol. Aug 2011;12(8):785-94. doi:10.1016/s1470-2045(11)70154-1 |
| Breast Female | Hormone replacement therapy | Binary | 1.03 | UK Biobank | 0.39 |  | Collaborative Group on Hormonal Factors in Breast Cancer. Breast cancer and hormone replacement therapy: collaborative reanalysis of data from 51 epidemiological studies of 52,705 women with breast cancer and 108,411 women without breast cancer. Lancet. 11;350(9084):1047-59, 1997 |
| Breast Female | Minutes of moderate activity daily (<30 minutes, 30-59 minutes, 60-89 minutes, >90 minutes) | Ordinal | 0.96 | UK Biobank | 1.00 |  | Hardefeldt PJ, Penninkilampi R, Edirimanne S, Eslick GD. Physical Activity and Weight Loss Reduce the Risk of Breast Cancer: A Meta-analysis of 139 Prospective and Retrospective Studies. Clin Breast Cancer. Aug 2018;18(4):e601-e612. doi:10.1016/j.clbc.2017.10.010 |
| Breast Female | Minutes of vigorous activity daily (<30 minutes, 30-59 minutes, 60-89 minutes, >90 minutes) | Ordinal | 0.98 | UK Biobank | 1.00 |  | Hardefeldt PJ, Penninkilampi R, Edirimanne S, Eslick GD. Physical Activity and Weight Loss Reduce the Risk of Breast Cancer: A Meta-analysis of 139 Prospective and Retrospective Studies. Clin Breast Cancer. Aug 2018;18(4):e601-e612. doi:10.1016/j.clbc.2017.10.010 |
| Breast Female | Nonsteroidal anti-inflammatory drug use | Binary | 0.95 | UK Biobank | 0.17 |  | de Pedro M, Baeza S, Escudero MT, Dierssen-Sotos T, Gómez-Acebo I, Pollán M, Llorca J. Effect of COX-2 inhibitors and other non-steroidal inflammatory drugs on breast cancer risk: a meta-analysis. Breast Cancer Res Treat. Jan 2015;149(2):525-36. doi:10.1007/s10549-015-3267-9 |
| Breast Female | Oopherectomy | Binary | 0.89 | UK Biobank | 0.07 |  | Hassan H, Allen I, Sofianopoulou E, et al. Long-term outcomes of hysterectomy with bilateral salpingo-oophorectomy: a systematic review and meta-analysis. Am J Obstet Gynecol. Jan 2024;230(1):44-57. doi:10.1016/j.ajog.2023.06.043 |
| Breast Female | Pack-years smoking history (fifth-root transformation) | Continuous | 1.02 | UK Biobank | 0.00 |  | Gram IT, Park SY, Kolonel LN, Maskarinec G, Wilkens LR, Henderson BE, Le Marchand L. Smoking and Risk of Breast Cancer in a Racially/Ethnically Diverse Population of Mainly Women Who Do Not Drink Alcohol: The MEC Study. Am J Epidemiol. Dec 1 2015;182(11):917-25. doi:10.1093/aje/kwv092 |
| Breast Female | Secondhand smoke exposure (ever) | Binary | 1.20 | Literature | 0.18 | 0.18 | Macacu A, Autier P, Boniol M, Boyle P. Active and passive smoking and risk of breast cancer: a meta-analysis. Breast Cancer Res Treat. Nov 2015;154(2):213-24. doi:10.1007/s10549-015-3628-4 |
| Breast Female | Servings of fruit per day (square-root transformation | Continuous | 0.94 | UK Biobank | 1.00 |  | Brennan SF, Cantwell MM, Cardwell CR, Velentzis LS, Woodside JV. Dietary patterns and breast cancer risk: a systematic review and meta-analysis. Am J Clin Nutr. May 2010;91(5):1294-302. doi:10.3945/ajcn.2009.28796 Farvid MS, Chen WY, Michels KB, Cho E, Willett WC, Eliassen AH. Fruit and vegetable consumption in adolescence and early adulthood and risk of breast cancer: population based cohort study. Bmj. May 11 2016;353:i2343. doi:10.1136/bmj.i2343 Chlebowski RT, Aragaki AK, Anderson GL, et al. Dietary Modification and Breast Cancer Mortality: Long-Term Follow-Up of the Women's Health Initiative Randomized Trial. J Clin Oncol. May 1 2020;38(13):1419-1428. doi:10.1200/jco.19.00435 |
| Breast Female | Servings of red meat (beef, lamb, or pork) per week (square-root transformation) | Binary | 1.05 | UK Biobank | 2.12 |  | Cho E, Chen WY, Hunter DJ, Stampfer MJ, Colditz GA, Hankinson SE, Willett WC. Red meat intake and risk of breast cancer among premenopausal women. *Arch Intern Med*. Nov 13 2006;166(20):2253-9. doi:10.1001/archinte.166.20.2253 |
| Breast Female | Smoking (current) | Binary | 1.09 | UK Biobank | 0.09 |  | Gram IT, Park SY, Kolonel LN, Maskarinec G, Wilkens LR, Henderson BE, Le Marchand L. Smoking and Risk of Breast Cancer in a Racially/Ethnically Diverse Population of Mainly Women Who Do Not Drink Alcohol: The MEC Study. Am J Epidemiol. Dec 1 2015;182(11):917-25. doi:10.1093/aje/kwv092 |
| Cervical Female | Body mass index | Continuous | 1.02 | UK Biobank | 29.80 |  | Norat T, Abar L, Vieira AR, et al. World Cancer Research Fund International Systematic Literature Review: The Associations between Food, Nutrition and Physical Activity and the Risk of Cervical Cancer. 2018. https://www.wcrf.org/wp-content/uploads/2021/02/Cervical-cancer-slr.pdf |
| Cervical Female | Family history of cervical cancer in first degree relative | Binary | 2.00 | Literature | 0.03 | 0.03 | Zelmanowicz Ade M, Schiffman M, Herrero R, et al. Family history as a co-factor for adenocarcinoma and squamous cell carcinoma of the uterine cervix: results from two studies conducted in Costa Rica and the United States. Int J Cancer. Sep 10 2005;116(4):599-605. doi:10.1002/ijc.21048 |
| Cervical Female | Human papilloma virus vaccine before age 17 | Binary | 0.17 | Literature | 0.17 | 0.17 | Lei J, Ploner A, Elfström KM, et al. HPV Vaccination and the Risk of Invasive Cervical Cancer. N Engl J Med. Oct 1 2020;383(14):1340-1348. doi:10.1056/NEJMoa1917338 |
| Cervical Female | Human papilloma virus vaccine before between age 17-30 | Binary | 0.47 | Literature | 0.34 | 0.34 | Lei J, Ploner A, Elfström KM, et al. HPV Vaccination and the Risk of Invasive Cervical Cancer. N Engl J Med. Oct 1 2020;383(14):1340-1348. doi:10.1056/NEJMoa1917338 |
| Cervical Female | Hysterectomy | Binary | 0.01 | Literature | 0.17 | 0.17 | Hassan H, Allen I, Sofianopoulou E, et al. Long-term outcomes of hysterectomy with bilateral salpingo-oophorectomy: a systematic review and meta-analysis. Am J Obstet Gynecol. Jan 2024;230(1):44-57. doi:10.1016/j.ajog.2023.06.043 |
| Cervical Female | Intrauterine device (any) | Binary | 0.55 | Literature | 0.14 | 0.14 | Castellsagué X, Díaz M, Vaccarella S, et al. Intrauterine device use, cervical infection with human papillomavirus, and risk of cervical cancer: a pooled analysis of 26 epidemiological studies. Lancet Oncol. Oct 2011;12(11):1023-31. doi:10.1016/s1470-2045(11)70223-6 |
| Cervical Female | Number of live births | Continuous | 1.03 | UK Biobank | 1.00 |  | Comparison of risk factors for invasive squamous cell carcinoma and adenocarcinoma of the cervix: collaborative reanalysis of individual data on 8,097 women with squamous cell carcinoma and 1,374 women with adenocarcinoma from 12 epidemiological studies. Int J Cancer. Feb 15 2007;120(4):885-91. doi:10.1002/ijc.22357 |
| Cervical Female | Smoking (current) | Binary | 1.50 | UK Biobank | 0.09 |  | Appleby P, Beral V, Berrington de González A, et al. Carcinoma of the cervix and tobacco smoking: collaborative reanalysis of individual data on 13,541 women with carcinoma of the cervix and 23,017 women without carcinoma of the cervix from 23 epidemiological studies. Int J Cancer. Mar 15 2006;118(6):1481-95. doi:10.1002/ijc.21493 |
| Colon Female | Alcohol consumption (2-3 drinks/day) | Binary | 1.14 | Literature | 0.12 | 0.12 | Bagnardi V, Blangiardo M, La Vecchia C, Corrao G. Alcohol consumption and the risk of cancer: a meta-analysis. Alcohol Res Health. 2001;25(4):263-70. https://www.ncbi.nlm.nih.gov/pmc/articles/PMC6705703/ |
| Colon Female | Alcohol consumption (4+ drinks/day) | Binary | 1.21 | Literature | 0.05 | 0.05 | Bagnardi V, Blangiardo M, La Vecchia C, Corrao G. Alcohol consumption and the risk of cancer: a meta-analysis. Alcohol Res Health. 2001;25(4):263-70. https://www.ncbi.nlm.nih.gov/pmc/articles/PMC6705703/ |
| Colon Female | Body mass index | Continuous | 1.01 | UK Biobank | 29.80 |  | Karahalios A, English DR, Simpson JA. Weight change and risk of colorectal cancer: a systematic review and meta-analysis. Am J Epidemiol. Jun 1 2015;181(11):832-45. doi:10.1093/aje/kwu357 |
| Colon Female | Calcium supplementation (1200mg-2000mg daily) | Binary | 0.80 | Literature | 0.49 | 0.50 | Shaukat A, Scouras N, Schünemann HJ. Role of supplemental calcium in the recurrence of colorectal adenomas: a metaanalysis of randomized controlled trials. Am J Gastroenterol. Feb 2005;100(2):390-4. doi:10.1111/j.1572-0241.2005.41220.x |
| Colon Female | Cups of coffee daily (fifth-root transformation) | Continuous | 0.94 | UK Biobank | 0.85 |  | Sinha R, Cross AJ, Daniel CR, et al. Caffeinated and decaffeinated coffee and tea intakes and risk of colorectal cancer in a large prospective study. Am J Clin Nutr. Aug 2012;96(2):374-81. doi:10.3945/ajcn.111.031328 Schmit SL, Rennert HS, Rennert G, Gruber SB. Coffee Consumption and the Risk of Colorectal Cancer. Cancer Epidemiol Biomarkers Prev. Apr 2016;25(4):634-9. doi:10.1158/1055-9965.Epi-15-0924 |
| Colon Female | Cystic fibrosis | Binary | 10.91 | Literature | 0.01 | 0.01 | Yamada A, Komaki Y, Komaki F, Micic D, Zullow S, Sakuraba A. Risk of gastrointestinal cancers in patients with cystic fibrosis: a systematic review and meta-analysis. Lancet Oncol. Jun 2018;19(6):758-767. doi:10.1016/s1470-2045(18)30188-8 |
| Colon Female | Diabetes mellitus type 2 | Binary | 1.25 | UK Biobank | 0.04 |  | Yuhara H, Steinmaus C, Cohen SE, Corley DA, Tei Y, Buffler PA. Is diabetes mellitus an independent risk factor for colon cancer and rectal cancer? Am J Gastroenterol. Nov 2011;106(11):1911-21; quiz 1922. doi:10.1038/ajg.2011.301 |
| Colon Female | Family history of colorectal cancer in first degree relative | Binary | 1.08 | Literature | 0.12 | 0.05 | Tuohy TM, Rowe KG, Mineau GP, Pimentel R, Burt RW, Samadder NJ. Risk of colorectal cancer and adenomas in the families of patients with adenomas: a population-based study in Utah. Cancer. Jan 1 2014;120(1):35-42. doi:10.1002/cncr.28227 Taylor DP, Stoddard GJ, Burt RW, Williams MS, Mitchell JA, Haug PJ, Cannon-Albright LA. How well does family history predict who will get colorectal cancer? Implications for cancer screening and counseling. Genet Med. May 2011;13(5):385-91. doi:10.1097/GIM.0b013e3182064384 Taylor DP, Burt RW, Williams MS, Haug PJ, Cannon-Albright LA. Population-based family history-specific risks for colorectal cancer: a constellation approach. Gastroenterology. Mar 2010;138(3):877-85. doi:10.1053/j.gastro.2009.11.044 |
| Colon Female | Glasses of water per day (cube-root transformation) | Continuous | 0.94 | UK Biobank | 1.42 |  | Shannon J, White E, Shattuck AL, Potter JD. Relationship of food groups and water intake to colon cancer risk. Cancer Epidemiol Biomarkers Prev. Jul 1996;5(7):495-502. https://www.researchgate.net/profile/Jackilen-Shannon/publication/14378713_Relationship_of_food_groups_and_water_intake_to_colon_cancer_risk/links/5890c8b4aca272f9a556beff/Relationship-of-food-groups-and-water-intake-to-colon-cancer-risk.pdf |
| Colon Female | Height (in cm) | Continuous | 1.01 | UK Biobank | 162.36 |  | Green J, Cairns BJ, Casabonne D, Wright FL, Reeves G, Beral V. Height and cancer incidence in the Million Women Study: prospective cohort, and meta-analysis of prospective studies of height and total cancer risk. Lancet Oncol. Aug 2011;12(8):785-94. doi:10.1016/s1470-2045(11)70154-1 |
| Colon Female | Hormone replacement therapy | Binary | 0.94 | UK Biobank | 0.39 |  | Chlebowski RT, Wactawski-Wende J, Ritenbaugh C, et al. Estrogen plus progestin and colorectal cancer in postmenopausal women. N Engl J Med. Mar 4 2004;350(10):991-1004. doi:10.1056/NEJMoa032071 |
| Colon Female | Inflammatory bowel disease | Binary | 1.81 | UK Biobank | 0.01 |  | Olén O, Erichsen R, Sachs MC, et al. Colorectal cancer in ulcerative colitis: a Scandinavian population-based cohort study. Lancet. Jan 11 2020;395(10218):123-131. doi:10.1016/s0140-6736(19)32545-0 Ekbom A, Helmick C, Zack M, Adami HO. Ulcerative colitis and colorectal cancer. A population-based study. N Engl J Med. Nov 1 1990;323(18):1228-33. doi:10.1056/nejm199011013231802 |
| Colon Female | Minutes of vigorous activity daily (<30 minutes, 30-59 minutes, 60-89 minutes, >90 minutes) | Ordinal | 0.95 | UK Biobank | 1.00 |  | World Cancer Research Fund/American Institute for Cancer Research. Continuous Update Project Expert Report 2018. Diet, nutrition, physical activity, and colorectal cancer. 2017. https://www.wcrf.org/wp-content/uploads/2021/02/Colorectal-cancer-report.pdf  Norat T, Vieira AR, Abar L et al. World Cancer Research Fund International Systematic Literature Review: The Associations between Food, Nutrition and Physical Activity and the Risk of Colorectal Cancer. 2017. https://www.wcrf.org/wp-content/uploads/2021/02/colorectal-cancer-slr.pdf |
| Colon Female | Nonsteroidal anti-inflammatory drug use | Binary | 0.88 | UK Biobank | 0.17 |  | Rostom A, Dubé C, Lewin G, et al. Nonsteroidal anti-inflammatory drugs and cyclooxygenase-2 inhibitors for primary prevention of colorectal cancer: a systematic review prepared for the U.S. Preventive Services Task Force. Ann Intern Med. Mar 6 2007;146(5):376-89. doi:10.7326/0003-4819-146-5-200703060-00010 |
| Colon Female | Pack-years smoking history (fifth-root transformation) | Continuous | 1.11 | UK Biobank | 0.00 |  | Botteri E, Iodice S, Bagnardi V, Raimondi S, Lowenfels AB, Maisonneuve P. Smoking and colorectal cancer: a meta-analysis. Jama. Dec 17 2008;300(23):2765-78. doi:10.1001/jama.2008.839 |
| Colon Female | Servings of red meat (beef, lamb, or pork) per week (square-root transformation) | Binary | 1.04 | UK Biobank | 2.12 |  | World Cancer Research Fund/American Institute for Cancer Research. Continuous Update Project Expert Report 2018. Diet, nutrition, physical activity, and colorectal cancer. 2017. https://www.wcrf.org/wp-content/uploads/2021/02/Colorectal-cancer-report.pdf  Norat T, Vieira AR, Abar L et al. World Cancer Research Fund International Systematic Literature Review: The Associations between Food, Nutrition and Physical Activity and the Risk of Colorectal Cancer. 2017. https://www.wcrf.org/wp-content/uploads/2021/02/colorectal-cancer-slr.pdf |
| Colon Female | Servings of vegetables per day (square-root transformation) | Continuous | 0.99 | UK Biobank | 1.22 |  | World Cancer Research Fund/American Institute for Cancer Research. Continuous Update Project Expert Report 2018. Diet, nutrition, physical activity, and colorectal cancer. 2017. https://www.wcrf.org/wp-content/uploads/2021/02/Colorectal-cancer-report.pdf. Norat T, Vieira AR, Abar L et al. World Cancer Research Fund International Systematic Literature Review: The Associations between Food, Nutrition and Physical Activity and the Risk of Colorectal Cancer. 2017. https://www.wcrf.org/wp-content/uploads/2021/02/colorectal-cancer-slr.pdf |
| Colon Female | Statin use | Binary | 0.90 | UK Biobank | 0.13 |  | Sacks FM, Pfeffer MA, Moye LA, et al. The effect of pravastatin on coronary events after myocardial infarction in patients with average cholesterol levels. Cholesterol and Recurrent Events Trial investigators. N Engl J Med. Oct 3 1996;335(14):1001-9. doi:10.1056/nejm199610033351401 Pedersen TR, Berg K, Cook TJ, et al. Safety and tolerability of cholesterol lowering with simvastatin during 5 years in the Scandinavian Simvastatin Survival Study. Arch Intern Med. Oct 14 1996;156(18):2085-92. |
| Colon Male | Alcohol consumption (2-3 drinks/day) | Binary | 1.14 | Literature | 0.12 | 0.12 | Bagnardi V, Blangiardo M, La Vecchia C, Corrao G. Alcohol consumption and the risk of cancer: a meta-analysis. Alcohol Res Health. 2001;25(4):263-70. https://www.ncbi.nlm.nih.gov/pmc/articles/PMC6705703/ |
| Colon Male | Alcohol consumption (4+ drinks/day) | Binary | 1.21 | Literature | 0.05 | 0.05 | Bagnardi V, Blangiardo M, La Vecchia C, Corrao G. Alcohol consumption and the risk of cancer: a meta-analysis. Alcohol Res Health. 2001;25(4):263-70. https://www.ncbi.nlm.nih.gov/pmc/articles/PMC6705703/ |
| Colon Male | Body mass index | Continuous | 1.03 | UK Biobank | 29.40 |  | Karahalios A, English DR, Simpson JA. Weight change and risk of colorectal cancer: a systematic review and meta-analysis. Am J Epidemiol. Jun 1 2015;181(11):832-45. doi:10.1093/aje/kwu357 |
| Colon Male | Calcium supplementation (1200mg-2000mg daily) | Binary | 0.80 | Literature | 0.49 | 0.50 | Shaukat A, Scouras N, Schünemann HJ. Role of supplemental calcium in the recurrence of colorectal adenomas: a metaanalysis of randomized controlled trials. Am J Gastroenterol. Feb 2005;100(2):390-4. doi:10.1111/j.1572-0241.2005.41220.x |
| Colon Male | Cystic fibrosis | Binary | 10.91 | Literature | 0.01 | 0.01 | Yamada A, Komaki Y, Komaki F, Micic D, Zullow S, Sakuraba A. Risk of gastrointestinal cancers in patients with cystic fibrosis: a systematic review and meta-analysis. Lancet Oncol. Jun 2018;19(6):758-767. doi:10.1016/s1470-2045(18)30188-8 |
| Colon Male | Family history of colorectal cancer in first degree relative | Binary | 1.09 | UK Biobank | 0.12 |  | Tuohy TM, Rowe KG, Mineau GP, Pimentel R, Burt RW, Samadder NJ. Risk of colorectal cancer and adenomas in the families of patients with adenomas: a population-based study in Utah. Cancer. Jan 1 2014;120(1):35-42. doi:10.1002/cncr.28227 Taylor DP, Stoddard GJ, Burt RW, Williams MS, Mitchell JA, Haug PJ, Cannon-Albright LA. How well does family history predict who will get colorectal cancer? Implications for cancer screening and counseling. Genet Med. May 2011;13(5):385-91. doi:10.1097/GIM.0b013e3182064384 Taylor DP, Burt RW, Williams MS, Haug PJ, Cannon-Albright LA. Population-based family history-specific risks for colorectal cancer: a constellation approach. Gastroenterology. Mar 2010;138(3):877-85. doi:10.1053/j.gastro.2009.11.044 |
| Colon Male | Height (in cm) | Continuous | 1.01 | UK Biobank | 175.48 |  | Green J, Cairns BJ, Casabonne D, Wright FL, Reeves G, Beral V. Height and cancer incidence in the Million Women Study: prospective cohort, and meta-analysis of prospective studies of height and total cancer risk. Lancet Oncol. Aug 2011;12(8):785-94. doi:10.1016/s1470-2045(11)70154-1 |
| Colon Male | Inflammatory bowel disease | Binary | 1.66 | UK Biobank | 0.02 |  | Olén O, Erichsen R, Sachs MC, et al. Colorectal cancer in ulcerative colitis: a Scandinavian population-based cohort study. Lancet. Jan 11 2020;395(10218):123-131. doi:10.1016/s0140-6736(19)32545-0 Ekbom A, Helmick C, Zack M, Adami HO. Ulcerative colitis and colorectal cancer. A population-based study. N Engl J Med. Nov 1 1990;323(18):1228-33. doi:10.1056/nejm199011013231802 |
| Colon Male | Minutes of moderate activity daily (<30 minutes, 30-59 minutes, 60-89 minutes, >90 minutes) | Ordinal | 0.95 | UK Biobank | 1.00 |  | World Cancer Research Fund/American Institute for Cancer Research. Continuous Update Project Expert Report 2018. Diet, nutrition, physical activity, and colorectal cancer. 2017. https://www.wcrf.org/wp-content/uploads/2021/02/Colorectal-cancer-report.pdf  Norat T, Vieira AR, Abar L et al. World Cancer Research Fund International Systematic Literature Review: The Associations between Food, Nutrition and Physical Activity and the Risk of Colorectal Cancer. 2017. https://www.wcrf.org/wp-content/uploads/2021/02/colorectal-cancer-slr.pdf |
| Colon Male | Minutes of vigorous activity daily (<30 minutes, 30-59 minutes, 60-89 minutes, >90 minutes) | Ordinal | 0.99 | UK Biobank | 1.00 |  | World Cancer Research Fund/American Institute for Cancer Research. Continuous Update Project Expert Report 2018. Diet, nutrition, physical activity, and colorectal cancer. 2017. https://www.wcrf.org/wp-content/uploads/2021/02/Colorectal-cancer-report.pdf  Norat T, Vieira AR, Abar L et al. World Cancer Research Fund International Systematic Literature Review: The Associations between Food, Nutrition and Physical Activity and the Risk of Colorectal Cancer. 2017. https://www.wcrf.org/wp-content/uploads/2021/02/colorectal-cancer-slr.pdf |
| Colon Male | Nonsteroidal anti-inflammatory drug use | Binary | 0.86 | UK Biobank | 0.12 |  | Rostom A, Dubé C, Lewin G, et al. Nonsteroidal anti-inflammatory drugs and cyclooxygenase-2 inhibitors for primary prevention of colorectal cancer: a systematic review prepared for the U.S. Preventive Services Task Force. Ann Intern Med. Mar 6 2007;146(5):376-89. doi:10.7326/0003-4819-146-5-200703060-00010 |
| Colon Male | Pack-years smoking history (fifth-root transformation) | Continuous | 1.10 | UK Biobank | 0.00 |  | Botteri E, Iodice S, Bagnardi V, Raimondi S, Lowenfels AB, Maisonneuve P. Smoking and colorectal cancer: a meta-analysis. Jama. Dec 17 2008;300(23):2765-78. doi:10.1001/jama.2008.839 |
| Colon Male | Servings of fatty fish per week (square-root transformation) | Continuous | 0.95 | UK Biobank | 1.16 |  | World Cancer Research Fund/American Institute for Cancer Research. Continuous Update Project Expert Report 2018. Diet, nutrition, physical activity, and colorectal cancer. 2017. https://www.wcrf.org/wp-content/uploads/2021/02/Colorectal-cancer-report.pdf  Norat T, Vieira AR, Abar L et al. World Cancer Research Fund International Systematic Literature Review: The Associations between Food, Nutrition and Physical Activity and the Risk of Colorectal Cancer. 2017. https://www.wcrf.org/wp-content/uploads/2021/02/colorectal-cancer-slr.pdf |
| Colon Male | Servings of fruit per day (square-root transformation | Continuous | 0.92 | UK Biobank | 1.00 |  | World Cancer Research Fund/American Institute for Cancer Research. Continuous Update Project Expert Report 2018. Diet, nutrition, physical activity, and colorectal cancer. 2017. https://www.wcrf.org/wp-content/uploads/2021/02/Colorectal-cancer-report.pdf  Norat T, Vieira AR, Abar L et al. World Cancer Research Fund International Systematic Literature Review: The Associations between Food, Nutrition and Physical Activity and the Risk of Colorectal Cancer. 2017. https://www.wcrf.org/wp-content/uploads/2021/02/colorectal-cancer-slr.pdf |
| Colon Male | Servings of processed meat (bacon, ham, sausages, burgers, nuggets, etc.) per week (square-root transformation) | Continuous | 1.08 | UK Biobank | 1.64 |  | World Cancer Research Fund/American Institute for Cancer Research. Continuous Update Project Expert Report 2018. Diet, nutrition, physical activity, and colorectal cancer. 2017. https://www.wcrf.org/wp-content/uploads/2021/02/Colorectal-cancer-report.pdf  Norat T, Vieira AR, Abar L et al. World Cancer Research Fund International Systematic Literature Review: The Associations between Food, Nutrition and Physical Activity and the Risk of Colorectal Cancer. 2017. https://www.wcrf.org/wp-content/uploads/2021/02/colorectal-cancer-slr.pdf |
| Colon Male | Servings of red meat (beef, lamb, or pork) per week (square-root transformation) | Binary | 1.12 | UK Biobank | 2.12 |  | World Cancer Research Fund/American Institute for Cancer Research. Continuous Update Project Expert Report 2018. Diet, nutrition, physical activity, and colorectal cancer. 2017. https://www.wcrf.org/wp-content/uploads/2021/02/Colorectal-cancer-report.pdf  Norat T, Vieira AR, Abar L et al. World Cancer Research Fund International Systematic Literature Review: The Associations between Food, Nutrition and Physical Activity and the Risk of Colorectal Cancer. 2017. https://www.wcrf.org/wp-content/uploads/2021/02/colorectal-cancer-slr.pdf |
| Colon Male | Servings of vegetables per day (square-root transformation) | Continuous | 0.99 | UK Biobank | 1.22 |  | World Cancer Research Fund/American Institute for Cancer Research. Continuous Update Project Expert Report 2018. Diet, nutrition, physical activity, and colorectal cancer. 2017. https://www.wcrf.org/wp-content/uploads/2021/02/Colorectal-cancer-report.pdf  Norat T, Vieira AR, Abar L et al. World Cancer Research Fund International Systematic Literature Review: The Associations between Food, Nutrition and Physical Activity and the Risk of Colorectal Cancer. 2017. https://www.wcrf.org/wp-content/uploads/2021/02/colorectal-cancer-slr.pdf |
| Colon Male | Statin use | Binary | 0.92 | UK Biobank | 0.23 |  | Sacks FM, Pfeffer MA, Moye LA, et al. The effect of pravastatin on coronary events after myocardial infarction in patients with average cholesterol levels. Cholesterol and Recurrent Events Trial investigators. N Engl J Med. Oct 3 1996;335(14):1001-9. doi:10.1056/nejm199610033351401 Pedersen TR, Berg K, Cook TJ, et al. Safety and tolerability of cholesterol lowering with simvastatin during 5 years in the Scandinavian Simvastatin Survival Study. Arch Intern Med. Oct 14 1996;156(18):2085-92. |
| Endometrial Female | Age of menarche (<=11, 12-13, >13) | Ordinal | 0.90 | UK Biobank | 1.00 |  | Setiawan VW, Yang HP, Pike MC, et al. Type I and II endometrial cancers: have they different risk factors? J Clin Oncol. Jul 10 2013;31(20):2607-18. doi:10.1200/jco.2012.48.2596 Katagiri R, Iwasaki M, Abe SK, et al. Reproductive Factors and Endometrial Cancer Risk Among Women. JAMA Netw Open. Sep 5 2023;6(9):e2332296. doi:10.1001/jamanetworkopen.2023.32296 |
| Endometrial Female | Age of menopause > 55 | Binary | 1.75 | UK Biobank | 0.00 |  | Katagiri R, Iwasaki M, Abe SK, et al. Reproductive Factors and Endometrial Cancer Risk Among Women. JAMA Netw Open. Sep 5 2023;6(9):e2332296. doi:10.1001/jamanetworkopen.2023.32296 |
| Endometrial Female | Any breastfeeding | Binary | 0.89 | Literature | 0.40 | 0.40 | Jordan SJ, Na R, Johnatty SE, et al. Breastfeeding and Endometrial Cancer Risk: An Analysis From the Epidemiology of Endometrial Cancer Consortium. Obstet Gynecol. Jun 2017;129(6):1059-1067. doi:10.1097/aog.0000000000002057 |
| Endometrial Female | Body mass index | Continuous | 1.10 | UK Biobank | 29.80 |  | Setiawan VW, Yang HP, Pike MC, et al. Type I and II endometrial cancers: have they different risk factors? J Clin Oncol. Jul 10 2013;31(20):2607-18. doi:10.1200/jco.2012.48.2596 |
| Endometrial Female | Cups of coffee daily (fifth-root transformation) | Continuous | 0.91 | UK Biobank | 0.85 |  | Bravi F, Scotti L, Bosetti C, Gallus S, Negri E, La Vecchia C, Tavani A. Coffee drinking and endometrial cancer risk: a metaanalysis of observational studies. Am J Obstet Gynecol. Feb 2009;200(2):130-5. doi:10.1016/j.ajog.2008.10.032 |
| Endometrial Female | Family history of colorectal cancer in first degree relative | Binary | 1.17 | Literature | 0.12 | 0.05 | Win AK, Reece JC, Ryan S. Family history and risk of endometrial cancer: a systematic review and meta-analysis. Obstet Gynecol. Jan 2015;125(1):89-98. doi:10.1097/aog.0000000000000563 |
| Endometrial Female | Family history of uterine cancer in first degree relative | Binary | 1.82 | Literature | 0.05 | 0.05 | Win AK, Reece JC, Ryan S. Family history and risk of endometrial cancer: a systematic review and meta-analysis. Obstet Gynecol. Jan 2015;125(1):89-98. doi:10.1097/aog.0000000000000563 |
| Endometrial Female | Hysterectomy | Binary | 0.01 | Literature | 0.17 | 0.17 | Hassan H, Allen I, Sofianopoulou E, et al. Long-term outcomes of hysterectomy with bilateral salpingo-oophorectomy: a systematic review and meta-analysis. Am J Obstet Gynecol. Jan 2024;230(1):44-57. doi:10.1016/j.ajog.2023.06.043 |
| Endometrial Female | Intrauterine device (hormonal) | Binary | 0.22 | Literature | 0.09 | 0.09 | Jareid M, Thalabard JC, Aarflot M, Bøvelstad HM, Lund E, Braaten T. Levonorgestrel-releasing intrauterine system use is associated with a decreased risk of ovarian and endometrial cancer, without increased risk of breast cancer. Results from the NOWAC Study. Gynecol Oncol. Apr 2018;149(1):127-132. doi:10.1016/j.ygyno.2018.02.006. |
| Endometrial Female | Intrauterine device (non-hormonal) | Binary | 0.54 | Literature | 0.05 | 0.05 | Beining RM, Dennis LK, Smith EM, Dokras A. Meta-Analysis of Intrauterine Device Use and Risk of Endometrial Cancer. Annals of Epidemiology. 2008/06/01/ 2008;18(6):492-499. doi:https://doi.org/10.1016/j.annepidem.2007.11.011 |
| Endometrial Female | Minutes of vigorous activity daily (<30 minutes, 30-59 minutes, 60-89 minutes, >90 minutes) | Ordinal | 0.97 | UK Biobank | 1.00 |  | World Cancer Research Fund/American Institute for Cancer Research. Continuous Update Project Expert Report 2018. Diet, nutrition, physical activity, and endometrial cancer. 2017. https://www.wcrf.org/wp-content/uploads/2021/02/Endometrial-cancer-report.pdf |
| Endometrial Female | Number of live births | Continuous | 0.84 | UK Biobank | 1.00 |  | Husby A, Wohlfahrt J, Melbye M. Pregnancy duration and endometrial cancer risk: nationwide cohort study. Bmj. Aug 14 2019;366:l4693. doi:10.1136/bmj.l4693 |
| Endometrial Female | Oral contraceptive use | Binary | 0.69 | UK Biobank | 0.81 |  | Endometrial cancer and oral contraceptives: an individual participant meta-analysis of 27 276 women with endometrial cancer from 36 epidemiological studies. Lancet Oncol. Sep 2015;16(9):1061-1070. doi:10.1016/s1470-2045(15)00212-0 Iversen L, Sivasubramaniam S, Lee AJ, Fielding S, Hannaford PC. Lifetime cancer risk and combined oral contraceptives: the Royal College of General Practitioners' Oral Contraception Study. Am J Obstet Gynecol. Jun 2017;216(6):580.e1-580.e9. doi:10.1016/j.ajog.2017.02.002 |
| Endometrial Female | Polycystic ovarian syndrome | Binary | 2.79 | Literature | 0.06 | 0.06 | Barry JA, Azizia MM, Hardiman PJ. Risk of endometrial, ovarian and breast cancer in women with polycystic ovary syndrome: a systematic review and meta-analysis. Hum Reprod Update. Sep-Oct 2014;20(5):748-58. doi:10.1093/humupd/dmu012 |
| Esophageal Female | Achalasia | Binary | 11.26 | UK Biobank | 0.00 |  | Sandler RS, Nyrén O, Ekbom A, Eisen GM, Yuen J, Josefsson S. The risk of esophageal cancer in patients with achalasia. A population-based study. Jama. Nov 1 1995;274(17):1359-62. |
| Esophageal Female | Alcohol consumption (2-3 drinks/day) | Binary | 1.51 | Literature | 0.12 | 0.12 | Bagnardi V, Blangiardo M, La Vecchia C, Corrao G. Alcohol consumption and the risk of cancer: a meta-analysis. Alcohol Res Health. 2001;25(4):263-70. https://www.ncbi.nlm.nih.gov/pmc/articles/PMC6705703/ |
| Esophageal Female | Alcohol consumption (4+ drinks/day) | Binary | 2.21 | Literature | 0.05 | 0.05 | Bagnardi V, Blangiardo M, La Vecchia C, Corrao G. Alcohol consumption and the risk of cancer: a meta-analysis. Alcohol Res Health. 2001;25(4):263-70. https://www.ncbi.nlm.nih.gov/pmc/articles/PMC6705703/ |
| Esophageal Female | Barrett's esophagus | Binary | 8.57 | UK Biobank | 0.01 |  | Hvid-Jensen F, Pedersen L, Drewes AM, Sørensen HT, Funch-Jensen P. Incidence of adenocarcinoma among patients with Barrett's esophagus. N Engl J Med. Oct 13 2011;365(15):1375-83. doi:10.1056/NEJMoa1103042 |
| Esophageal Female | Chronic hepatitis B | Binary | 5.30 | UK Biobank | 0.00 |  | Geng H, Xing Y, Zhang J, Cao K, Ye M, Wang G, Liu C. Association between viral infection other than human papillomavirus and risk of esophageal carcinoma: a comprehensive meta-analysis of epidemiological studies. Arch Virol. Jan 2022;167(1):1-20. doi:10.1007/s00705-021-05268-8 |
| Esophageal Female | Gastroesophageal reflux disease | Binary | 1.90 | UK Biobank | 0.12 |  | Lagergren J, Bergström R, Lindgren A, Nyrén O. Symptomatic gastroesophageal reflux as a risk factor for esophageal adenocarcinoma. N Engl J Med. Mar 18 1999;340(11):825-31. doi:10.1056/nejm199903183401101 |
| Esophageal Female | Minutes of vigorous activity daily (<30 minutes, 30-59 minutes, 60-89 minutes, >90 minutes) | Ordinal | 0.92 | UK Biobank | 1.00 |  | World Cancer Research Fund/American Institute for Cancer Research. Continuous Update Project Expert Report 2018. Diet, nutrition, physical activity, and oesophageal cancer. 2017. https://www.wcrf.org/wp-content/uploads/2021/02/oesophageal-cancer-report.pdf |
| Esophageal Female | Nonsteroidal anti-inflammatory drug use | Binary | 0.70 | UK Biobank | 0.17 |  | Liao LM, Vaughan TL, Corley DA, et al. Nonsteroidal anti-inflammatory drug use reduces risk of adenocarcinomas of the esophagus and esophagogastric junction in a pooled analysis. Gastroenterology. Mar 2012;142(3):442-452.e5; quiz e22-3. doi:10.1053/j.gastro.2011.11.019 |
| Esophageal Female | Pack-years smoking history (fifth-root transformation) | Continuous | 1.33 | UK Biobank | 0.00 |  | Cook MB, Kamangar F, Whiteman DC, et al. Cigarette smoking and adenocarcinomas of the esophagus and esophagogastric junction: a pooled analysis from the international BEACON consortium. J Natl Cancer Inst. Sep 8 2010;102(17):1344-53. doi:10.1093/jnci/djq289 Iribarren C, Tekawa IS, Sidney S, Friedman GD. Effect of cigar smoking on the risk of cardiovascular disease, chronic obstructive pulmonary disease, and cancer in men. N Engl J Med. Jun 10 1999;340(23):1773-80. doi:10.1056/nejm199906103402301 Randi G, Scotti L, Bosetti C, et al. Pipe smoking and cancers of the upper digestive tract. Int J Cancer. Nov 1 2007;121(9):2049-2051. doi:10.1002/ijc.22791 Freedman ND, Abnet CC, Caporaso NE, et al. Impact of changing US cigarette smoking patterns on incident cancer: risks of 20 smoking-related cancers among the women and men of the NIH-AARP cohort. Int J Epidemiol. Jun 2016;45(3):846-56. doi:10.1093/ije/dyv175 |
| Esophageal Female | Preference to drink very hot beverages (e.g. when >140 degrees F) | Binary | 1.51 | Literature | 0.15 | 0.15 | Islami F, Poustchi H, Pourshams A, et al. A prospective study of tea drinking temperature and risk of esophageal squamous cell carcinoma. Int J Cancer. Jan 1 2020;146(1):18-25. doi:10.1002/ijc.32220 |
| Esophageal Female | Smoking (current) | Binary | 1.59 | UK Biobank | 0.09 |  | Cook MB, Kamangar F, Whiteman DC, et al. Cigarette smoking and adenocarcinomas of the esophagus and esophagogastric junction: a pooled analysis from the international BEACON consortium. J Natl Cancer Inst. Sep 8 2010;102(17):1344-53. doi:10.1093/jnci/djq289 Iribarren C, Tekawa IS, Sidney S, Friedman GD. Effect of cigar smoking on the risk of cardiovascular disease, chronic obstructive pulmonary disease, and cancer in men. N Engl J Med. Jun 10 1999;340(23):1773-80. doi:10.1056/nejm199906103402301 Randi G, Scotti L, Bosetti C, et al. Pipe smoking and cancers of the upper digestive tract. Int J Cancer. Nov 1 2007;121(9):2049-2051. doi:10.1002/ijc.22791 Freedman ND, Abnet CC, Caporaso NE, et al. Impact of changing US cigarette smoking patterns on incident cancer: risks of 20 smoking-related cancers among the women and men of the NIH-AARP cohort. Int J Epidemiol. Jun 2016;45(3):846-56. doi:10.1093/ije/dyv175 |
| Esophageal Male | Achalasia | Binary | 4.35 | UK Biobank | 0.00 |  | Sandler RS, Nyrén O, Ekbom A, Eisen GM, Yuen J, Josefsson S. The risk of esophageal cancer in patients with achalasia. A population-based study. Jama. Nov 1 1995;274(17):1359-62. |
| Esophageal Male | Alcohol consumption (2-3 drinks/day) | Binary | 1.51 | Literature | 0.12 | 0.12 | Bagnardi V, Blangiardo M, La Vecchia C, Corrao G. Alcohol consumption and the risk of cancer: a meta-analysis. Alcohol Res Health. 2001;25(4):263-70. https://www.ncbi.nlm.nih.gov/pmc/articles/PMC6705703/ |
| Esophageal Male | Alcohol consumption (4+ drinks/day) | Binary | 2.21 | Literature | 0.05 | 0.05 | Bagnardi V, Blangiardo M, La Vecchia C, Corrao G. Alcohol consumption and the risk of cancer: a meta-analysis. Alcohol Res Health. 2001;25(4):263-70. https://www.ncbi.nlm.nih.gov/pmc/articles/PMC6705703/ |
| Esophageal Male | Aspirin use | Binary | 0.93 | UK Biobank | 0.19 |  | Liao LM, Vaughan TL, Corley DA, et al. Nonsteroidal anti-inflammatory drug use reduces risk of adenocarcinomas of the esophagus and esophagogastric junction in a pooled analysis. Gastroenterology. Mar 2012;142(3):442-452.e5; quiz e22-3. doi:10.1053/j.gastro.2011.11.019 |
| Esophageal Male | Barrett's esophagus | Binary | 9.21 | UK Biobank | 0.02 |  | Hvid-Jensen F, Pedersen L, Drewes AM, Sørensen HT, Funch-Jensen P. Incidence of adenocarcinoma among patients with Barrett's esophagus. N Engl J Med. Oct 13 2011;365(15):1375-83. doi:10.1056/NEJMoa1103042 |
| Esophageal Male | Chronic hepatitis B | Binary | 4.05 | UK Biobank | 0.00 |  | Geng H, Xing Y, Zhang J, Cao K, Ye M, Wang G, Liu C. Association between viral infection other than human papillomavirus and risk of esophageal carcinoma: a comprehensive meta-analysis of epidemiological studies. Arch Virol. Jan 2022;167(1):1-20. doi:10.1007/s00705-021-05268-8 |
| Esophageal Male | Gastroesophageal reflux disease | Binary | 1.75 | UK Biobank | 0.11 |  | Lagergren J, Bergström R, Lindgren A, Nyrén O. Symptomatic gastroesophageal reflux as a risk factor for esophageal adenocarcinoma. N Engl J Med. Mar 18 1999;340(11):825-31. doi:10.1056/nejm199903183401101 |
| Esophageal Male | Minutes of moderate activity daily (<30 minutes, 30-59 minutes, 60-89 minutes, >90 minutes) | Ordinal | 0.98 | UK Biobank | 1.00 |  | World Cancer Research Fund/American Institute for Cancer Research. Continuous Update Project Expert Report 2018. Diet, nutrition, physical activity, and oesophageal cancer. 2017. https://www.wcrf.org/wp-content/uploads/2021/02/oesophageal-cancer-report.pdf |
| Esophageal Male | Minutes of vigorous activity daily (<30 minutes, 30-59 minutes, 60-89 minutes, >90 minutes) | Ordinal | 0.96 | UK Biobank | 1.00 |  | World Cancer Research Fund/American Institute for Cancer Research. Continuous Update Project Expert Report 2018. Diet, nutrition, physical activity, and oesophageal cancer. 2017. https://www.wcrf.org/wp-content/uploads/2021/02/oesophageal-cancer-report.pdf |
| Esophageal Male | Nonsteroidal anti-inflammatory drug use | Binary | 0.86 | UK Biobank | 0.12 |  | Liao LM, Vaughan TL, Corley DA, et al. Nonsteroidal anti-inflammatory drug use reduces risk of adenocarcinomas of the esophagus and esophagogastric junction in a pooled analysis. Gastroenterology. Mar 2012;142(3):442-452.e5; quiz e22-3. doi:10.1053/j.gastro.2011.11.019 |
| Esophageal Male | Pack-years smoking history (fifth-root transformation) | Continuous | 1.34 | UK Biobank | 0.00 |  | Cook MB, Kamangar F, Whiteman DC, et al. Cigarette smoking and adenocarcinomas of the esophagus and esophagogastric junction: a pooled analysis from the international BEACON consortium. J Natl Cancer Inst. Sep 8 2010;102(17):1344-53. doi:10.1093/jnci/djq289 Iribarren C, Tekawa IS, Sidney S, Friedman GD. Effect of cigar smoking on the risk of cardiovascular disease, chronic obstructive pulmonary disease, and cancer in men. N Engl J Med. Jun 10 1999;340(23):1773-80. doi:10.1056/nejm199906103402301 Randi G, Scotti L, Bosetti C, et al. Pipe smoking and cancers of the upper digestive tract. Int J Cancer. Nov 1 2007;121(9):2049-2051. doi:10.1002/ijc.22791 Freedman ND, Abnet CC, Caporaso NE, et al. Impact of changing US cigarette smoking patterns on incident cancer: risks of 20 smoking-related cancers among the women and men of the NIH-AARP cohort. Int J Epidemiol. Jun 2016;45(3):846-56. doi:10.1093/ije/dyv175 |
| Esophageal Male | Preference to drink very hot beverages (e.g. when >140 degrees F) | Binary | 1.51 | Literature | 0.15 | 0.15 | Islami F, Poustchi H, Pourshams A, et al. A prospective study of tea drinking temperature and risk of esophageal squamous cell carcinoma. Int J Cancer. Jan 1 2020;146(1):18-25. doi:10.1002/ijc.32220 |
| Esophageal Male | Servings of vegetables per day (square-root transformation) | Continuous | 0.89 | UK Biobank | 1.22 |  | World Cancer Research Fund/American Institute for Cancer Research. Continuous Update Project Expert Report 2018. Diet, nutrition, physical activity, and oesophageal cancer. 2017. https://www.wcrf.org/wp-content/uploads/2021/02/oesophageal-cancer-report.pdf |
| Esophageal Male | Smoking (current) | Binary | 1.56 | UK Biobank | 0.13 |  | Cook MB, Kamangar F, Whiteman DC, et al. Cigarette smoking and adenocarcinomas of the esophagus and esophagogastric junction: a pooled analysis from the international BEACON consortium. J Natl Cancer Inst. Sep 8 2010;102(17):1344-53. doi:10.1093/jnci/djq289 Iribarren C, Tekawa IS, Sidney S, Friedman GD. Effect of cigar smoking on the risk of cardiovascular disease, chronic obstructive pulmonary disease, and cancer in men. N Engl J Med. Jun 10 1999;340(23):1773-80. doi:10.1056/nejm199906103402301 Randi G, Scotti L, Bosetti C, et al. Pipe smoking and cancers of the upper digestive tract. Int J Cancer. Nov 1 2007;121(9):2049-2051. doi:10.1002/ijc.22791 Freedman ND, Abnet CC, Caporaso NE, et al. Impact of changing US cigarette smoking patterns on incident cancer: risks of 20 smoking-related cancers among the women and men of the NIH-AARP cohort. Int J Epidemiol. Jun 2016;45(3):846-56. doi:10.1093/ije/dyv175 |
| Head_And_Neck Female | Alcohol consumption (2-3 drinks/day) | Binary | 1.73 | Literature | 0.12 | 0.12 | Bagnardi V, Blangiardo M, La Vecchia C, Corrao G. Alcohol consumption and the risk of cancer: a meta-analysis. Alcohol Res Health. 2001;25(4):263-70. https://www.ncbi.nlm.nih.gov/pmc/articles/PMC6705703/ |
| Head_And_Neck Female | Alcohol consumption (4+ drinks/day) | Binary | 2.77 | Literature | 0.05 | 0.05 | Bagnardi V, Blangiardo M, La Vecchia C, Corrao G. Alcohol consumption and the risk of cancer: a meta-analysis. Alcohol Res Health. 2001;25(4):263-70. https://www.ncbi.nlm.nih.gov/pmc/articles/PMC6705703/ |
| Head_And_Neck Female | Alcohol-based mouthwash use > once a day | Binary | 1.31 | Literature | 0.12 | 0.12 | Hashim D, Sartori S, Brennan P, et al. The role of oral hygiene in head and neck cancer: results from International Head and Neck Cancer Epidemiology (INHANCE) consortium. Ann Oncol. Aug 2016;27(8):1619-25. doi:10.1093/annonc/mdw224 |
| Head_And_Neck Female | Chewing tobacco use | Binary | 1.20 | Literature | 0.04 | 0.04 | Li X, Koskinen AI, Hemminki O, Försti A, Sundquist J, Sundquist K, Hemminki K. Family History of Head and Neck Cancers. Cancers (Basel). Aug 16 2021;13(16)doi:10.3390/cancers13164115 |
| Head_And_Neck Female | Chronic hepatitis B | Binary | 5.10 | UK Biobank | 0.00 |  | Tan R, Zhu X, Sun Y, et al. The association of HBV infection and head and neck cancer: a systematic review and meta-analysis. BMC Cancer. Feb 16 2024;24(1):225. doi:10.1186/s12885-024-11967-7 |
| Head_And_Neck Female | Chronic hepatitis C | Binary | 2.70 | UK Biobank | 0.00 |  | Mahale P, Sturgis EM, Tweardy DJ, Ariza-Heredia EJ, Torres HA. Association Between Hepatitis C Virus and Head and Neck Cancers. J Natl Cancer Inst. Aug 2016;108(8)doi:10.1093/jnci/djw035 |
| Head_And_Neck Female | Chronic kidney disease | Binary | 1.30 | UK Biobank | 0.04 |  | Stengel B. Chronic kidney disease and cancer: a troubling connection. J Nephrol. May-Jun 2010;23(3):253-62. |
| Head_And_Neck Female | Cups of coffee daily (fifth-root transformation) | Continuous | 0.92 | UK Biobank | 0.85 |  | World Cancer Research Fund/American Institute for Cancer Research. Continuous Update Project Expert Report 2018. Diet, nutrition, physical activity, and cancers of the mouth, pharynx, and larynx. 2017. https://www.wcrf.org/wp-content/uploads/2021/02/mouth-pharynx-larynx-cancer-report.pdf |
| Head_And_Neck Female | Daily toothbrushing | Binary | 0.83 | Literature | 0.98 | 0.98 | Hashim D, Sartori S, Brennan P, et al. The role of oral hygiene in head and neck cancer: results from International Head and Neck Cancer Epidemiology (INHANCE) consortium. Ann Oncol. Aug 2016;27(8):1619-25. doi:10.1093/annonc/mdw224 |
| Head_And_Neck Female | Dentist visit at least annually | Binary | 0.82 | Literature | 0.65 | 0.65 | Hashim D, Sartori S, Brennan P, et al. The role of oral hygiene in head and neck cancer: results from International Head and Neck Cancer Epidemiology (INHANCE) consortium. Ann Oncol. Aug 2016;27(8):1619-25. doi:10.1093/annonc/mdw224 |
| Head_And_Neck Female | Family history of head and neck cancer in first degree relative | Binary | 1.78 | Literature | 0.03 | 0.03 | Li X, Koskinen AI, Hemminki O, Försti A, Sundquist J, Sundquist K, Hemminki K. Family History of Head and Neck Cancers. Cancers (Basel). Aug 16 2021;13(16)doi:10.3390/cancers13164115 |
| Head_And_Neck Female | Human papilloma virus vaccine before age 17 | Binary | 0.95 | Literature | 0.17 | 0.17 | US Food and Drug Administration Label for Human Papilloma 9-valent Vaccine. https://www.fda.gov/media/90064/download |
| Head_And_Neck Female | Human papilloma virus vaccine before between age 17-31 | Binary | 0.95 | Literature | 0.34 | 0.34 | US Food and Drug Administration Label for Human Papilloma 9-valent Vaccine. https://www.fda.gov/media/90064/download |
| Head_And_Neck Female | No gingivitis | Binary | 0.94 | Literature | 0.58 | 0.58 | Hashim D, Sartori S, Brennan P, et al. The role of oral hygiene in head and neck cancer: results from International Head and Neck Cancer Epidemiology (INHANCE) consortium. Ann Oncol. Aug 2016;27(8):1619-25. doi:10.1093/annonc/mdw224 |
| Head_And_Neck Female | Pack-years smoking history (fifth-root transformation) | Continuous | 1.46 | UK Biobank | 0.00 |  | Wyss A, Hashibe M, Chuang SC, et al. Cigarette, cigar, and pipe smoking and the risk of head and neck cancers: pooled analysis in the International Head and Neck Cancer Epidemiology Consortium. Am J Epidemiol. Sep 1 2013;178(5):679-90. doi:10.1093/aje/kwt029 |
| Head_And_Neck Female | Servings of processed meat (bacon, ham, sausages, burgers, nuggets, etc.) per week (square-root transformation) | Continuous | 1.11 | UK Biobank | 1.64 |  | Morales-Berstein F, Biessy C, Viallon V, et al. Ultra-processed foods, adiposity and risk of head and neck cancer and oesophageal adenocarcinoma in the European Prospective Investigation into Cancer and Nutrition study: a mediation analysis. European Journal of Nutrition. 2024/03/01 2024;63(2):377-396. doi:10.1007/s00394-023-03270-1 |
| Head_And_Neck Female | Servings of vegetables per day (square-root transformation) | Continuous | 0.94 | UK Biobank | 1.22 |  | Freedman ND, Park Y, Subar AF, Hollenbeck AR, Leitzmann MF, Schatzkin A, Abnet CC. Fruit and vegetable intake and head and neck cancer risk in a large United States prospective cohort study. Int J Cancer. May 15 2008;122(10):2330-6. doi:10.1002/ijc.23319 Boeing H, Dietrich T, Hoffmann K, et al. Intake of fruits and vegetables and risk of cancer of the upper aero-digestive tract: the prospective EPIC-study. Cancer Causes Control. Sep 2006;17(7):957-69. doi:10.1007/s10552-006-0036-4 |
| Head_And_Neck Female | Smoking (current) | Binary | 2.37 | UK Biobank | 0.09 |  | Wyss A, Hashibe M, Chuang SC, et al. Cigarette, cigar, and pipe smoking and the risk of head and neck cancers: pooled analysis in the International Head and Neck Cancer Epidemiology Consortium. Am J Epidemiol. Sep 1 2013;178(5):679-90. doi:10.1093/aje/kwt029 |
| Head_And_Neck Female | Snuff use | Binary | 1.71 | Literature | 0.07 | 0.07 | Wyss AB, Hashibe M, Lee YA, et al. Smokeless Tobacco Use and the Risk of Head and Neck Cancer: Pooled Analysis of US Studies in the INHANCE Consortium. Am J Epidemiol. Nov 15 2016;184(10):703-716. doi:10.1093/aje/kww075 |
| Head_And_Neck Male | Alcohol consumption (2-3 drinks/day) | Binary | 1.73 | Literature | 0.12 | 0.12 | Bagnardi V, Blangiardo M, La Vecchia C, Corrao G. Alcohol consumption and the risk of cancer: a meta-analysis. Alcohol Res Health. 2001;25(4):263-70. https://www.ncbi.nlm.nih.gov/pmc/articles/PMC6705703/ |
| Head_And_Neck Male | Alcohol consumption (4+ drinks/day) | Binary | 2.77 | Literature | 0.05 | 0.05 | Bagnardi V, Blangiardo M, La Vecchia C, Corrao G. Alcohol consumption and the risk of cancer: a meta-analysis. Alcohol Res Health. 2001;25(4):263-70. https://www.ncbi.nlm.nih.gov/pmc/articles/PMC6705703/ |
| Head_And_Neck Male | Alcohol-based mouthwash use > once a day | Binary | 1.31 | Literature | 0.12 | 0.12 | Hashim D, Sartori S, Brennan P, et al. The role of oral hygiene in head and neck cancer: results from International Head and Neck Cancer Epidemiology (INHANCE) consortium. Ann Oncol. Aug 2016;27(8):1619-25. doi:10.1093/annonc/mdw224 |
| Head_And_Neck Male | Chewing tobacco use | Binary | 1.20 | Literature | 0.04 | 0.04 | Li X, Koskinen AI, Hemminki O, Försti A, Sundquist J, Sundquist K, Hemminki K. Family History of Head and Neck Cancers. Cancers (Basel). Aug 16 2021;13(16)doi:10.3390/cancers13164115 |
| Head_And_Neck Male | Chronic hepatitis B | Binary | 1.20 | UK Biobank | 0.00 |  | Tan R, Zhu X, Sun Y, et al. The association of HBV infection and head and neck cancer: a systematic review and meta-analysis. BMC Cancer. Feb 16 2024;24(1):225. doi:10.1186/s12885-024-11967-7 |
| Head_And_Neck Male | Chronic hepatitis C | Binary | 3.16 | UK Biobank | 0.00 |  | Mahale P, Sturgis EM, Tweardy DJ, Ariza-Heredia EJ, Torres HA. Association Between Hepatitis C Virus and Head and Neck Cancers. J Natl Cancer Inst. Aug 2016;108(8)doi:10.1093/jnci/djw035 |
| Head_And_Neck Male | Chronic kidney disease | Binary | 1.33 | UK Biobank | 0.05 |  | Stengel B. Chronic kidney disease and cancer: a troubling connection. J Nephrol. May-Jun 2010;23(3):253-62. |
| Head_And_Neck Male | Cups of coffee daily (fifth-root transformation) | Continuous | 0.84 | UK Biobank | 0.90 |  | World Cancer Research Fund/American Institute for Cancer Research. Continuous Update Project Expert Report 2018. Diet, nutrition, physical activity, and cancers of the mouth, pharynx, and larynx. 2017. https://www.wcrf.org/wp-content/uploads/2021/02/mouth-pharynx-larynx-cancer-report.pdf |
| Head_And_Neck Male | Daily toothbrushing | Binary | 0.83 | Literature | 0.98 | 0.98 | Hashim D, Sartori S, Brennan P, et al. The role of oral hygiene in head and neck cancer: results from International Head and Neck Cancer Epidemiology (INHANCE) consortium. Ann Oncol. Aug 2016;27(8):1619-25. doi:10.1093/annonc/mdw224 |
| Head_And_Neck Male | Dentist visit at least annually | Binary | 0.82 | Literature | 0.65 | 0.65 | Hashim D, Sartori S, Brennan P, et al. The role of oral hygiene in head and neck cancer: results from International Head and Neck Cancer Epidemiology (INHANCE) consortium. Ann Oncol. Aug 2016;27(8):1619-25. doi:10.1093/annonc/mdw224 |
| Head_And_Neck Male | Family history of head and neck cancer in first degree relative | Binary | 1.78 | Literature | 0.03 | 0.03 | Li X, Koskinen AI, Hemminki O, Försti A, Sundquist J, Sundquist K, Hemminki K. Family History of Head and Neck Cancers. Cancers (Basel). Aug 16 2021;13(16)doi:10.3390/cancers13164115 |
| Head_And_Neck Male | Human papilloma virus vaccine before age 17 | Binary | 0.83 | Literature | 0.06 | 0.06 | US Food and Drug Administration Label for Human Papilloma 9-valent Vaccine. https://www.fda.gov/media/90064/download |
| Head_And_Neck Male | Human papilloma virus vaccine before between age 17-32 | Binary | 0.83 | Literature | 0.11 | 0.11 | US Food and Drug Administration Label for Human Papilloma 9-valent Vaccine. https://www.fda.gov/media/90064/download |
| Head_And_Neck Male | Multivitamin use | Binary | 1.09 | Literature | 0.04 | 0.50 | Lim J-e, Weinstein SJ, Liao LM, Sinha R, Huang J, Albanes D. Multivitamin Use and Overall and Site-Specific Cancer Risks in the National Institutes of Health–AARP Diet and Health Study. The Journal of Nutrition. 2022/01/01/ 2022;152(1):211-216. doi:https://doi.org/10.1093/jn/nxab322 |
| Head_And_Neck Male | No gingivitis | Binary | 0.94 | Literature | 0.58 | 0.58 | Hashim D, Sartori S, Brennan P, et al. The role of oral hygiene in head and neck cancer: results from International Head and Neck Cancer Epidemiology (INHANCE) consortium. Ann Oncol. Aug 2016;27(8):1619-25. doi:10.1093/annonc/mdw224 |
| Head_And_Neck Male | Pack-years smoking history (fifth-root transformation) | Continuous | 1.38 | UK Biobank | 0.00 |  | Wyss A, Hashibe M, Chuang SC, et al. Cigarette, cigar, and pipe smoking and the risk of head and neck cancers: pooled analysis in the International Head and Neck Cancer Epidemiology Consortium. Am J Epidemiol. Sep 1 2013;178(5):679-90. doi:10.1093/aje/kwt029 |
| Head_And_Neck Male | Servings of fruit per day (square-root transformation | Continuous | 0.87 | UK Biobank | 1.00 |  | Freedman ND, Park Y, Subar AF, Hollenbeck AR, Leitzmann MF, Schatzkin A, Abnet CC. Fruit and vegetable intake and head and neck cancer risk in a large United States prospective cohort study. Int J Cancer. May 15 2008;122(10):2330-6. doi:10.1002/ijc.23319 Boeing H, Dietrich T, Hoffmann K, et al. Intake of fruits and vegetables and risk of cancer of the upper aero-digestive tract: the prospective EPIC-study. Cancer Causes Control. Sep 2006;17(7):957-69. doi:10.1007/s10552-006-0036-4 |
| Head_And_Neck Male | Servings of processed meat (bacon, ham, sausages, burgers, nuggets, etc.) per week (square-root transformation) | Continuous | 1.07 | UK Biobank | 1.64 |  | Morales-Berstein F, Biessy C, Viallon V, et al. Ultra-processed foods, adiposity and risk of head and neck cancer and oesophageal adenocarcinoma in the European Prospective Investigation into Cancer and Nutrition study: a mediation analysis. European Journal of Nutrition. 2024/03/01 2024;63(2):377-396. doi:10.1007/s00394-023-03270-1 |
| Head_And_Neck Male | Servings of vegetables per day (square-root transformation) | Continuous | 0.97 | UK Biobank | 1.22 |  | Freedman ND, Park Y, Subar AF, Hollenbeck AR, Leitzmann MF, Schatzkin A, Abnet CC. Fruit and vegetable intake and head and neck cancer risk in a large United States prospective cohort study. Int J Cancer. May 15 2008;122(10):2330-6. doi:10.1002/ijc.23319 Boeing H, Dietrich T, Hoffmann K, et al. Intake of fruits and vegetables and risk of cancer of the upper aero-digestive tract: the prospective EPIC-study. Cancer Causes Control. Sep 2006;17(7):957-69. doi:10.1007/s10552-006-0036-4 |
| Head_And_Neck Male | Smoking (current) | Binary | 2.19 | UK Biobank | 0.13 |  | Wyss A, Hashibe M, Chuang SC, et al. Cigarette, cigar, and pipe smoking and the risk of head and neck cancers: pooled analysis in the International Head and Neck Cancer Epidemiology Consortium. Am J Epidemiol. Sep 1 2013;178(5):679-90. doi:10.1093/aje/kwt029 |
| Head_And_Neck Male | Snuff use | Binary | 1.71 | Literature | 0.07 | 0.07 | Wyss AB, Hashibe M, Lee YA, et al. Smokeless Tobacco Use and the Risk of Head and Neck Cancer: Pooled Analysis of US Studies in the INHANCE Consortium. Am J Epidemiol. Nov 15 2016;184(10):703-716. doi:10.1093/aje/kww075 |
| Kidney Female | Alcohol consumption (0-1 drinks/day) | Binary | 0.90 | Literature | 0.48 | 0.48 | Bellocco R, Pasquali E, Rota M, et al. Alcohol drinking and risk of renal cell carcinoma: results of a meta-analysis. Ann Oncol. Sep 2012;23(9):2235-2244. doi:10.1093/annonc/mds022 |
| Kidney Female | Alcohol consumption (2-3 drinks/day) | Binary | 0.79 | Literature | 0.12 | 0.12 | Bagnardi V, Blangiardo M, La Vecchia C, Corrao G. Alcohol consumption and the risk of cancer: a meta-analysis. Alcohol Res Health. 2001;25(4):263-70. https://www.ncbi.nlm.nih.gov/pmc/articles/PMC6705703/ |
| Kidney Female | Body mass index | Continuous | 1.05 | UK Biobank | 29.80 |  | Pischon T, Lahmann PH, Boeing H, et al. Body size and risk of renal cell carcinoma in the European Prospective Investigation into Cancer and Nutrition (EPIC). Int J Cancer. Feb 1 2006;118(3):728-38. doi:10.1002/ijc.21398 Adams KF, Leitzmann MF, Albanes D, Kipnis V, Moore SC, Schatzkin A, Chow WH. Body size and renal cell cancer incidence in a large US cohort study. Am J Epidemiol. Aug 1 2008;168(3):268-77. doi:10.1093/aje/kwn122 |
| Kidney Female | Chronic hepatitis C | Binary | 2.76 | UK Biobank | 0.00 |  | Gordon SC, Moonka D, Brown KA, Rogers C, Huang MA, Bhatt N, Lamerato L. Risk for renal cell carcinoma in chronic hepatitis C infection. Cancer Epidemiol Biomarkers Prev. Apr 2010;19(4):1066-73. doi:10.1158/1055-9965.Epi-09-1275 |
| Kidney Female | Chronic kidney disease | Binary | 3.30 | UK Biobank | 0.04 |  | Lowrance WT, Ordoñez J, Udaltsova N, Russo P, Go AS. CKD and the risk of incident cancer. J Am Soc Nephrol. Oct 2014;25(10):2327-34. doi:10.1681/asn.2013060604 |
| Kidney Female | End-stage renal disease on dialysis | Binary | 2.01 | UK Biobank | 0.00 |  | Truong LD, Krishnan B, Cao JT, Barrios R, Suki WN. Renal neoplasm in acquired cystic kidney disease. Am J Kidney Dis. Jul 1995;26(1):1-12. doi:10.1016/0272-6386(95)90146-9 |
| Kidney Female | Hypertension | Binary | 1.53 | UK Biobank | 0.51 |  | Hidayat K, Du X, Zou SY, Shi BM. Blood pressure and kidney cancer risk: meta-analysis of prospective studies. J Hypertens. Jul 2017;35(7):1333-1344. doi:10.1097/hjh.0000000000001286 |
| Kidney Female | Kidney stones | Binary | 1.98 | UK Biobank | 0.01 |  | Cheungpasitporn W, Thongprayoon C, O'Corragain OA, Edmonds PJ, Ungprasert P, Kittanamongkolchai W, Erickson SB. The risk of kidney cancer in patients with kidney stones: a systematic review and meta-analysis. Qjm. Mar 2015;108(3):205-12. doi:10.1093/qjmed/hcu195 |
| Kidney Female | Smoking (current) | Binary | 1.78 | UK Biobank | 0.09 |  | Cumberbatch MG, Rota M, Catto JW, La Vecchia C. The Role of Tobacco Smoke in Bladder and Kidney Carcinogenesis: A Comparison of Exposures and Meta-analysis of Incidence and Mortality Risks. Eur Urol. Sep 2016;70(3):458-66. doi:10.1016/j.eururo.2015.06.042 |
| Kidney Male | Alcohol consumption (0-1 drinks/day) | Binary | 0.90 | Literature | 0.48 | 0.48 | Bellocco R, Pasquali E, Rota M, et al. Alcohol drinking and risk of renal cell carcinoma: results of a meta-analysis. Ann Oncol. Sep 2012;23(9):2235-2244. doi:10.1093/annonc/mds022 |
| Kidney Male | Alcohol consumption (2-3 drinks/day) | Binary | 0.79 | Literature | 0.12 | 0.12 | Bagnardi V, Blangiardo M, La Vecchia C, Corrao G. Alcohol consumption and the risk of cancer: a meta-analysis. Alcohol Res Health. 2001;25(4):263-70. https://www.ncbi.nlm.nih.gov/pmc/articles/PMC6705703/ |
| Kidney Male | Body mass index | Continuous | 1.04 | UK Biobank | 29.40 |  | Pischon T, Lahmann PH, Boeing H, et al. Body size and risk of renal cell carcinoma in the European Prospective Investigation into Cancer and Nutrition (EPIC). Int J Cancer. Feb 1 2006;118(3):728-38. doi:10.1002/ijc.21398 Adams KF, Leitzmann MF, Albanes D, Kipnis V, Moore SC, Schatzkin A, Chow WH. Body size and renal cell cancer incidence in a large US cohort study. Am J Epidemiol. Aug 1 2008;168(3):268-77. doi:10.1093/aje/kwn122 |
| Kidney Male | Chronic kidney disease | Binary | 3.72 | UK Biobank | 0.05 |  | Lowrance WT, Ordoñez J, Udaltsova N, Russo P, Go AS. CKD and the risk of incident cancer. J Am Soc Nephrol. Oct 2014;25(10):2327-34. doi:10.1681/asn.2013060604 |
| Kidney Male | End-stage renal disease on dialysis | Binary | 1.57 | UK Biobank | 0.00 |  | Truong LD, Krishnan B, Cao JT, Barrios R, Suki WN. Renal neoplasm in acquired cystic kidney disease. Am J Kidney Dis. Jul 1995;26(1):1-12. doi:10.1016/0272-6386(95)90146-9 |
| Kidney Male | Hypertension | Binary | 1.08 | UK Biobank | 0.51 |  | Hidayat K, Du X, Zou SY, Shi BM. Blood pressure and kidney cancer risk: meta-analysis of prospective studies. J Hypertens. Jul 2017;35(7):1333-1344. doi:10.1097/hjh.0000000000001286 |
| Kidney Male | Kidney stones | Binary | 1.99 | UK Biobank | 0.03 |  | Cheungpasitporn W, Thongprayoon C, O'Corragain OA, Edmonds PJ, Ungprasert P, Kittanamongkolchai W, Erickson SB. The risk of kidney cancer in patients with kidney stones: a systematic review and meta-analysis. Qjm. Mar 2015;108(3):205-12. doi:10.1093/qjmed/hcu195 |
| Kidney Male | Pack-years smoking history (fifth-root transformation) | Continuous | 1.07 | UK Biobank | 0.00 |  | Cumberbatch MG, Rota M, Catto JW, La Vecchia C. The Role of Tobacco Smoke in Bladder and Kidney Carcinogenesis: A Comparison of Exposures and Meta-analysis of Incidence and Mortality Risks. Eur Urol. Sep 2016;70(3):458-66. doi:10.1016/j.eururo.2015.06.042 |
| Kidney Male | Smoking (current) | Binary | 1.37 | UK Biobank | 0.13 |  | Cumberbatch MG, Rota M, Catto JW, La Vecchia C. The Role of Tobacco Smoke in Bladder and Kidney Carcinogenesis: A Comparison of Exposures and Meta-analysis of Incidence and Mortality Risks. Eur Urol. Sep 2016;70(3):458-66. doi:10.1016/j.eururo.2015.06.042 |
| Leukemia Female | CT scan of head as a child | Binary | 1.40 | Literature | 0.02 | 0.02 | Pearce MS, Salotti JA, Little MP, et al. Radiation exposure from CT scans in childhood and subsequent risk of leukaemia and brain tumours: a retrospective cohort study. The Lancet. 2012/08/04/ 2012;380(9840):499-505. doi:https://doi.org/10.1016/S0140-6736(12)60815-0 |
| Leukemia Female | Family history of leukemia in first degree relative | Binary | 3.19 | Literature | 0.00 | 0.00 | Sud A, Chattopadhyay S, Thomsen H, Sundquist K, Sundquist J, Houlston RS, Hemminki K. Analysis of 153 115 patients with hematological malignancies refines the spectrum of familial risk. Blood. Sep 19 2019;134(12):960-969. doi:10.1182/blood.2019001362 |
| Leukemia Female | Multivitamin use | Binary | 1.26 | Literature | 0.06 | 0.50 | Lim J-e, Weinstein SJ, Liao LM, Sinha R, Huang J, Albanes D. Multivitamin Use and Overall and Site-Specific Cancer Risks in the National Institutes of Health–AARP Diet and Health Study. The Journal of Nutrition. 2022/01/01/ 2022;152(1):211-216. doi:https://doi.org/10.1093/jn/nxab322 |
| Leukemia Female | Pack-years smoking history (fifth-root transformation) | Continuous | 1.11 | UK Biobank | 0.00 |  | Musselman JR, Blair CK, Cerhan JR, Nguyen P, Hirsch B, Ross JA. Risk of adult acute and chronic myeloid leukemia with cigarette smoking and cessation. Cancer Epidemiol. Aug 2013;37(4):410-6. doi:10.1016/j.canep.2013.03.012 |
| Leukemia Female | Servings of fruit per day (square-root transformation | Continuous | 1.00 | UK Biobank | 1.00 |  | Yamamura Y, Oum R, Gbito KY, Garcia-Manero G, Strom SS. Dietary intake of vegetables, fruits, and meats/beans as potential risk factors of acute myeloid leukemia: a Texas case-control study. Nutr Cancer. 2013;65(8):1132-40. doi:10.1080/01635581.2013.834946 |
| Leukemia Female | Servings of red meat (beef, lamb, or pork) per week (square-root transformation) | Binary | 1.01 | UK Biobank | 2.12 |  | Yamamura Y, Oum R, Gbito KY, Garcia-Manero G, Strom SS. Dietary intake of vegetables, fruits, and meats/beans as potential risk factors of acute myeloid leukemia: a Texas case-control study. Nutr Cancer. 2013;65(8):1132-40. doi:10.1080/01635581.2013.834946 |
| Leukemia Male | CT scan of head as a child | Binary | 1.40 | Literature | 0.02 | 0.02 | Pearce MS, Salotti JA, Little MP, et al. Radiation exposure from CT scans in childhood and subsequent risk of leukaemia and brain tumours: a retrospective cohort study. The Lancet. 2012/08/04/ 2012;380(9840):499-505. doi:https://doi.org/10.1016/S0140-6736(12)60815-0 |
| Leukemia Male | Family history of leukemia in first degree relative | Binary | 3.19 | Literature | 0.00 | 0.00 | Sud A, Chattopadhyay S, Thomsen H, Sundquist K, Sundquist J, Houlston RS, Hemminki K. Analysis of 153 115 patients with hematological malignancies refines the spectrum of familial risk. Blood. Sep 19 2019;134(12):960-969. doi:10.1182/blood.2019001362 |
| Leukemia Male | Multivitamin use | Binary | 1.20 | Literature | 0.04 | 0.50 | Lim J-e, Weinstein SJ, Liao LM, Sinha R, Huang J, Albanes D. Multivitamin Use and Overall and Site-Specific Cancer Risks in the National Institutes of Health–AARP Diet and Health Study. The Journal of Nutrition. 2022/01/01/ 2022;152(1):211-216. doi:https://doi.org/10.1093/jn/nxab322 |
| Leukemia Male | Pack-years smoking history (fifth-root transformation) | Continuous | 1.07 | UK Biobank | 0.00 |  | Musselman JR, Blair CK, Cerhan JR, Nguyen P, Hirsch B, Ross JA. Risk of adult acute and chronic myeloid leukemia with cigarette smoking and cessation. Cancer Epidemiol. Aug 2013;37(4):410-6. doi:10.1016/j.canep.2013.03.012 |
| Leukemia Male | Servings of fruit per day (square-root transformation | Continuous | 0.98 | UK Biobank | 1.00 |  | Yamamura Y, Oum R, Gbito KY, Garcia-Manero G, Strom SS. Dietary intake of vegetables, fruits, and meats/beans as potential risk factors of acute myeloid leukemia: a Texas case-control study. Nutr Cancer. 2013;65(8):1132-40. doi:10.1080/01635581.2013.834946 |
| Leukemia Male | Servings of red meat (beef, lamb, or pork) per week (square-root transformation) | Binary | 1.08 | UK Biobank | 2.12 |  | Yamamura Y, Oum R, Gbito KY, Garcia-Manero G, Strom SS. Dietary intake of vegetables, fruits, and meats/beans as potential risk factors of acute myeloid leukemia: a Texas case-control study. Nutr Cancer. 2013;65(8):1132-40. doi:10.1080/01635581.2013.834946 |
| Leukemia Male | Servings of vegetables per day (square-root transformation) | Continuous | 0.95 | UK Biobank | 1.22 |  | Yamamura Y, Oum R, Gbito KY, Garcia-Manero G, Strom SS. Dietary intake of vegetables, fruits, and meats/beans as potential risk factors of acute myeloid leukemia: a Texas case-control study. Nutr Cancer. 2013;65(8):1132-40. doi:10.1080/01635581.2013.834946 |
| Leukemia Male | Smoking (current) | Binary | 1.24 | UK Biobank | 0.13 |  | Musselman JR, Blair CK, Cerhan JR, Nguyen P, Hirsch B, Ross JA. Risk of adult acute and chronic myeloid leukemia with cigarette smoking and cessation. Cancer Epidemiol. Aug 2013;37(4):410-6. doi:10.1016/j.canep.2013.03.012 |
| Liver Female | ≥ 1 serving daily of sugar-sweetened beverages | Binary | 1.85 | Literature | 0.07 | 0.07 | Zhao L, Zhang X, Coday M, et al. Sugar-Sweetened and Artificially Sweetened Beverages and Risk of Liver Cancer and Chronic Liver Disease Mortality. Jama. Aug 8 2023;330(6):537-546. doi:10.1001/jama.2023.12618 |
| Liver Female | Alcohol consumption (2-3 drinks/day) | Binary | 1.20 | Literature | 0.12 | 0.12 | Bagnardi V, Blangiardo M, La Vecchia C, Corrao G. Alcohol consumption and the risk of cancer: a meta-analysis. Alcohol Res Health. 2001;25(4):263-70. https://www.ncbi.nlm.nih.gov/pmc/articles/PMC6705703/ |
| Liver Female | Alcohol consumption (4+ drinks/day) | Binary | 1.41 | Literature | 0.05 | 0.05 | Bagnardi V, Blangiardo M, La Vecchia C, Corrao G. Alcohol consumption and the risk of cancer: a meta-analysis. Alcohol Res Health. 2001;25(4):263-70. https://www.ncbi.nlm.nih.gov/pmc/articles/PMC6705703/ |
| Liver Female | Aspirin use | Binary | 0.82 | UK Biobank | 0.10 |  | Simon TG, Ma Y, Ludvigsson JF, et al. Association Between Aspirin Use and Risk of Hepatocellular Carcinoma. JAMA Oncol. Dec 1 2018;4(12):1683-1690. doi:10.1001/jamaoncol.2018.4154 Simon TG, Duberg AS, Aleman S, Chung RT, Chan AT, Ludvigsson JF. Association of Aspirin with Hepatocellular Carcinoma and Liver-Related Mortality. N Engl J Med. Mar 12 2020;382(11):1018-1028. doi:10.1056/NEJMoa1912035 |
| Liver Female | Body mass index | Continuous | 1.03 | UK Biobank | 29.80 |  | Larsson SC, Wolk A. Overweight, obesity and risk of liver cancer: a meta-analysis of cohort studies. Br J Cancer. Oct 8 2007;97(7):1005-8. doi:10.1038/sj.bjc.6603932 |
| Liver Female | Chronic hepatitis B | Binary | 1.72 | UK Biobank | 0.00 |  | El-Serag HB. Epidemiology of viral hepatitis and hepatocellular carcinoma. Gastroenterology. May 2012;142(6):1264-1273.e1. doi:10.1053/j.gastro.2011.12.061 |
| Liver Female | Chronic hepatitis C | Binary | 11.20 | UK Biobank | 0.00 |  | El-Serag HB. Epidemiology of viral hepatitis and hepatocellular carcinoma. Gastroenterology. May 2012;142(6):1264-1273.e1. doi:10.1053/j.gastro.2011.12.061 |
| Liver Female | Cirrhosis | Binary | 14.94 | UK Biobank | 0.00 |  | Sangiovanni A, Prati GM, Fasani P, et al. The natural history of compensated cirrhosis due to hepatitis C virus: A 17-year cohort study of 214 patients. Hepatology. Jun 2006;43(6):1303-10. doi:10.1002/hep.21176 |
| Liver Female | Cups of coffee daily (fifth-root transformation) | Continuous | 0.93 | UK Biobank | 0.85 |  | Bravi F, Tavani A, Bosetti C, Boffetta P, La Vecchia C. Coffee and the risk of hepatocellular carcinoma and chronic liver disease: a systematic review and meta-analysis of prospective studies. Eur J Cancer Prev. Sep 2017;26(5):368-377. doi:10.1097/cej.0000000000000252 |
| Liver Female | Diabetes mellitus type 2 | Binary | 1.44 | UK Biobank | 0.04 |  | Wang C, Wang X, Gong G, et al. Increased risk of hepatocellular carcinoma in patients with diabetes mellitus: a systematic review and meta-analysis of cohort studies. Int J Cancer. Apr 1 2012;130(7):1639-48. doi:10.1002/ijc.26165 Yang WS, Va P, Bray F, Gao S, Gao J, Li HL, Xiang YB. The role of pre-existing diabetes mellitus on hepatocellular carcinoma occurrence and prognosis: a meta-analysis of prospective cohort studies. PLoS One. 2011;6(12):e27326. doi:10.1371/journal.pone.0027326 |
| Liver Female | Family history of liver cancer in first degree relative | Binary | 4.10 | Literature | 0.03 | 0.03 | Hassan MM, Spitz MR, Thomas MB, et al. The association of family history of liver cancer with hepatocellular carcinoma: a case-control study in the United States. J Hepatol. Feb 2009;50(2):334-41. doi:10.1016/j.jhep.2008.08.016 |
| Liver Female | Hereditary hemochromatosis | Binary | 21.00 | Literature | 0.00 | 0.00 | Elmberg M, Hultcrantz R, Ekbom A, et al. Cancer risk in patients with hereditary hemochromatosis and in their first-degree relatives. Gastroenterology. Dec 2003;125(6):1733-41. doi:10.1053/j.gastro.2003.09.035 |
| Liver Female | Minutes of moderate activity daily (<30 minutes, 30-59 minutes, 60-89 minutes, >90 minutes) | Ordinal | 0.98 | UK Biobank | 1.00 |  | Baumeister SE, Schlesinger S, Aleksandrova K, et al. Association between physical activity and risk of hepatobiliary cancers: A multinational cohort study. J Hepatol. May 2019;70(5):885-892. doi:10.1016/j.jhep.2018.12.014 |
| Liver Female | Nonalcoholic fatty liver disease | Binary | 2.24 | UK Biobank | 0.01 |  | Mittal S, El-Serag HB, Sada YH, et al. Hepatocellular Carcinoma in the Absence of Cirrhosis in United States Veterans is Associated With Nonalcoholic Fatty Liver Disease. Clin Gastroenterol Hepatol. Jan 2016;14(1):124-31.e1. doi:10.1016/j.cgh.2015.07.019 |
| Liver Female | Nonsteroidal anti-inflammatory drug use | Binary | 0.77 | UK Biobank | 0.17 |  | Tan RZH, Lockart I, Abdel Shaheed C, Danta M. Systematic review with meta-analysis: The effects of non-steroidal anti-inflammatory drugs and anti-platelet therapy on the incidence and recurrence of hepatocellular carcinoma. Aliment Pharmacol Ther. Aug 2021;54(4):356-367. doi:10.1111/apt.16515 |
| Liver Female | Pack-years smoking history (fifth-root transformation) | Continuous | 1.20 | UK Biobank | 0.00 |  | Trichopoulos D, Bamia C, Lagiou P, et al. Hepatocellular carcinoma risk factors and disease burden in a European cohort: a nested case-control study. J Natl Cancer Inst. Nov 16 2011;103(22):1686-95. doi:10.1093/jnci/djr395 Lee YC, Cohet C, Yang YC, Stayner L, Hashibe M, Straif K. Meta-analysis of epidemiologic studies on cigarette smoking and liver cancer. Int J Epidemiol. Dec 2009;38(6):1497-511. doi:10.1093/ije/dyp280 |
| Liver Female | Servings of fatty fish per week (square-root transformation) | Continuous | 0.92 | UK Biobank | 1.19 |  | Koumbi L. Dietary factors can protect against liver cancer development. World J Hepatol. Jan 28 2017;9(3):119-125. doi:10.4254/wjh.v9.i3.119 Freedman ND, Cross AJ, McGlynn KA, et al. Association of meat and fat intake with liver disease and hepatocellular carcinoma in the NIH-AARP cohort. J Natl Cancer Inst. Sep 8 2010;102(17):1354-65. doi:10.1093/jnci/djq301 Fedirko V, Trichopolou A, Bamia C, et al. Consumption of fish and meats and risk of hepatocellular carcinoma: the European Prospective Investigation into Cancer and Nutrition (EPIC). Ann Oncol. Aug 2013;24(8):2166-73. doi:10.1093/annonc/mdt168 |
| Liver Female | Smoking (current) | Binary | 1.14 | UK Biobank | 0.09 |  | Trichopoulos D, Bamia C, Lagiou P, et al. Hepatocellular carcinoma risk factors and disease burden in a European cohort: a nested case-control study. J Natl Cancer Inst. Nov 16 2011;103(22):1686-95. doi:10.1093/jnci/djr395 Lee YC, Cohet C, Yang YC, Stayner L, Hashibe M, Straif K. Meta-analysis of epidemiologic studies on cigarette smoking and liver cancer. Int J Epidemiol. Dec 2009;38(6):1497-511. doi:10.1093/ije/dyp280 |
| Liver Female | Statin use | Binary | 0.83 | UK Biobank | 0.13 |  | Singh S, Singh PP, Singh AG, Murad MH, Sanchez W. Statins are associated with a reduced risk of hepatocellular cancer: a systematic review and meta-analysis. Gastroenterology. Feb 2013;144(2):323-332. doi:10.1053/j.gastro.2012.10.005 |
| Liver Male | ≥ 1 serving daily of sugar-sweetened beverages | Binary | 1.85 | Literature | 0.07 | 0.07 | Zhao L, Zhang X, Coday M, et al. Sugar-Sweetened and Artificially Sweetened Beverages and Risk of Liver Cancer and Chronic Liver Disease Mortality. Jama. Aug 8 2023;330(6):537-546. doi:10.1001/jama.2023.12618 |
| Liver Male | Alcohol consumption (2-3 drinks/day) | Binary | 1.20 | Literature | 0.12 | 0.12 | Bagnardi V, Blangiardo M, La Vecchia C, Corrao G. Alcohol consumption and the risk of cancer: a meta-analysis. Alcohol Res Health. 2001;25(4):263-70. https://www.ncbi.nlm.nih.gov/pmc/articles/PMC6705703/ |
| Liver Male | Alcohol consumption (4+ drinks/day) | Binary | 1.41 | Literature | 0.05 | 0.05 | Bagnardi V, Blangiardo M, La Vecchia C, Corrao G. Alcohol consumption and the risk of cancer: a meta-analysis. Alcohol Res Health. 2001;25(4):263-70. https://www.ncbi.nlm.nih.gov/pmc/articles/PMC6705703/ |
| Liver Male | Body mass index | Continuous | 1.03 | UK Biobank | 29.40 |  | Larsson SC, Wolk A. Overweight, obesity and risk of liver cancer: a meta-analysis of cohort studies. Br J Cancer. Oct 8 2007;97(7):1005-8. doi:10.1038/sj.bjc.6603932 |
| Liver Male | Chronic hepatitis B | Binary | 2.35 | UK Biobank | 0.00 |  | El-Serag HB. Epidemiology of viral hepatitis and hepatocellular carcinoma. Gastroenterology. May 2012;142(6):1264-1273.e1. doi:10.1053/j.gastro.2011.12.061 |
| Liver Male | Chronic hepatitis C | Binary | 5.18 | UK Biobank | 0.00 |  | El-Serag HB. Epidemiology of viral hepatitis and hepatocellular carcinoma. Gastroenterology. May 2012;142(6):1264-1273.e1. doi:10.1053/j.gastro.2011.12.061 |
| Liver Male | Cirrhosis | Binary | 46.24 | UK Biobank | 0.01 |  | Sangiovanni A, Prati GM, Fasani P, et al. The natural history of compensated cirrhosis due to hepatitis C virus: A 17-year cohort study of 214 patients. Hepatology. Jun 2006;43(6):1303-10. doi:10.1002/hep.21176 |
| Liver Male | Cups of coffee daily (fifth-root transformation) | Continuous | 1.00 | UK Biobank | 0.90 |  | Bravi F, Tavani A, Bosetti C, Boffetta P, La Vecchia C. Coffee and the risk of hepatocellular carcinoma and chronic liver disease: a systematic review and meta-analysis of prospective studies. Eur J Cancer Prev. Sep 2017;26(5):368-377. doi:10.1097/cej.0000000000000252 |
| Liver Male | Diabetes mellitus type 2 | Binary | 2.19 | UK Biobank | 0.08 |  | Wang C, Wang X, Gong G, et al. Increased risk of hepatocellular carcinoma in patients with diabetes mellitus: a systematic review and meta-analysis of cohort studies. Int J Cancer. Apr 1 2012;130(7):1639-48. doi:10.1002/ijc.26165 Yang WS, Va P, Bray F, Gao S, Gao J, Li HL, Xiang YB. The role of pre-existing diabetes mellitus on hepatocellular carcinoma occurrence and prognosis: a meta-analysis of prospective cohort studies. PLoS One. 2011;6(12):e27326. doi:10.1371/journal.pone.0027326 |
| Liver Male | Family history of liver cancer in first degree relative | Binary | 4.10 | Literature | 0.03 | 0.03 | Hassan MM, Spitz MR, Thomas MB, et al. The association of family history of liver cancer with hepatocellular carcinoma: a case-control study in the United States. J Hepatol. Feb 2009;50(2):334-41. doi:10.1016/j.jhep.2008.08.016 |
| Liver Male | Hereditary hemochromatosis | Binary | 21.00 | Literature | 0.00 | 0.00 | Elmberg M, Hultcrantz R, Ekbom A, et al. Cancer risk in patients with hereditary hemochromatosis and in their first-degree relatives. Gastroenterology. Dec 2003;125(6):1733-41. doi:10.1053/j.gastro.2003.09.035 |
| Liver Male | Minutes of moderate activity daily (<30 minutes, 30-59 minutes, 60-89 minutes, >90 minutes) | Ordinal | 0.95 | UK Biobank | 1.00 |  | Baumeister SE, Schlesinger S, Aleksandrova K, et al. Association between physical activity and risk of hepatobiliary cancers: A multinational cohort study. J Hepatol. May 2019;70(5):885-892. doi:10.1016/j.jhep.2018.12.014 |
| Liver Male | Nonalcoholic fatty liver disease | Binary | 1.82 | UK Biobank | 0.02 |  | Mittal S, El-Serag HB, Sada YH, et al. Hepatocellular Carcinoma in the Absence of Cirrhosis in United States Veterans is Associated With Nonalcoholic Fatty Liver Disease. Clin Gastroenterol Hepatol. Jan 2016;14(1):124-31.e1. doi:10.1016/j.cgh.2015.07.019 |
| Liver Male | Nonsteroidal anti-inflammatory drug use | Binary | 0.98 | UK Biobank | 0.12 |  | Tan RZH, Lockart I, Abdel Shaheed C, Danta M. Systematic review with meta-analysis: The effects of non-steroidal anti-inflammatory drugs and anti-platelet therapy on the incidence and recurrence of hepatocellular carcinoma. Aliment Pharmacol Ther. Aug 2021;54(4):356-367. doi:10.1111/apt.16515 |
| Liver Male | Pack-years smoking history (fifth-root transformation) | Continuous | 1.07 | UK Biobank | 0.00 |  | Trichopoulos D, Bamia C, Lagiou P, et al. Hepatocellular carcinoma risk factors and disease burden in a European cohort: a nested case-control study. J Natl Cancer Inst. Nov 16 2011;103(22):1686-95. doi:10.1093/jnci/djr395 Lee YC, Cohet C, Yang YC, Stayner L, Hashibe M, Straif K. Meta-analysis of epidemiologic studies on cigarette smoking and liver cancer. Int J Epidemiol. Dec 2009;38(6):1497-511. doi:10.1093/ije/dyp280 |
| Liver Male | Servings of fatty fish per week (square-root transformation) | Continuous | 0.89 | UK Biobank | 1.16 |  | Koumbi L. Dietary factors can protect against liver cancer development. World J Hepatol. Jan 28 2017;9(3):119-125. doi:10.4254/wjh.v9.i3.119 Freedman ND, Cross AJ, McGlynn KA, et al. Association of meat and fat intake with liver disease and hepatocellular carcinoma in the NIH-AARP cohort. J Natl Cancer Inst. Sep 8 2010;102(17):1354-65. doi:10.1093/jnci/djq301 Fedirko V, Trichopolou A, Bamia C, et al. Consumption of fish and meats and risk of hepatocellular carcinoma: the European Prospective Investigation into Cancer and Nutrition (EPIC). Ann Oncol. Aug 2013;24(8):2166-73. doi:10.1093/annonc/mdt168 |
| Liver Male | Servings of fruit per day (square-root transformation | Continuous | 0.94 | UK Biobank | 1.00 |  | Koumbi L. Dietary factors can protect against liver cancer development. World J Hepatol. Jan 28 2017;9(3):119-125. doi:10.4254/wjh.v9.i3.119 |
| Liver Male | Servings of vegetables per day (square-root transformation) | Continuous | 0.88 | UK Biobank | 1.22 |  | Yang Y, Zhang D, Feng N, Chen G, Liu J, Chen G, Zhu Y. Increased intake of vegetables, but not fruit, reduces risk for hepatocellular carcinoma: a meta-analysis. Gastroenterology. Nov 2014;147(5):1031-42. doi:10.1053/j.gastro.2014.08.005 |
| Liver Male | Smoking (current) | Binary | 1.30 | UK Biobank | 0.13 |  | Trichopoulos D, Bamia C, Lagiou P, et al. Hepatocellular carcinoma risk factors and disease burden in a European cohort: a nested case-control study. J Natl Cancer Inst. Nov 16 2011;103(22):1686-95. doi:10.1093/jnci/djr395 Lee YC, Cohet C, Yang YC, Stayner L, Hashibe M, Straif K. Meta-analysis of epidemiologic studies on cigarette smoking and liver cancer. Int J Epidemiol. Dec 2009;38(6):1497-511. doi:10.1093/ije/dyp280 |
| Liver Male | Statin use | Binary | 0.69 | UK Biobank | 0.23 |  | Singh S, Singh PP, Singh AG, Murad MH, Sanchez W. Statins are associated with a reduced risk of hepatocellular cancer: a systematic review and meta-analysis. Gastroenterology. Feb 2013;144(2):323-332. doi:10.1053/j.gastro.2012.10.005 |
| Lung Female Nonsmoker | Alcohol consumption (2-3 drinks/day) | Binary | 1.02 | Literature | 0.12 | 0.12 | Bagnardi V, Blangiardo M, La Vecchia C, Corrao G. Alcohol consumption and the risk of cancer: a meta-analysis. Alcohol Res Health. 2001;25(4):263-70. https://www.ncbi.nlm.nih.gov/pmc/articles/PMC6705703/ |
| Lung Female Nonsmoker | Alcohol consumption (4+ drinks/day) | Binary | 1.04 | Literature | 0.05 | 0.05 | Bagnardi V, Blangiardo M, La Vecchia C, Corrao G. Alcohol consumption and the risk of cancer: a meta-analysis. Alcohol Res Health. 2001;25(4):263-70. https://www.ncbi.nlm.nih.gov/pmc/articles/PMC6705703/ |
| Lung Female Nonsmoker | Chronic bronchitis | Binary | 1.47 | Literature | 0.02 | 0.02 | Brenner DR, Boffetta P, Duell EJ, et al. Previous lung diseases and lung cancer risk: a pooled analysis from the International Lung Cancer Consortium. Am J Epidemiol. Oct 1 2012;176(7):573-85. doi:10.1093/aje/kws151;176(7):573. Epub 2012 Sep 17. |
| Lung Female Nonsmoker | Emphysema | Binary | 2.44 | Literature | 0.04 | 0.04 | Brenner DR, Boffetta P, Duell EJ, et al. Previous lung diseases and lung cancer risk: a pooled analysis from the International Lung Cancer Consortium. Am J Epidemiol. Oct 1 2012;176(7):573-85. doi:10.1093/aje/kws151 |
| Lung Female Nonsmoker | Family history of lung cancer in first degree relative | Binary | 1.14 | UK Biobank | 0.15 |  | Matakidou A, Eisen T, Houlston RS. Systematic review of the relationship between family history and lung cancer risk. Br J Cancer. Oct 3 2005;93(7):825-33. doi:10.1038/sj.bjc.6602769 |
| Lung Female Nonsmoker | High levels of radon in home | Binary | 1.11 | Literature | 0.16 | 0.16 | Darby S, Hill D, Auvinen A, et al. Radon in homes and risk of lung cancer: collaborative analysis of individual data from 13 European case-control studies. Bmj. Jan 29 2005;330(7485):223. doi:10.1136/bmj.38308.477650.63 |
| Lung Female Nonsmoker | History of pneumonia | Binary | 1.57 | Literature | 0.25 | 0.25 | Brenner DR, Boffetta P, Duell EJ, et al. Previous lung diseases and lung cancer risk: a pooled analysis from the International Lung Cancer Consortium. Am J Epidemiol. Oct 1 2012;176(7):573-85. doi:10.1093/aje/kws151 |
| Lung Female Nonsmoker | Maternal smoking around birth | Binary | 1.11 | UK Biobank | 0.25 |  | Asomaning K, Miller DP, Liu G, Wain JC, Lynch TJ, Su L, Christiani DC. Second hand smoke, age of exposure and lung cancer risk. Lung Cancer. Jul 2008;61(1):13-20. doi:10.1016/j.lungcan.2007.11.013 |
| Lung Female Nonsmoker | Minutes of vigorous activity daily (<30 minutes, 30-59 minutes, 60-89 minutes, >90 minutes) | Ordinal | 0.97 | UK Biobank | 1.00 |  | World Cancer Research Fund/American Institute for Cancer Research. Continuous Update Project Expert Report 2018. Diet, nutrition, physical activity, and lung cancer. 2017. https://www.wcrf.org/wp-content/uploads/2021/02/lung-cancer-report.pdf |
| Lung Female Nonsmoker | Occupational asbestos exposure | Binary | 1.49 | Literature | 0.00 | 0.00 | Moon EK, Son M, Jin YW, Park S, Lee WJ. Variations of lung cancer risk from asbestos exposure: impact on estimation of population attributable fraction. Ind Health. 2013;51(1):128-33. doi:10.2486/indhealth.ms1350 |
| Lung Female Nonsmoker | Secondhand smoke at home 1-19 years | Binary | 1.10 | Literature | 0.15 | 0.15 | Kim CH, Lee YC, Hung RJ, et al. Exposure to secondhand tobacco smoke and lung cancer by histological type: a pooled analysis of the International Lung Cancer Consortium (ILCCO). Int J Cancer. Oct 15 2014;135(8):1918-30. doi:10.1002/ijc.28835 |
| Lung Female Nonsmoker | Secondhand smoke at home 20+ years | Binary | 1.36 | Literature | 0.06 | 0.06 | Kim CH, Lee YC, Hung RJ, et al. Exposure to secondhand tobacco smoke and lung cancer by histological type: a pooled analysis of the International Lung Cancer Consortium (ILCCO). Int J Cancer. Oct 15 2014;135(8):1918-30. doi:10.1002/ijc.28835 |
| Lung Female Nonsmoker | Secondhand smoke at work 1-19 years | Binary | 1.03 | Literature | 0.15 | 0.15 | Kim CH, Lee YC, Hung RJ, et al. Exposure to secondhand tobacco smoke and lung cancer by histological type: a pooled analysis of the International Lung Cancer Consortium (ILCCO). Int J Cancer. Oct 15 2014;135(8):1918-30. doi:10.1002/ijc.28835 |
| Lung Female Nonsmoker | Secondhand smoke at work 20+ years | Binary | 1.19 | Literature | 0.06 | 0.06 | Kim CH, Lee YC, Hung RJ, et al. Exposure to secondhand tobacco smoke and lung cancer by histological type: a pooled analysis of the International Lung Cancer Consortium (ILCCO). Int J Cancer. Oct 15 2014;135(8):1918-30. doi:10.1002/ijc.28835 |
| Lung Female Nonsmoker | Servings of fruit per day (square-root transformation | Continuous | 0.89 | UK Biobank | 1.00 |  | Koumbi L. Dietary factors can protect against liver cancer development. World J Hepatol. Jan 28 2017;9(3):119-125. doi:10.4254/wjh.v9.i3.119 |
| Lung Female Nonsmoker | Servings of vegetables per day (square-root transformation) | Continuous | 0.89 | UK Biobank | 1.22 |  | World Cancer Research Fund/American Institute for Cancer Research. Continuous Update Project Expert Report 2018. Diet, nutrition, physical activity, and lung cancer. 2017. https://www.wcrf.org/wp-content/uploads/2021/02/lung-cancer-report.pdf |
| Lung Female Smoker | Alcohol consumption (2-3 drinks/day) | Binary | 1.02 | Literature | 0.12 | 0.12 | Bagnardi V, Blangiardo M, La Vecchia C, Corrao G. Alcohol consumption and the risk of cancer: a meta-analysis. Alcohol Res Health. 2001;25(4):263-70. https://www.ncbi.nlm.nih.gov/pmc/articles/PMC6705703/ |
| Lung Female Smoker | Alcohol consumption (4+ drinks/day) | Binary | 1.04 | Literature | 0.05 | 0.05 | Bagnardi V, Blangiardo M, La Vecchia C, Corrao G. Alcohol consumption and the risk of cancer: a meta-analysis. Alcohol Res Health. 2001;25(4):263-70. https://www.ncbi.nlm.nih.gov/pmc/articles/PMC6705703/ |
| Lung Female Smoker | Chronic bronchitis | Binary | 1.47 | Literature | 0.02 | 0.02 | Brenner DR, Boffetta P, Duell EJ, et al. Previous lung diseases and lung cancer risk: a pooled analysis from the International Lung Cancer Consortium. Am J Epidemiol. Oct 1 2012;176(7):573-85. doi:10.1093/aje/kws151;176(7):573. Epub 2012 Sep 17. |
| Lung Female Smoker | Emphysema | Binary | 2.44 | Literature | 0.04 | 0.04 | Brenner DR, Boffetta P, Duell EJ, et al. Previous lung diseases and lung cancer risk: a pooled analysis from the International Lung Cancer Consortium. Am J Epidemiol. Oct 1 2012;176(7):573-85. doi:10.1093/aje/kws151 |
| Lung Female Smoker | Family history of lung cancer in first degree relative | Binary | 1.03 | UK Biobank | 0.15 |  | Matakidou A, Eisen T, Houlston RS. Systematic review of the relationship between family history and lung cancer risk. Br J Cancer. Oct 3 2005;93(7):825-33. doi:10.1038/sj.bjc.6602769 |
| Lung Female Smoker | High levels of radon in home | Binary | 1.11 | Literature | 0.16 | 0.16 | Darby S, Hill D, Auvinen A, et al. Radon in homes and risk of lung cancer: collaborative analysis of individual data from 13 European case-control studies. Bmj. Jan 29 2005;330(7485):223. doi:10.1136/bmj.38308.477650.63 |
| Lung Female Smoker | High-dose (20-30mg/day) beta carotene supplementation | Binary | 1.24 | Literature | 0.40 | 0.40 | World Cancer Research Fund/American Institute for Cancer Research. Continuous Update Project Expert Report 2018. Diet, nutrition, physical activity, and lung cancer. 2017. https://www.wcrf.org/wp-content/uploads/2021/02/lung-cancer-report.pdf |
| Lung Female Smoker | History of pneumonia | Binary | 1.57 | Literature | 0.25 | 0.25 | Brenner DR, Boffetta P, Duell EJ, et al. Previous lung diseases and lung cancer risk: a pooled analysis from the International Lung Cancer Consortium. Am J Epidemiol. Oct 1 2012;176(7):573-85. doi:10.1093/aje/kws151 |
| Lung Female Smoker | Maternal smoking around birth | Binary | 1.36 | UK Biobank | 0.25 |  | Asomaning K, Miller DP, Liu G, Wain JC, Lynch TJ, Su L, Christiani DC. Second hand smoke, age of exposure and lung cancer risk. Lung Cancer. Jul 2008;61(1):13-20. doi:10.1016/j.lungcan.2007.11.013 |
| Lung Female Smoker | Minutes of moderate activity daily (<30 minutes, 30-59 minutes, 60-89 minutes, >90 minutes) | Ordinal | 0.97 | UK Biobank | 1.00 |  | World Cancer Research Fund/American Institute for Cancer Research. Continuous Update Project Expert Report 2018. Diet, nutrition, physical activity, and lung cancer. 2017. https://www.wcrf.org/wp-content/uploads/2021/02/lung-cancer-report.pdf |
| Lung Female Smoker | Minutes of vigorous activity daily (<30 minutes, 30-59 minutes, 60-89 minutes, >90 minutes) | Ordinal | 0.93 | UK Biobank | 1.00 |  | World Cancer Research Fund/American Institute for Cancer Research. Continuous Update Project Expert Report 2018. Diet, nutrition, physical activity, and lung cancer. 2017. https://www.wcrf.org/wp-content/uploads/2021/02/lung-cancer-report.pdf |
| Lung Female Smoker | Occupational asbestos exposure | Binary | 1.49 | Literature | 0.00 | 0.00 | Moon EK, Son M, Jin YW, Park S, Lee WJ. Variations of lung cancer risk from asbestos exposure: impact on estimation of population attributable fraction. Ind Health. 2013;51(1):128-33. doi:10.2486/indhealth.ms1350 |
| Lung Female Smoker | Pack-years smoking history | Continuous | 1.02 | UK Biobank | 22.20 |  | Alberg AJ, Samet JM. Epidemiology of lung cancer. Chest. Jan 2003;123(1 Suppl):21s-49s. doi:10.1378/chest.123.1_suppl.21s¶Thun MJ, Carter BD, Feskanich D, et al. 50-year trends in smoking-related mortality in the United States. N Engl J Med. Jan 24 2013;368(4):351-64. doi:10.1056/NEJMsa1211127 |
| Lung Female Smoker | Servings of fruit per day (square-root transformation | Continuous | 0.83 | UK Biobank | 1.00 |  | World Cancer Research Fund/American Institute for Cancer Research. Continuous Update Project Expert Report 2018. Diet, nutrition, physical activity, and lung cancer. 2017. https://www.wcrf.org/wp-content/uploads/2021/02/lung-cancer-report.pdf |
| Lung Female Smoker | Servings of red meat (beef, lamb, or pork) per week (square-root transformation) | Binary | 1.03 | UK Biobank | 2.12 |  | World Cancer Research Fund/American Institute for Cancer Research. Continuous Update Project Expert Report 2018. Diet, nutrition, physical activity, and lung cancer. 2017. https://www.wcrf.org/wp-content/uploads/2021/02/lung-cancer-report.pdf |
| Lung Female Smoker | Servings of vegetables per day (square-root transformation) | Continuous | 0.99 | UK Biobank | 1.22 |  | World Cancer Research Fund/American Institute for Cancer Research. Continuous Update Project Expert Report 2018. Diet, nutrition, physical activity, and lung cancer. 2017. https://www.wcrf.org/wp-content/uploads/2021/02/lung-cancer-report.pdf |
| Lung Male Nonsmoker | Alcohol consumption (2-3 drinks/day) | Binary | 1.02 | Literature | 0.12 | 0.12 | Bagnardi V, Blangiardo M, La Vecchia C, Corrao G. Alcohol consumption and the risk of cancer: a meta-analysis. Alcohol Res Health. 2001;25(4):263-70. https://www.ncbi.nlm.nih.gov/pmc/articles/PMC6705703/ |
| Lung Male Nonsmoker | Alcohol consumption (4+ drinks/day) | Binary | 1.04 | Literature | 0.05 | 0.05 | Bagnardi V, Blangiardo M, La Vecchia C, Corrao G. Alcohol consumption and the risk of cancer: a meta-analysis. Alcohol Res Health. 2001;25(4):263-70. https://www.ncbi.nlm.nih.gov/pmc/articles/PMC6705703/ |
| Lung Male Nonsmoker | Chronic bronchitis | Binary | 1.47 | Literature | 0.02 | 0.02 | Brenner DR, Boffetta P, Duell EJ, et al. Previous lung diseases and lung cancer risk: a pooled analysis from the International Lung Cancer Consortium. Am J Epidemiol. Oct 1 2012;176(7):573-85. doi:10.1093/aje/kws151;176(7):573. Epub 2012 Sep 17. |
| Lung Male Nonsmoker | Emphysema | Binary | 2.44 | Literature | 0.04 | 0.04 | Brenner DR, Boffetta P, Duell EJ, et al. Previous lung diseases and lung cancer risk: a pooled analysis from the International Lung Cancer Consortium. Am J Epidemiol. Oct 1 2012;176(7):573-85. doi:10.1093/aje/kws151 |
| Lung Male Nonsmoker | Family history of lung cancer in first degree relative | Binary | 1.14 | UK Biobank | 0.14 |  | Matakidou A, Eisen T, Houlston RS. Systematic review of the relationship between family history and lung cancer risk. Br J Cancer. Oct 3 2005;93(7):825-33. doi:10.1038/sj.bjc.6602769 |
| Lung Male Nonsmoker | High levels of radon in home | Binary | 1.11 | Literature | 0.16 | 0.16 | Darby S, Hill D, Auvinen A, et al. Radon in homes and risk of lung cancer: collaborative analysis of individual data from 13 European case-control studies. Bmj. Jan 29 2005;330(7485):223. doi:10.1136/bmj.38308.477650.63 |
| Lung Male Nonsmoker | History of pneumonia | Binary | 1.57 | Literature | 0.25 | 0.25 | Brenner DR, Boffetta P, Duell EJ, et al. Previous lung diseases and lung cancer risk: a pooled analysis from the International Lung Cancer Consortium. Am J Epidemiol. Oct 1 2012;176(7):573-85. doi:10.1093/aje/kws151 |
| Lung Male Nonsmoker | Maternal smoking around birth | Binary | 1.11 | UK Biobank | 0.26 |  | Asomaning K, Miller DP, Liu G, Wain JC, Lynch TJ, Su L, Christiani DC. Second hand smoke, age of exposure and lung cancer risk. Lung Cancer. Jul 2008;61(1):13-20. doi:10.1016/j.lungcan.2007.11.013 |
| Lung Male Nonsmoker | Minutes of vigorous activity daily (<30 minutes, 30-59 minutes, 60-89 minutes, >90 minutes) | Ordinal | 0.97 | UK Biobank | 1.00 |  | World Cancer Research Fund/American Institute for Cancer Research. Continuous Update Project Expert Report 2018. Diet, nutrition, physical activity, and lung cancer. 2017. https://www.wcrf.org/wp-content/uploads/2021/02/lung-cancer-report.pdf |
| Lung Male Nonsmoker | Occupational asbestos exposure | Binary | 1.49 | Literature | 0.00 | 0.00 | Moon EK, Son M, Jin YW, Park S, Lee WJ. Variations of lung cancer risk from asbestos exposure: impact on estimation of population attributable fraction. Ind Health. 2013;51(1):128-33. doi:10.2486/indhealth.ms1350 |
| Lung Male Nonsmoker | Secondhand smoke at home 1-19 years | Binary | 1.10 | Literature | 0.15 | 0.15 | Kim CH, Lee YC, Hung RJ, et al. Exposure to secondhand tobacco smoke and lung cancer by histological type: a pooled analysis of the International Lung Cancer Consortium (ILCCO). Int J Cancer. Oct 15 2014;135(8):1918-30. doi:10.1002/ijc.28835 |
| Lung Male Nonsmoker | Secondhand smoke at home 20+ years | Binary | 1.36 | Literature | 0.06 | 0.06 | Kim CH, Lee YC, Hung RJ, et al. Exposure to secondhand tobacco smoke and lung cancer by histological type: a pooled analysis of the International Lung Cancer Consortium (ILCCO). Int J Cancer. Oct 15 2014;135(8):1918-30. doi:10.1002/ijc.28835 |
| Lung Male Nonsmoker | Secondhand smoke at work 1-19 years | Binary | 1.03 | Literature | 0.15 | 0.15 | Kim CH, Lee YC, Hung RJ, et al. Exposure to secondhand tobacco smoke and lung cancer by histological type: a pooled analysis of the International Lung Cancer Consortium (ILCCO). Int J Cancer. Oct 15 2014;135(8):1918-30. doi:10.1002/ijc.28835 |
| Lung Male Nonsmoker | Secondhand smoke at work 20+ years | Binary | 1.19 | Literature | 0.06 | 0.06 | Kim CH, Lee YC, Hung RJ, et al. Exposure to secondhand tobacco smoke and lung cancer by histological type: a pooled analysis of the International Lung Cancer Consortium (ILCCO). Int J Cancer. Oct 15 2014;135(8):1918-30. doi:10.1002/ijc.28835 |
| Lung Male Nonsmoker | Servings of fruit per day (square-root transformation | Continuous | 0.89 | UK Biobank | 1.00 |  | World Cancer Research Fund/American Institute for Cancer Research. Continuous Update Project Expert Report 2018. Diet, nutrition, physical activity, and lung cancer. 2017. https://www.wcrf.org/wp-content/uploads/2021/02/lung-cancer-report.pdf |
| Lung Male Nonsmoker | Servings of vegetables per day (square-root transformation) | Continuous | 0.89 | UK Biobank | 1.22 |  | World Cancer Research Fund/American Institute for Cancer Research. Continuous Update Project Expert Report 2018. Diet, nutrition, physical activity, and lung cancer. 2017. https://www.wcrf.org/wp-content/uploads/2021/02/lung-cancer-report.pdf |
| Lung Male Smoker | Alcohol consumption (2-3 drinks/day) | Binary | 1.02 | Literature | 0.12 | 0.12 | Bagnardi V, Blangiardo M, La Vecchia C, Corrao G. Alcohol consumption and the risk of cancer: a meta-analysis. Alcohol Res Health. 2001;25(4):263-70. https://www.ncbi.nlm.nih.gov/pmc/articles/PMC6705703/ |
| Lung Male Smoker | Alcohol consumption (4+ drinks/day) | Binary | 1.04 | Literature | 0.05 | 0.05 | Bagnardi V, Blangiardo M, La Vecchia C, Corrao G. Alcohol consumption and the risk of cancer: a meta-analysis. Alcohol Res Health. 2001;25(4):263-70. https://www.ncbi.nlm.nih.gov/pmc/articles/PMC6705703/ |
| Lung Male Smoker | Chronic bronchitis | Binary | 1.47 | Literature | 0.02 | 0.02 | Brenner DR, Boffetta P, Duell EJ, et al. Previous lung diseases and lung cancer risk: a pooled analysis from the International Lung Cancer Consortium. Am J Epidemiol. Oct 1 2012;176(7):573-85. doi:10.1093/aje/kws151;176(7):573. Epub 2012 Sep 17. |
| Lung Male Smoker | Emphysema | Binary | 2.44 | Literature | 0.04 | 0.04 | Brenner DR, Boffetta P, Duell EJ, et al. Previous lung diseases and lung cancer risk: a pooled analysis from the International Lung Cancer Consortium. Am J Epidemiol. Oct 1 2012;176(7):573-85. doi:10.1093/aje/kws151 |
| Lung Male Smoker | Family history of lung cancer in first degree relative | Binary | 1.03 | UK Biobank | 0.14 |  | Matakidou A, Eisen T, Houlston RS. Systematic review of the relationship between family history and lung cancer risk. Br J Cancer. Oct 3 2005;93(7):825-33. doi:10.1038/sj.bjc.6602769 |
| Lung Male Smoker | High levels of radon in home | Binary | 1.11 | Literature | 0.16 | 0.16 | Darby S, Hill D, Auvinen A, et al. Radon in homes and risk of lung cancer: collaborative analysis of individual data from 13 European case-control studies. Bmj. Jan 29 2005;330(7485):223. doi:10.1136/bmj.38308.477650.63 |
| Lung Male Smoker | High-dose (20-30mg/day) beta carotene supplementation | Binary | 1.24 | Literature | 0.40 | 0.40 | World Cancer Research Fund/American Institute for Cancer Research. Continuous Update Project Expert Report 2018. Diet, nutrition, physical activity, and lung cancer. 2017. https://www.wcrf.org/wp-content/uploads/2021/02/lung-cancer-report.pdf |
| Lung Male Smoker | History of pneumonia | Binary | 1.57 | Literature | 0.25 | 0.25 | Brenner DR, Boffetta P, Duell EJ, et al. Previous lung diseases and lung cancer risk: a pooled analysis from the International Lung Cancer Consortium. Am J Epidemiol. Oct 1 2012;176(7):573-85. doi:10.1093/aje/kws151 |
| Lung Male Smoker | Maternal smoking around birth | Binary | 1.36 | UK Biobank | 0.26 |  | Asomaning K, Miller DP, Liu G, Wain JC, Lynch TJ, Su L, Christiani DC. Second hand smoke, age of exposure and lung cancer risk. Lung Cancer. Jul 2008;61(1):13-20. doi:10.1016/j.lungcan.2007.11.013 |
| Lung Male Smoker | Minutes of moderate activity daily (<30 minutes, 30-59 minutes, 60-89 minutes, >90 minutes) | Ordinal | 0.97 | UK Biobank | 1.00 |  | World Cancer Research Fund/American Institute for Cancer Research. Continuous Update Project Expert Report 2018. Diet, nutrition, physical activity, and lung cancer. 2017. https://www.wcrf.org/wp-content/uploads/2021/02/lung-cancer-report.pdf |
| Lung Male Smoker | Minutes of vigorous activity daily (<30 minutes, 30-59 minutes, 60-89 minutes, >90 minutes) | Ordinal | 0.93 | UK Biobank | 1.00 |  | World Cancer Research Fund/American Institute for Cancer Research. Continuous Update Project Expert Report 2018. Diet, nutrition, physical activity, and lung cancer. 2017. https://www.wcrf.org/wp-content/uploads/2021/02/lung-cancer-report.pdf |
| Lung Male Smoker | Occupational asbestos exposure | Binary | 1.49 | Literature | 0.00 | 0.00 | Moon EK, Son M, Jin YW, Park S, Lee WJ. Variations of lung cancer risk from asbestos exposure: impact on estimation of population attributable fraction. Ind Health. 2013;51(1):128-33. doi:10.2486/indhealth.ms1350 |
| Lung Male Smoker | Pack-years smoking history | Continuous | 1.02 | UK Biobank | 29.09 |  | Alberg AJ, Samet JM. Epidemiology of lung cancer. Chest. Jan 2003;123(1 Suppl):21s-49s. doi:10.1378/chest.123.1_suppl.21s¶Thun MJ, Carter BD, Feskanich D, et al. 50-year trends in smoking-related mortality in the United States. N Engl J Med. Jan 24 2013;368(4):351-64. doi:10.1056/NEJMsa1211127 |
| Lung Male Smoker | Servings of fruit per day (square-root transformation | Continuous | 0.83 | UK Biobank | 1.00 |  | World Cancer Research Fund/American Institute for Cancer Research. Continuous Update Project Expert Report 2018. Diet, nutrition, physical activity, and lung cancer. 2017. https://www.wcrf.org/wp-content/uploads/2021/02/lung-cancer-report.pdf |
| Lung Male Smoker | Servings of red meat (beef, lamb, or pork) per week (square-root transformation) | Binary | 1.03 | UK Biobank | 2.12 |  | World Cancer Research Fund/American Institute for Cancer Research. Continuous Update Project Expert Report 2018. Diet, nutrition, physical activity, and lung cancer. 2017. https://www.wcrf.org/wp-content/uploads/2021/02/lung-cancer-report.pdf |
| Lung Male Smoker | Servings of vegetables per day (square-root transformation) | Continuous | 0.99 | UK Biobank | 1.22 |  | World Cancer Research Fund/American Institute for Cancer Research. Continuous Update Project Expert Report 2018. Diet, nutrition, physical activity, and lung cancer. 2017. https://www.wcrf.org/wp-content/uploads/2021/02/lung-cancer-report.pdf |
| Lymphoma Female | Body mass index | Continuous | 1.01 | UK Biobank | 29.80 |  | Strongman H, Brown A, Smeeth L, Bhaskaran K. Body mass index and Hodgkin's lymphoma: UK population-based cohort study of 5.8 million individuals. British Journal of Cancer. 2019/04/01 2019;120(7):768-770. doi:10.1038/s41416-019-0401-1 |
| Lymphoma Female | Celiac disease | Binary | 2.06 | UK Biobank | 0.01 |  | Smedby KE, Hjalgrim H, Askling J, et al. Autoimmune and chronic inflammatory disorders and risk of non-Hodgkin lymphoma by subtype. J Natl Cancer Inst. Jan 4 2006;98(1):51-60. doi:10.1093/jnci/djj004 |
| Lymphoma Female | Chronic hepatitis C | Binary | 1.50 | UK Biobank | 0.00 |  | Talamini R, Montella M, Crovatto M, et al. Non-Hodgkin's lymphoma and hepatitis C virus: a case-control study from northern and southern Italy. Int J Cancer. Jun 20 2004;110(3):380-5. doi:10.1002/ijc.20137 |
| Lymphoma Female | Family history of lymphoma in first degree relative | Binary | 3.30 | Literature | 0.00 | 0.00 | Goldin LR, Pfeiffer RM, Gridley G, et al. Familial aggregation of Hodgkin lymphoma and related tumors. Cancer. May 1 2004;100(9):1902-8. doi:10.1002/cncr.20189 |
| Lymphoma Female | Hashimoto's thyroiditis | Binary | 1.98 | UK Biobank | 0.00 |  | Hyjek E, Isaacson PG. Primary B cell lymphoma of the thyroid and its relationship to Hashimoto's Thyroiditis. Human Pathology. 1988/11/01/ 1988;19(11):1315-1326. doi:https://doi.org/10.1016/S0046-8177(88)80287-9 |
| Lymphoma Female | Inflammatory bowel disease | Binary | 1.64 | UK Biobank | 0.01 |  | Kandiel A, Fraser AG, Korelitz BI, Brensinger C, Lewis JD. Increased risk of lymphoma among inflammatory bowel disease patients treated with azathioprine and 6-mercaptopurine. Gut. Aug 2005;54(8):1121-5. doi:10.1136/gut.2004.049460 |
| Lymphoma Female | Minutes of moderate activity daily (<30 minutes, 30-59 minutes, 60-89 minutes, >90 minutes) | Ordinal | 0.97 | UK Biobank | 1.00 |  | Davies GA, Strader C, Chibbar R, Papatheodorou S, Dmytriw AA. The relationship between physical activity and lymphoma: a systematic review and meta analysis. BMC Cancer. Oct 6 2020;20(1):962. doi:10.1186/s12885-020-07431-x |
| Lymphoma Female | Pack-years smoking history (fifth-root transformation) | Continuous | 1.06 | UK Biobank | 0.00 |  | Sergentanis TN, Kanavidis P, Michelakos T, Petridou ET. Cigarette smoking and risk of lymphoma in adults: a comprehensive meta-analysis on Hodgkin and non-Hodgkin disease. Eur J Cancer Prev. Mar 2013;22(2):131-50. doi:10.1097/CEJ.0b013e328355ed08 |
| Lymphoma Female | Rheumatoid arthritis | Binary | 1.75 | UK Biobank | 0.03 |  | Fallah M, Liu X, Ji J, Försti A, Sundquist K, Hemminki K. Hodgkin lymphoma after autoimmune diseases by age at diagnosis and histological subtype. Ann Oncol. Jul 2014;25(7):1397-1404. doi:10.1093/annonc/mdu144 |
| Lymphoma Female | Servings of processed meat (bacon, ham, sausages, burgers, nuggets, etc.) per week (square-root transformation) | Continuous | 1.08 | UK Biobank | 1.64 |  | Rohrmann S, Linseisen J, Jakobsen MU, et al. Consumption of meat and dairy and lymphoma risk in the European Prospective Investigation into Cancer and Nutrition. International Journal of Cancer. 2011;128(3):623-634. doi:https://doi.org/10.1002/ijc.25387 |
| Lymphoma Female | Sjogren's syndrome | Binary | 3.76 | UK Biobank | 0.00 |  | Fallah M, Liu X, Ji J, Försti A, Sundquist K, Hemminki K. Hodgkin lymphoma after autoimmune diseases by age at diagnosis and histological subtype. Ann Oncol. Jul 2014;25(7):1397-1404. doi:10.1093/annonc/mdu144 |
| Lymphoma Female | Systemic lupus erythematosus | Binary | 1.67 | UK Biobank | 0.00 |  | Fallah M, Liu X, Ji J, Försti A, Sundquist K, Hemminki K. Hodgkin lymphoma after autoimmune diseases by age at diagnosis and histological subtype. Ann Oncol. Jul 2014;25(7):1397-1404. doi:10.1093/annonc/mdu144 |
| Lymphoma Male | Aspirin use | Binary | 0.94 | UK Biobank | 0.19 |  | Liebow M, Larson MC, Thompson CA, et al. Aspirin and other nonsteroidal anti-inflammatory drugs, statins and risk of non-Hodgkin lymphoma. International Journal of Cancer. 2021;149(3):535-545. doi:https://doi.org/10.1002/ijc.33541 |
| Lymphoma Male | Body mass index | Continuous | 1.02 | UK Biobank | 29.40 |  | Strongman H, Brown A, Smeeth L, Bhaskaran K. Body mass index and Hodgkin's lymphoma: UK population-based cohort study of 5.8 million individuals. British Journal of Cancer. 2019/04/01 2019;120(7):768-770. doi:10.1038/s41416-019-0401-1 |
| Lymphoma Male | Celiac disease | Binary | 2.79 | UK Biobank | 0.01 |  | Smedby KE, Hjalgrim H, Askling J, et al. Autoimmune and chronic inflammatory disorders and risk of non-Hodgkin lymphoma by subtype. J Natl Cancer Inst. Jan 4 2006;98(1):51-60. doi:10.1093/jnci/djj004 |
| Lymphoma Male | Chronic hepatitis C | Binary | 3.23 | UK Biobank | 0.00 |  | Talamini R, Montella M, Crovatto M, et al. Non-Hodgkin's lymphoma and hepatitis C virus: a case-control study from northern and southern Italy. Int J Cancer. Jun 20 2004;110(3):380-5. doi:10.1002/ijc.20137 |
| Lymphoma Male | Family history of lymphoma in first degree relative | Binary | 3.30 | Literature | 0.00 | 0.00 | Goldin LR, Pfeiffer RM, Gridley G, et al. Familial aggregation of Hodgkin lymphoma and related tumors. Cancer. May 1 2004;100(9):1902-8. doi:10.1002/cncr.20189 |
| Lymphoma Male | Hashimoto's thyroiditis | Binary | 10.11 | UK Biobank | 0.00 |  | Hyjek E, Isaacson PG. Primary B cell lymphoma of the thyroid and its relationship to Hashimoto's Thyroiditis. Human Pathology. 1988/11/01/ 1988;19(11):1315-1326. doi:https://doi.org/10.1016/S0046-8177(88)80287-9 |
| Lymphoma Male | Inflammatory bowel disease | Binary | 1.40 | UK Biobank | 0.02 |  | Kandiel A, Fraser AG, Korelitz BI, Brensinger C, Lewis JD. Increased risk of lymphoma among inflammatory bowel disease patients treated with azathioprine and 6-mercaptopurine. Gut. Aug 2005;54(8):1121-5. doi:10.1136/gut.2004.049460 |
| Lymphoma Male | Minutes of vigorous activity daily (<30 minutes, 30-59 minutes, 60-89 minutes, >90 minutes) | Ordinal | 0.96 | UK Biobank | 1.00 |  | Davies GA, Strader C, Chibbar R, Papatheodorou S, Dmytriw AA. The relationship between physical activity and lymphoma: a systematic review and meta analysis. BMC Cancer. Oct 6 2020;20(1):962. doi:10.1186/s12885-020-07431-x |
| Lymphoma Male | Rheumatoid arthritis | Binary | 2.00 | UK Biobank | 0.01 |  | Fallah M, Liu X, Ji J, Försti A, Sundquist K, Hemminki K. Hodgkin lymphoma after autoimmune diseases by age at diagnosis and histological subtype. Ann Oncol. Jul 2014;25(7):1397-1404. doi:10.1093/annonc/mdu144 |
| Lymphoma Male | Sjogren's syndrome | Binary | 5.07 | UK Biobank | 0.00 |  | Fallah M, Liu X, Ji J, Försti A, Sundquist K, Hemminki K. Hodgkin lymphoma after autoimmune diseases by age at diagnosis and histological subtype. Ann Oncol. Jul 2014;25(7):1397-1404. doi:10.1093/annonc/mdu144 |
| Lymphoma Male | Smoking (current) | Binary | 1.18 | UK Biobank | 0.13 |  | Sergentanis TN, Kanavidis P, Michelakos T, Petridou ET. Cigarette smoking and risk of lymphoma in adults: a comprehensive meta-analysis on Hodgkin and non-Hodgkin disease. Eur J Cancer Prev. Mar 2013;22(2):131-50. doi:10.1097/CEJ.0b013e328355ed08 |
| Lymphoma Male | Systemic lupus erythematosus | Binary | 2.91 | UK Biobank | 0.00 |  | Fallah M, Liu X, Ji J, Försti A, Sundquist K, Hemminki K. Hodgkin lymphoma after autoimmune diseases by age at diagnosis and histological subtype. Ann Oncol. Jul 2014;25(7):1397-1404. doi:10.1093/annonc/mdu144 |
| Melanoma Female | >100 moles on entire body | Binary | 4.69 | Literature | 0.00 | 0.02 | Swerdlow AJ, English J, MacKie RM, O'Doherty CJ, Hunter JA, Clark J, Hole DJ. Benign melanocytic naevi as a risk factor for malignant melanoma. Br Med J (Clin Res Ed). Jun 14 1986;292(6535):1555-9. doi:10.1136/bmj.292.6535.1555 |
| Melanoma Female | 16-40 moles on entire body | Binary | 1.00 | Literature | 0.00 | 0.22 | Swerdlow AJ, English J, MacKie RM, O'Doherty CJ, Hunter JA, Clark J, Hole DJ. Benign melanocytic naevi as a risk factor for malignant melanoma. Br Med J (Clin Res Ed). Jun 14 1986;292(6535):1555-9. doi:10.1136/bmj.292.6535.1555 |
| Melanoma Female | 41-60 moles on entire body | Binary | 1.52 | Literature | 0.00 | 0.18 | Swerdlow AJ, English J, MacKie RM, O'Doherty CJ, Hunter JA, Clark J, Hole DJ. Benign melanocytic naevi as a risk factor for malignant melanoma. Br Med J (Clin Res Ed). Jun 14 1986;292(6535):1555-9. doi:10.1136/bmj.292.6535.1555 |
| Melanoma Female | 61-80 moles on entire body | Binary | 2.22 | Literature | 0.00 | 0.06 | Swerdlow AJ, English J, MacKie RM, O'Doherty CJ, Hunter JA, Clark J, Hole DJ. Benign melanocytic naevi as a risk factor for malignant melanoma. Br Med J (Clin Res Ed). Jun 14 1986;292(6535):1555-9. doi:10.1136/bmj.292.6535.1555 |
| Melanoma Female | 81-100 moles on entire body | Binary | 3.22 | Literature | 0.00 | 0.06 | Swerdlow AJ, English J, MacKie RM, O'Doherty CJ, Hunter JA, Clark J, Hole DJ. Benign melanocytic naevi as a risk factor for malignant melanoma. Br Med J (Clin Res Ed). Jun 14 1986;292(6535):1555-9. doi:10.1136/bmj.292.6535.1555 |
| Melanoma Female | Blonde hair | Binary | 1.30 | UK Biobank | 0.12 |  | Gandini S, Sera F, Cattaruzza MS, et al. Meta-analysis of risk factors for cutaneous melanoma: III. Family history, actinic damage and phenotypic factors. Eur J Cancer. Sep 2005;41(14):2040-59. doi:10.1016/j.ejca.2005.03.034 |
| Melanoma Female | Cups of tea daily (fifth-root transformation) | Continuous | 0.93 | UK Biobank | 0.97 |  | Caini S, Masala G, Saieva C, et al. Coffee, tea and melanoma risk: findings from the European Prospective Investigation into Cancer and Nutrition. Int J Cancer. May 15 2017;140(10):2246-2255. doi:10.1002/ijc.30659 |
| Melanoma Female | Daily sunscreen use | Binary | 0.50 | Literature | 0.14 | 0.14 | Green AC, Williams GM, Logan V, Strutton GM. Reduced melanoma after regular sunscreen use: randomized trial follow-up. J Clin Oncol. Jan 20 2011;29(3):257-63. doi:10.1200/jco.2010.28.7078 |
| Melanoma Female | Ease of skin burning in sun (very tan, moderately tan, mildly tan, never tan/only burn) | Ordinal | 1.02 | UK Biobank | 1.00 |  | Gandini S, Sera F, Cattaruzza MS, et al. Meta-analysis of risk factors for cutaneous melanoma: III. Family history, actinic damage and phenotypic factors. Eur J Cancer. Sep 2005;41(14):2040-59. doi:10.1016/j.ejca.2005.03.034 |
| Melanoma Female | Endometriosis | Binary | 1.60 | Literature | 0.02 | 0.10 | Kvaskoff M, Mesrine S, Fournier A, Boutron-Ruault MC, Clavel-Chapelon F. Personal history of endometriosis and risk of cutaneous melanoma in a large prospective cohort of French women. Arch Intern Med. Oct 22 2007;167(19):2061-5. doi:10.1001/archinte.167.19.2061 |
| Melanoma Female | Ever used indoor tanning bed | Binary | 1.27 | Literature | 0.36 | 0.36 | An S, Kim K, Moon S, Ko KP, Kim I, Lee JE, Park SK. Indoor Tanning and the Risk of Overall and Early-Onset Melanoma and Non-Melanoma Skin Cancer: Systematic Review and Meta-Analysis. Cancers (Basel). Nov 25 2021;13(23)doi:10.3390/cancers13235940 |
| Melanoma Female | Fair skin | Binary | 1.72 | UK Biobank | 0.61 |  | Gandini S, Sera F, Cattaruzza MS, et al. Meta-analysis of risk factors for cutaneous melanoma: III. Family history, actinic damage and phenotypic factors. Eur J Cancer. Sep 2005;41(14):2040-59. doi:10.1016/j.ejca.2005.03.034 |
| Melanoma Female | Family history of melanoma in first degree relative | Binary | 1.74 | Literature | 0.03 | 0.03 | Begg CB, Hummer A, Mujumdar U, et al. Familial aggregation of melanoma risks in a large population-based sample of melanoma cases. Cancer Causes Control. Nov 2004;15(9):957-65. doi:10.1007/s10522-004-2474-2 |
| Melanoma Female | High freckle density | Binary | 2.10 | Literature | 0.09 | 0.09 | Gandini S, Sera F, Cattaruzza MS, et al. Meta-analysis of risk factors for cutaneous melanoma: III. Family history, actinic damage and phenotypic factors. Eur J Cancer. Sep 2005;41(14):2040-59. doi:10.1016/j.ejca.2005.03.034 |
| Melanoma Female | Hours spent outdoors in the summer daily (natural log transformation) | Continuous | 1.02 | UK Biobank | 0.10 |  | Whiteman DC, Stickley M, Watt P, Hughes MC, Davis MB, Green AC. Anatomic site, sun exposure, and risk of cutaneous melanoma. J Clin Oncol. Jul 1 2006;24(19):3172-7. doi:10.1200/jco.2006.06.1325 |
| Melanoma Female | Non-melanoma skin cancer | Binary | 5.34 | UK Biobank | 0.03 |  | Marghoob AA, Slade J, Salopek TG, Kopf AW, Bart RS, Rigel DS. Basal cell and squamous cell carcinomas are important risk factors for cutaneous malignant melanoma. Screening implications. Cancer. Jan 15 1995;75(2 Suppl):707-14. doi:10.1002/1097-0142(19950115)75:2+<707::aid-cncr2820751415>3.0.co;2-w |
| Melanoma Female | Number of severe sunburns before age 15 | Continuous | 1.04 | UK Biobank | 1.00 |  | Wu S, Han J, Laden F, Qureshi AA. Long-term ultraviolet flux, other potential risk factors, and skin cancer risk: a cohort study. Cancer Epidemiol Biomarkers Prev. Jun 2014;23(6):1080-9. doi:10.1158/1055-9965.Epi-13-0821 |
| Melanoma Female | Olive skin | Binary | 1.05 | UK Biobank | 0.00 |  | Gandini S, Sera F, Cattaruzza MS, et al. Meta-analysis of risk factors for cutaneous melanoma: III. Family history, actinic damage and phenotypic factors. Eur J Cancer. Sep 2005;41(14):2040-59. doi:10.1016/j.ejca.2005.03.034 |
| Melanoma Female | Red hair | Binary | 1.61 | UK Biobank | 0.05 |  | Gandini S, Sera F, Cattaruzza MS, et al. Meta-analysis of risk factors for cutaneous melanoma: III. Family history, actinic damage and phenotypic factors. Eur J Cancer. Sep 2005;41(14):2040-59. doi:10.1016/j.ejca.2005.03.034 |
| Melanoma Male | >100 moles on entire body | Binary | 4.69 | Literature | 0.00 | 0.02 | Swerdlow AJ, English J, MacKie RM, O'Doherty CJ, Hunter JA, Clark J, Hole DJ. Benign melanocytic naevi as a risk factor for malignant melanoma. Br Med J (Clin Res Ed). Jun 14 1986;292(6535):1555-9. doi:10.1136/bmj.292.6535.1555 |
| Melanoma Male | 16-40 moles on entire body | Binary | 1.00 | Literature | 0.00 | 0.22 | Swerdlow AJ, English J, MacKie RM, O'Doherty CJ, Hunter JA, Clark J, Hole DJ. Benign melanocytic naevi as a risk factor for malignant melanoma. Br Med J (Clin Res Ed). Jun 14 1986;292(6535):1555-9. doi:10.1136/bmj.292.6535.1555 |
| Melanoma Male | 41-60 moles on entire body | Binary | 1.52 | Literature | 0.00 | 0.18 | Swerdlow AJ, English J, MacKie RM, O'Doherty CJ, Hunter JA, Clark J, Hole DJ. Benign melanocytic naevi as a risk factor for malignant melanoma. Br Med J (Clin Res Ed). Jun 14 1986;292(6535):1555-9. doi:10.1136/bmj.292.6535.1555 |
| Melanoma Male | 61-80 moles on entire body | Binary | 2.22 | Literature | 0.00 | 0.06 | Swerdlow AJ, English J, MacKie RM, O'Doherty CJ, Hunter JA, Clark J, Hole DJ. Benign melanocytic naevi as a risk factor for malignant melanoma. Br Med J (Clin Res Ed). Jun 14 1986;292(6535):1555-9. doi:10.1136/bmj.292.6535.1555 |
| Melanoma Male | 81-100 moles on entire body | Binary | 3.22 | Literature | 0.00 | 0.06 | Swerdlow AJ, English J, MacKie RM, O'Doherty CJ, Hunter JA, Clark J, Hole DJ. Benign melanocytic naevi as a risk factor for malignant melanoma. Br Med J (Clin Res Ed). Jun 14 1986;292(6535):1555-9. doi:10.1136/bmj.292.6535.1555 |
| Melanoma Male | Blonde hair | Binary | 1.34 | UK Biobank | 0.09 |  | Gandini S, Sera F, Cattaruzza MS, et al. Meta-analysis of risk factors for cutaneous melanoma: III. Family history, actinic damage and phenotypic factors. Eur J Cancer. Sep 2005;41(14):2040-59. doi:10.1016/j.ejca.2005.03.034 |
| Melanoma Male | Daily sunscreen use | Binary | 0.50 | Literature | 0.14 | 0.14 | Green AC, Williams GM, Logan V, Strutton GM. Reduced melanoma after regular sunscreen use: randomized trial follow-up. J Clin Oncol. Jan 20 2011;29(3):257-63. doi:10.1200/jco.2010.28.7078 |
| Melanoma Male | Ease of skin burning in sun (very tan, moderately tan, mildly tan, never tan/only burn) | Ordinal | 1.05 | UK Biobank | 1.00 |  | Gandini S, Sera F, Cattaruzza MS, et al. Meta-analysis of risk factors for cutaneous melanoma: III. Family history, actinic damage and phenotypic factors. Eur J Cancer. Sep 2005;41(14):2040-59. doi:10.1016/j.ejca.2005.03.034 |
| Melanoma Male | Ever used indoor tanning bed | Binary | 1.27 | Literature | 0.36 | 0.36 | An S, Kim K, Moon S, Ko KP, Kim I, Lee JE, Park SK. Indoor Tanning and the Risk of Overall and Early-Onset Melanoma and Non-Melanoma Skin Cancer: Systematic Review and Meta-Analysis. Cancers (Basel). Nov 25 2021;13(23)doi:10.3390/cancers13235940 |
| Melanoma Male | Fair skin | Binary | 1.62 | UK Biobank | 0.61 |  | Gandini S, Sera F, Cattaruzza MS, et al. Meta-analysis of risk factors for cutaneous melanoma: III. Family history, actinic damage and phenotypic factors. Eur J Cancer. Sep 2005;41(14):2040-59. doi:10.1016/j.ejca.2005.03.034 |
| Melanoma Male | Family history of melanoma in first degree relative | Binary | 1.74 | Literature | 0.03 | 0.03 | Begg CB, Hummer A, Mujumdar U, et al. Familial aggregation of melanoma risks in a large population-based sample of melanoma cases. Cancer Causes Control. Nov 2004;15(9):957-65. doi:10.1007/s10522-004-2474-2 |
| Melanoma Male | High freckle density | Binary | 2.10 | Literature | 0.09 | 0.09 | Gandini S, Sera F, Cattaruzza MS, et al. Meta-analysis of risk factors for cutaneous melanoma: III. Family history, actinic damage and phenotypic factors. Eur J Cancer. Sep 2005;41(14):2040-59. doi:10.1016/j.ejca.2005.03.034 |
| Melanoma Male | Hours spent outdoors in the summer daily (natural log transformation) | Continuous | 1.02 | UK Biobank | 0.10 |  | Whiteman DC, Stickley M, Watt P, Hughes MC, Davis MB, Green AC. Anatomic site, sun exposure, and risk of cutaneous melanoma. J Clin Oncol. Jul 1 2006;24(19):3172-7. doi:10.1200/jco.2006.06.1325 |
| Melanoma Male | Non-melanoma skin cancer | Binary | 6.21 | UK Biobank | 0.05 |  | Marghoob AA, Slade J, Salopek TG, Kopf AW, Bart RS, Rigel DS. Basal cell and squamous cell carcinomas are important risk factors for cutaneous malignant melanoma. Screening implications. Cancer. Jan 15 1995;75(2 Suppl):707-14. doi:10.1002/1097-0142(19950115)75:2+<707::aid-cncr2820751415>3.0.co;2-w |
| Melanoma Male | Number of severe sunburns before age 16 | Binary | 1.03 | UK Biobank | 1.00 |  | Wu S, Han J, Laden F, Qureshi AA. Long-term ultraviolet flux, other potential risk factors, and skin cancer risk: a cohort study. Cancer Epidemiol Biomarkers Prev. Jun 2014;23(6):1080-9. doi:10.1158/1055-9965.Epi-13-0821 |
| Melanoma Male | Olive skin | Binary | 1.08 | UK Biobank | 0.00 |  | Gandini S, Sera F, Cattaruzza MS, et al. Meta-analysis of risk factors for cutaneous melanoma: III. Family history, actinic damage and phenotypic factors. Eur J Cancer. Sep 2005;41(14):2040-59. doi:10.1016/j.ejca.2005.03.034 |
| Melanoma Male | Red hair | Binary | 1.39 | UK Biobank | 0.03 |  | Gandini S, Sera F, Cattaruzza MS, et al. Meta-analysis of risk factors for cutaneous melanoma: III. Family history, actinic damage and phenotypic factors. Eur J Cancer. Sep 2005;41(14):2040-59. doi:10.1016/j.ejca.2005.03.034 |
| Multiple_Myeloma Female | Body mass index | Continuous | 1.01 | UK Biobank | 29.80 |  | Lee DJ, El-Khoury H, Tramontano AC, et al. Mass spectrometry-detected MGUS is associated with obesity and other novel modifiable risk factors in a high-risk population. Blood Advances. 2024;8(7):1737-1746. doi:10.1182/bloodadvances.2023010843 |
| Multiple_Myeloma Female | Family history of multiple myeloma in first degree relative | Binary | 3.70 | Literature | 0.00 | 0.00 | Sud A, Chattopadhyay S, Thomsen H, Sundquist K, Sundquist J, Houlston RS, Hemminki K. Analysis of 153 115 patients with hematological malignancies refines the spectrum of familial risk. Blood. Sep 19 2019;134(12):960-969. doi:10.1182/blood.2019001362 |
| Multiple_Myeloma Female | Minutes of moderate activity daily (<30 minutes, 30-59 minutes, 60-89 minutes, >90 minutes) | Ordinal | 1.00 | UK Biobank | 1.00 |  | Lee DJ, El-Khoury H, Tramontano AC, et al. Mass spectrometry-detected MGUS is associated with obesity and other novel modifiable risk factors in a high-risk population. Blood Advances. 2024;8(7):1737-1746. doi:10.1182/bloodadvances.2023010843 |
| Multiple_Myeloma Female | Minutes of vigorous activity daily (<30 minutes, 30-59 minutes, 60-89 minutes, >90 minutes) | Ordinal | 0.93 | UK Biobank | 1.00 |  | Lee DJ, El-Khoury H, Tramontano AC, et al. Mass spectrometry-detected MGUS is associated with obesity and other novel modifiable risk factors in a high-risk population. Blood Advances. 2024;8(7):1737-1746. doi:10.1182/bloodadvances.2023010843 |
| Multiple_Myeloma Female | Monoclonal gammopathy of undetermined significance | Binary | 49.51 | UK Biobank | 0.00 |  | Kyle RA, Therneau TM, Rajkumar SV, Offord JR, Larson DR, Plevak MF, Melton LJ, 3rd. A long-term study of prognosis in monoclonal gammopathy of undetermined significance. N Engl J Med. Feb 21 2002;346(8):564-9. doi:10.1056/NEJMoa01133202 |
| Multiple_Myeloma Male | Body mass index | Continuous | 1.01 | UK Biobank | 29.40 |  | Lee DJ, El-Khoury H, Tramontano AC, et al. Mass spectrometry-detected MGUS is associated with obesity and other novel modifiable risk factors in a high-risk population. Blood Advances. 2024;8(7):1737-1746. doi:10.1182/bloodadvances.2023010843 |
| Multiple_Myeloma Male | Family history of multiple myeloma in first degree relative | Binary | 3.70 | Literature | 0.00 | 0.00 | Sud A, Chattopadhyay S, Thomsen H, Sundquist K, Sundquist J, Houlston RS, Hemminki K. Analysis of 153 115 patients with hematological malignancies refines the spectrum of familial risk. Blood. Sep 19 2019;134(12):960-969. doi:10.1182/blood.2019001362 |
| Multiple_Myeloma Male | Minutes of vigorous activity daily (<30 minutes, 30-59 minutes, 60-89 minutes, >90 minutes) | Ordinal | 0.97 | UK Biobank | 1.00 |  | Lee DJ, El-Khoury H, Tramontano AC, et al. Mass spectrometry-detected MGUS is associated with obesity and other novel modifiable risk factors in a high-risk population. Blood Advances. 2024;8(7):1737-1746. doi:10.1182/bloodadvances.2023010843 |
| Multiple_Myeloma Male | Monoclonal gammopathy of undetermined significance | Binary | 38.32 | UK Biobank | 0.00 |  | Kyle RA, Therneau TM, Rajkumar SV, Offord JR, Larson DR, Plevak MF, Melton LJ, 3rd. A long-term study of prognosis in monoclonal gammopathy of undetermined significance. N Engl J Med. Feb 21 2002;346(8):564-9. doi:10.1056/NEJMoa01133202 |
| Ovarian Female | Alcohol consumption (2-3 drinks/day) | Binary | 1.11 | Literature | 0.12 | 0.12 | Bagnardi V, Blangiardo M, La Vecchia C, Corrao G. Alcohol consumption and the risk of cancer: a meta-analysis. Alcohol Res Health. 2001;25(4):263-70. https://www.ncbi.nlm.nih.gov/pmc/articles/PMC6705703/ |
| Ovarian Female | Alcohol consumption (4+ drinks/day) | Binary | 1.23 | Literature | 0.05 | 0.05 | Bagnardi V, Blangiardo M, La Vecchia C, Corrao G. Alcohol consumption and the risk of cancer: a meta-analysis. Alcohol Res Health. 2001;25(4):263-70. https://www.ncbi.nlm.nih.gov/pmc/articles/PMC6705703/ |
| Ovarian Female | Aspirin use | Binary | 0.88 | UK Biobank | 0.10 |  | Trabert B, Ness RB, Lo-Ciganic WH, et al. Aspirin, nonaspirin nonsteroidal anti-inflammatory drug, and acetaminophen use and risk of invasive epithelial ovarian cancer: a pooled analysis in the Ovarian Cancer Association Consortium. J Natl Cancer Inst. Feb 2014;106(2):djt431. doi:10.1093/jnci/djt431 |
| Ovarian Female | Breastfeeding >12 months | Binary | 0.64 | Literature | 0.25 | 0.25 | Li DP, Du C, Zhang ZM, et al. Breastfeeding and ovarian cancer risk: a systematic review and meta-analysis of 40 epidemiological studies. Asian Pac J Cancer Prev. 2014;15(12):4829-37. doi:10.7314/apjcp.2014.15.12.4829 |
| Ovarian Female | Breastfeeding 12-17 months | Binary | 0.73 | Literature | 0.20 | 0.20 | Li DP, Du C, Zhang ZM, et al. Breastfeeding and ovarian cancer risk: a systematic review and meta-analysis of 40 epidemiological studies. Asian Pac J Cancer Prev. 2014;15(12):4829-37. doi:10.7314/apjcp.2014.15.12.4829 |
| Ovarian Female | Endometriosis | Binary | 5.08 | Literature | 0.02 | 0.10 | Barnard ME, Farland LV, Yan B, et al. Endometriosis Typology and Ovarian Cancer Risk. Jama. Jul 17 2024;doi:10.1001/jama.2024.9210 |
| Ovarian Female | Family history of ovarian cancer in first degree relative | Binary | 3.12 | Literature | 0.03 | 0.03 | Kerlikowske K, Brown JS, Grady DG. Should women with familial ovarian cancer undergo prophylactic oophorectomy? Obstet Gynecol. Oct 1992;80(4):700-7. |
| Ovarian Female | Height (in cm) | Continuous | 1.02 | UK Biobank | 162.36 |  | Green J, Cairns BJ, Casabonne D, Wright FL, Reeves G, Beral V. Height and cancer incidence in the Million Women Study: prospective cohort, and meta-analysis of prospective studies of height and total cancer risk. Lancet Oncol. Aug 2011;12(8):785-94. doi:10.1016/s1470-2045(11)70154-1 |
| Ovarian Female | Hormone replacement therapy | Binary | 1.07 | UK Biobank | 0.39 |  | Beral V, Gaitskell K, Hermon C, Moser K, Reeves G, Peto R. Menopausal hormone use and ovarian cancer risk: individual participant meta-analysis of 52 epidemiological studies. Lancet. May 9 2015;385(9980):1835-42. doi:10.1016/s0140-6736(14)61687-1 |
| Ovarian Female | Intrauterine device (any) | Binary | 0.68 | Literature | 0.14 | 0.14 | Wheeler LJ, Desanto K, Teal SB, Sheeder J, Guntupalli SR. Intrauterine Device Use and Ovarian Cancer Risk: A Systematic Review and Meta-analysis. Obstet Gynecol. Oct 2019;134(4):791-800. doi:10.1097/aog.0000000000003463 |
| Ovarian Female | Number of live births | Continuous | 0.89 | UK Biobank | 1.00 |  | Tsilidis KK, Allen NE, Key TJ, et al. Oral contraceptive use and reproductive factors and risk of ovarian cancer in the European Prospective Investigation into Cancer and Nutrition. Br J Cancer. Oct 25 2011;105(9):1436-42. doi:10.1038/bjc.2011.371 |
| Ovarian Female | Occupational asbestos exposure | Binary | 1.77 | Literature | 0.00 | 0.00 | Camargo MC, Stayner LT, Straif K, Reina M, Al-Alem U, Demers PA, Landrigan PJ. Occupational exposure to asbestos and ovarian cancer: a meta-analysis. Environ Health Perspect. Sep 2011;119(9):1211-7. doi:10.1289/ehp.1003283 |
| Ovarian Female | Oopherectomy | Binary | 0.51 | UK Biobank | 0.07 |  | Finch A, Beiner M, Lubinski J, et al. Salpingo-oophorectomy and the Risk of Ovarian, Fallopian Tube, and Peritoneal Cancers in Women With a BRCA1 or BRCA2 Mutation. JAMA. 2006;296(2):185-192. doi:10.1001/jama.296.2.185 |
| Ovarian Female | Oral contraceptive use | Binary | 0.76 | UK Biobank | 0.81 |  | Havrilesky LJ, Moorman PG, Lowery WJ, et al. Oral contraceptive pills as primary prevention for ovarian cancer: a systematic review and meta-analysis. Obstet Gynecol. Jul 2013;122(1):139-147. doi:10.1097/AOG.0b013e318291c235 |
| Ovarian Female | Pack-years smoking history (fifth-root transformation) | Continuous | 1.04 | UK Biobank | 0.00 |  | Jordan SJ, Whiteman DC, Purdie DM, Green AC, Webb PM. Does smoking increase risk of ovarian cancer? A systematic review. Gynecol Oncol. Dec 2006;103(3):1122-9. doi:10.1016/j.ygyno.2006.08.012 |
| Ovarian Female | Pelvic inflammatory disease | Binary | 1.92 | Literature | 0.04 | 0.04 | Lin HW, Tu YY, Lin SY, et al. Risk of ovarian cancer in women with pelvic inflammatory disease: a population-based study. Lancet Oncol. Sep 2011;12(9):900-4. doi:10.1016/s1470-2045(11)70165-6 |
| Ovarian Female | Tubal ligation | Binary | 0.70 | Literature | 0.22 | 0.22 | Rice MS, Murphy MA, Tworoger SS. Tubal ligation, hysterectomy and ovarian cancer: A meta-analysis. J Ovarian Res. May 15 2012;5(1):13. doi:10.1186/1757-2215-5-13 |
| Pancreatic Female | Alcohol consumption (>=2 drinks/day) | Binary | 1.22 | Literature | 0.17 | 0.17 | Genkinger JM, Spiegelman D, Anderson KE, et al. Alcohol intake and pancreatic cancer risk: a pooled analysis of fourteen cohort studies. Cancer Epidemiol Biomarkers Prev. Mar 2009;18(3):765-76. doi:10.1158/1055-9965.Epi-08-0880 |
| Pancreatic Female | Body mass index | Continuous | 1.02 | UK Biobank | 29.80 |  | Michaud DS, Giovannucci E, Willett WC, Colditz GA, Stampfer MJ, Fuchs CS. Physical activity, obesity, height, and the risk of pancreatic cancer. Jama. Aug 22-29 2001;286(8):921-9. doi:10.1001/jama.286.8.921 |
| Pancreatic Female | Chronic pancreatitis | Binary | 14.88 | UK Biobank | 0.00 |  | Lowenfels AB, Maisonneuve P, Cavallini G, et al. Pancreatitis and the risk of pancreatic cancer. International Pancreatitis Study Group. N Engl J Med. May 20 1993;328(20):1433-7. doi:10.1056/nejm199305203282001 |
| Pancreatic Female | Cystic fibrosis | Binary | 6.18 | Literature | 0.01 | 0.01 | Yamada A, Komaki Y, Komaki F, Micic D, Zullow S, Sakuraba A. Risk of gastrointestinal cancers in patients with cystic fibrosis: a systematic review and meta-analysis. Lancet Oncol. Jun 2018;19(6):758-767. doi:10.1016/s1470-2045(18)30188-8 |
| Pancreatic Female | Diabetes mellitus type 2 | Binary | 1.28 | UK Biobank | 0.04 |  | Batabyal P, Vander Hoorn S, Christophi C, Nikfarjam M. Association of diabetes mellitus and pancreatic adenocarcinoma: a meta-analysis of 88 studies. Ann Surg Oncol. Jul 2014;21(7):2453-62. doi:10.1245/s10434-014-3625-6 |
| Pancreatic Female | Family history of pancreatic cancer in first degree relative | Binary | 4.60 | Literature | 0.01 | 0.01 | Olson SH, Kurtz RC. Epidemiology of pancreatic cancer and the role of family history. J Surg Oncol. Jan 2013;107(1):1-7. doi:10.1002/jso.23149 |
| Pancreatic Female | Height (in cm) | Continuous | 1.01 | UK Biobank | 162.36 |  | Michaud DS, Giovannucci E, Willett WC, Colditz GA, Stampfer MJ, Fuchs CS. Physical activity, obesity, height, and the risk of pancreatic cancer. Jama. Aug 22-29 2001;286(8):921-9. doi:10.1001/jama.286.8.921 |
| Pancreatic Female | Minutes of moderate activity daily (<30 minutes, 30-59 minutes, 60-89 minutes, >90 minutes) | Ordinal | 0.99 | UK Biobank | 1.00 |  | Michaud DS, Giovannucci E, Willett WC, Colditz GA, Stampfer MJ, Fuchs CS. Physical activity, obesity, height, and the risk of pancreatic cancer. Jama. Aug 22-29 2001;286(8):921-9. doi:10.1001/jama.286.8.921 |
| Pancreatic Female | Pack-years smoking history (fifth-root transformation) | Continuous | 1.09 | UK Biobank | 0.00 |  | Maisonneuve P, Lowenfels AB. Risk factors for pancreatic cancer: a summary review of meta-analytical studies. Int J Epidemiol. Feb 2015;44(1):186-98. doi:10.1093/ije/dyu240 |
| Pancreatic Female | Servings of red meat (beef, lamb, or pork) per week (square-root transformation) | Binary | 1.13 | UK Biobank | 2.12 |  | Nöthlings U, Wilkens LR, Murphy SP, Hankin JH, Henderson BE, Kolonel LN. Meat and fat intake as risk factors for pancreatic cancer: the multiethnic cohort study. J Natl Cancer Inst. Oct 5 2005;97(19):1458-65. doi:10.1093/jnci/dji292 Larsson SC, Wolk A. Red and processed meat consumption and risk of pancreatic cancer: meta-analysis of prospective studies. Br J Cancer. Jan 31 2012;106(3):603-7. doi:10.1038/bjc.2011.585 |
| Pancreatic Female | Servings of vegetables per day (square-root transformation) | Continuous | 0.95 | UK Biobank | 1.22 |  | Howe GR, Jain M, Miller AB. Dietary factors and risk of pancreatic cancer: results of a Canadian population-based case-control study. Int J Cancer. Apr 15 1990;45(4):604-8. doi:10.1002/ijc.2910450405 |
| Pancreatic Female | Smoking (current) | Binary | 1.81 | UK Biobank | 0.09 |  | Maisonneuve P, Lowenfels AB. Risk factors for pancreatic cancer: a summary review of meta-analytical studies. Int J Epidemiol. Feb 2015;44(1):186-98. doi:10.1093/ije/dyu240 |
| Pancreatic Male | Alcohol consumption (>=2 drinks/day) | Binary | 1.22 | Literature | 0.17 | 0.17 | Genkinger JM, Spiegelman D, Anderson KE, et al. Alcohol intake and pancreatic cancer risk: a pooled analysis of fourteen cohort studies. Cancer Epidemiol Biomarkers Prev. Mar 2009;18(3):765-76. doi:10.1158/1055-9965.Epi-08-0880 |
| Pancreatic Male | Body mass index | Continuous | 1.03 | UK Biobank | 29.40 |  | Michaud DS, Giovannucci E, Willett WC, Colditz GA, Stampfer MJ, Fuchs CS. Physical activity, obesity, height, and the risk of pancreatic cancer. Jama. Aug 22-29 2001;286(8):921-9. doi:10.1001/jama.286.8.921 |
| Pancreatic Male | Chronic pancreatitis | Binary | 12.88 | UK Biobank | 0.00 |  | Lowenfels AB, Maisonneuve P, Cavallini G, et al. Pancreatitis and the risk of pancreatic cancer. International Pancreatitis Study Group. N Engl J Med. May 20 1993;328(20):1433-7. doi:10.1056/nejm199305203282001 |
| Pancreatic Male | Cystic fibrosis | Binary | 6.18 | Literature | 0.01 | 0.01 | Yamada A, Komaki Y, Komaki F, Micic D, Zullow S, Sakuraba A. Risk of gastrointestinal cancers in patients with cystic fibrosis: a systematic review and meta-analysis. Lancet Oncol. Jun 2018;19(6):758-767. doi:10.1016/s1470-2045(18)30188-8 |
| Pancreatic Male | Diabetes mellitus type 2 | Binary | 1.13 | UK Biobank | 0.08 |  | Batabyal P, Vander Hoorn S, Christophi C, Nikfarjam M. Association of diabetes mellitus and pancreatic adenocarcinoma: a meta-analysis of 88 studies. Ann Surg Oncol. Jul 2014;21(7):2453-62. doi:10.1245/s10434-014-3625-6 |
| Pancreatic Male | Family history of pancreatic cancer in first degree relative | Binary | 4.60 | Literature | 0.01 | 0.01 | Olson SH, Kurtz RC. Epidemiology of pancreatic cancer and the role of family history. J Surg Oncol. Jan 2013;107(1):1-7. doi:10.1002/jso.23149 |
| Pancreatic Male | Height (in cm) | Continuous | 1.02 | UK Biobank | 175.48 |  | Michaud DS, Giovannucci E, Willett WC, Colditz GA, Stampfer MJ, Fuchs CS. Physical activity, obesity, height, and the risk of pancreatic cancer. Jama. Aug 22-29 2001;286(8):921-9. doi:10.1001/jama.286.8.921 |
| Pancreatic Male | Minutes of vigorous activity daily (<30 minutes, 30-59 minutes, 60-89 minutes, >90 minutes) | Ordinal | 0.97 | UK Biobank | 1.00 |  | Michaud DS, Giovannucci E, Willett WC, Colditz GA, Stampfer MJ, Fuchs CS. Physical activity, obesity, height, and the risk of pancreatic cancer. Jama. Aug 22-29 2001;286(8):921-9. doi:10.1001/jama.286.8.921 |
| Pancreatic Male | Servings of fruit per day (square-root transformation | Continuous | 0.94 | UK Biobank | 1.00 |  | Maisonneuve P, Lowenfels AB. Risk factors for pancreatic cancer: a summary review of meta-analytical studies. Int J Epidemiol. Feb 2015;44(1):186-98. doi:10.1093/ije/dyu240 |
| Pancreatic Male | Servings of processed meat (bacon, ham, sausages, burgers, nuggets, etc.) per week (square-root transformation) | Continuous | 1.05 | UK Biobank | 1.64 |  | Nöthlings U, Wilkens LR, Murphy SP, Hankin JH, Henderson BE, Kolonel LN. Meat and fat intake as risk factors for pancreatic cancer: the multiethnic cohort study. J Natl Cancer Inst. Oct 5 2005;97(19):1458-65. doi:10.1093/jnci/dji292 Larsson SC, Wolk A. Red and processed meat consumption and risk of pancreatic cancer: meta-analysis of prospective studies. Br J Cancer. Jan 31 2012;106(3):603-7. doi:10.1038/bjc.2011.585 |
| Pancreatic Male | Servings of red meat (beef, lamb, or pork) per week (square-root transformation) | Binary | 1.06 | UK Biobank | 2.12 |  | Nöthlings U, Wilkens LR, Murphy SP, Hankin JH, Henderson BE, Kolonel LN. Meat and fat intake as risk factors for pancreatic cancer: the multiethnic cohort study. J Natl Cancer Inst. Oct 5 2005;97(19):1458-65. doi:10.1093/jnci/dji292 Larsson SC, Wolk A. Red and processed meat consumption and risk of pancreatic cancer: meta-analysis of prospective studies. Br J Cancer. Jan 31 2012;106(3):603-7. doi:10.1038/bjc.2011.585 |
| Pancreatic Male | Smoking (current) | Binary | 1.48 | UK Biobank | 0.13 |  | Maisonneuve P, Lowenfels AB. Risk factors for pancreatic cancer: a summary review of meta-analytical studies. Int J Epidemiol. Feb 2015;44(1):186-98. doi:10.1093/ije/dyu240 |
| Prostate Male | >=1 serving of soy daily | Binary | 0.70 | Literature | 0.00 | 0.00 | Yan L, Spitznagel EL. Meta-analysis of soy food and risk of prostate cancer in men. Int J Cancer. Nov 20 2005;117(4):667-9. doi:10.1002/ijc.21266 |
| Prostate Male | Alcohol consumption (2-3 drinks/day) | Binary | 1.05 | Literature | 0.12 | 0.12 | Bagnardi V, Blangiardo M, La Vecchia C, Corrao G. Alcohol consumption and the risk of cancer: a meta-analysis. Alcohol Res Health. 2001;25(4):263-70. https://www.ncbi.nlm.nih.gov/pmc/articles/PMC6705703/ |
| Prostate Male | Alcohol consumption (4+ drinks/day) | Binary | 1.09 | Literature | 0.05 | 0.05 | Bagnardi V, Blangiardo M, La Vecchia C, Corrao G. Alcohol consumption and the risk of cancer: a meta-analysis. Alcohol Res Health. 2001;25(4):263-70. https://www.ncbi.nlm.nih.gov/pmc/articles/PMC6705703/ |
| Prostate Male | Family history of breast cancer in first degree relative | Binary | 1.21 | Literature | 0.11 | 0.11 | Barber L, Gerke T, Markt SC, et al. Family History of Breast or Prostate Cancer and Prostate Cancer Risk. Clin Cancer Res. Dec 1 2018;24(23):5910-5917. doi:10.1158/1078-0432.Ccr-18-0370 |
| Prostate Male | Family history of prostate cancer in first degree relative | Binary | 1.68 | Literature | 0.14 | 0.10 | Barber L, Gerke T, Markt SC, et al. Family History of Breast or Prostate Cancer and Prostate Cancer Risk. Clin Cancer Res. Dec 1 2018;24(23):5910-5917. doi:10.1158/1078-0432.Ccr-18-0370 |
| Prostate Male | Minutes of moderate activity daily (<30 minutes, 30-59 minutes, 60-89 minutes, >90 minutes) | Ordinal | 1.00 | UK Biobank | 1.00 |  | Giovannucci EL, Liu Y, Leitzmann MF, Stampfer MJ, Willett WC. A prospective study of physical activity and incident and fatal prostate cancer. Arch Intern Med. May 9 2005;165(9):1005-10. doi:10.1001/archinte.165.9.1005 |
| Prostate Male | Servings of processed meat (bacon, ham, sausages, burgers, nuggets, etc.) per week (square-root transformation) | Continuous | 1.00 | UK Biobank | 1.64 |  | Sinha R, Park Y, Graubard BI, Leitzmann MF, Hollenbeck A, Schatzkin A, Cross AJ. Meat and meat-related compounds and risk of prostate cancer in a large prospective cohort study in the United States. Am J Epidemiol. Nov 1 2009;170(9):1165-77. doi:10.1093/aje/kwp280 |
| Prostate Male | Servings of red meat (beef, lamb, or pork) per week (square-root transformation) | Binary | 1.09 | UK Biobank | 2.12 |  | Sinha R, Park Y, Graubard BI, Leitzmann MF, Hollenbeck A, Schatzkin A, Cross AJ. Meat and meat-related compounds and risk of prostate cancer in a large prospective cohort study in the United States. Am J Epidemiol. Nov 1 2009;170(9):1165-77. doi:10.1093/aje/kwp280 |
| Prostate Male | Statin use | Binary | 0.84 | UK Biobank | 0.23 |  | Cao Z, Yao J, He Y, et al. Association Between Statin Exposure and Incidence and Prognosis of Prostate Cancer: A Meta-analysis Based on Observational Studies. Am J Clin Oncol. Jul 1 2023;46(7):323-334. doi:10.1097/coc.0000000000001012 |
| Stomach Female | Alcohol consumption (2-3 drinks/day) | Binary | 1.07 | Literature | 0.12 | 0.12 | Bagnardi V, Blangiardo M, La Vecchia C, Corrao G. Alcohol consumption and the risk of cancer: a meta-analysis. Alcohol Res Health. 2001;25(4):263-70. https://www.ncbi.nlm.nih.gov/pmc/articles/PMC6705703/ |
| Stomach Female | Alcohol consumption (4+ drinks/day) | Binary | 1.15 | Literature | 0.05 | 0.05 | Bagnardi V, Blangiardo M, La Vecchia C, Corrao G. Alcohol consumption and the risk of cancer: a meta-analysis. Alcohol Res Health. 2001;25(4):263-70. https://www.ncbi.nlm.nih.gov/pmc/articles/PMC6705703/ |
| Stomach Female | Body mass index | Continuous | 1.04 | UK Biobank | 29.80 |  | Yang P, Zhou Y, Chen B, Wan HW, Jia GQ, Bai HL, Wu XT. Overweight, obesity and gastric cancer risk: results from a meta-analysis of cohort studies. Eur J Cancer. Nov 2009;45(16):2867-73. doi:10.1016/j.ejca.2009.04.019 |
| Stomach Female | Ever add salt to food (never/rarely, sometimes, usually, always) | Ordinal | 1.08 | UK Biobank | 1.00 |  | Tsugane S, Sasazuki S. Diet and the risk of gastric cancer: review of epidemiological evidence. Gastric Cancer. 2007;10(2):75-83. doi:10.1007/s10120-007-0420-0 Kronsteiner-Gicevic S, Thompson AS, Gaggl M, Bell W, Cassidy A, Kühn T. Adding salt to food at table as an indicator of gastric cancer risk among adults: a prospective study. Gastric Cancer. Jul 2024;27(4):714-721. doi:10.1007/s10120-024-01502-9 |
| Stomach Female | H. Pylori infection | Binary | 2.04 | Literature | 0.30 | 0.30 | Eslick GD, Lim LL-Y, Byles JE, Xia HH-X, Talley NJ. Association ofHelicobacter PyloriInfection With Gastric Carcinoma: A Meta-Analysis. Official journal of the American College of Gastroenterology \| ACG. 1999;94(9):2373-2379. doi:10.1111/j.1572-0241.1999.01360.x |
| Stomach Female | Hiatal hernia | Binary | 2.26 | Literature | 0.07 | 0.07 | Wu AH, Tseng C-C, Bernstein L. Hiatal hernia, reflux symptoms, body size, and risk of esophageal and gastric adenocarcinoma. Cancer. 2003;98(5):940-948. doi:https://doi.org/10.1002/cncr.11568 |
| Stomach Female | Nonsteroidal anti-inflammatory drug use | Binary | 0.60 | UK Biobank | 0.17 |  | Wu CY, Wu MS, Kuo KN, Wang CB, Chen YJ, Lin JT. Effective reduction of gastric cancer risk with regular use of nonsteroidal anti-inflammatory drugs in Helicobacter pylori-infected patients. J Clin Oncol. Jun 20 2010;28(18):2952-7. doi:10.1200/jco.2009.26.0695 |
| Stomach Female | Pack-years smoking history (fifth-root transformation) | Continuous | 1.16 | UK Biobank | 0.00 |  | Ladeiras-Lopes R, Pereira AK, Nogueira A, Pinheiro-Torres T, Pinto I, Santos-Pereira R, Lunet N. Smoking and gastric cancer: systematic review and meta-analysis of cohort studies. Cancer Causes Control. Sep 2008;19(7):689-701. doi:10.1007/s10552-008-9132-y |
| Stomach Female | Servings of fatty fish per week (square-root transformation) | Continuous | 0.96 | UK Biobank | 1.19 |  | Tsugane S, Sasazuki S. Diet and the risk of gastric cancer: review of epidemiological evidence. Gastric Cancer. 2007;10(2):75-83. doi:10.1007/s10120-007-0420-0 |
| Stomach Female | Servings of fruit per day (square-root transformation | Continuous | 0.99 | UK Biobank | 1.00 |  | Tsugane S, Sasazuki S. Diet and the risk of gastric cancer: review of epidemiological evidence. Gastric Cancer. 2007;10(2):75-83. doi:10.1007/s10120-007-0420-0 |
| Stomach Female | Servings of vegetables per day (square-root transformation) | Continuous | 1.00 | UK Biobank | 1.22 |  | Tsugane S, Sasazuki S. Diet and the risk of gastric cancer: review of epidemiological evidence. Gastric Cancer. 2007;10(2):75-83. doi:10.1007/s10120-007-0420-0 |
| Stomach Female | Smoking (current) | Binary | 1.09 | UK Biobank | 0.09 |  | Ladeiras-Lopes R, Pereira AK, Nogueira A, Pinheiro-Torres T, Pinto I, Santos-Pereira R, Lunet N. Smoking and gastric cancer: systematic review and meta-analysis of cohort studies. Cancer Causes Control. Sep 2008;19(7):689-701. doi:10.1007/s10552-008-9132-y |
| Stomach Male | Alcohol consumption (2-3 drinks/day) | Binary | 1.07 | Literature | 0.12 | 0.12 | Bagnardi V, Blangiardo M, La Vecchia C, Corrao G. Alcohol consumption and the risk of cancer: a meta-analysis. Alcohol Res Health. 2001;25(4):263-70. https://www.ncbi.nlm.nih.gov/pmc/articles/PMC6705703/ |
| Stomach Male | Alcohol consumption (4+ drinks/day) | Binary | 1.15 | Literature | 0.05 | 0.05 | Bagnardi V, Blangiardo M, La Vecchia C, Corrao G. Alcohol consumption and the risk of cancer: a meta-analysis. Alcohol Res Health. 2001;25(4):263-70. https://www.ncbi.nlm.nih.gov/pmc/articles/PMC6705703/ |
| Stomach Male | Body mass index | Continuous | 1.05 | UK Biobank | 29.40 |  | Yang P, Zhou Y, Chen B, Wan HW, Jia GQ, Bai HL, Wu XT. Overweight, obesity and gastric cancer risk: results from a meta-analysis of cohort studies. Eur J Cancer. Nov 2009;45(16):2867-73. doi:10.1016/j.ejca.2009.04.019 |
| Stomach Male | Ever add salt to food (never/rarely, sometimes, usually, always) | Ordinal | 1.02 | UK Biobank | 1.00 |  | Tsugane S, Sasazuki S. Diet and the risk of gastric cancer: review of epidemiological evidence. Gastric Cancer. 2007;10(2):75-83. doi:10.1007/s10120-007-0420-0 Kronsteiner-Gicevic S, Thompson AS, Gaggl M, Bell W, Cassidy A, Kühn T. Adding salt to food at table as an indicator of gastric cancer risk among adults: a prospective study. Gastric Cancer. Jul 2024;27(4):714-721. doi:10.1007/s10120-024-01502-9 |
| Stomach Male | H. Pylori infection | Binary | 2.04 | Literature | 0.30 | 0.30 | Eslick GD, Lim LL-Y, Byles JE, Xia HH-X, Talley NJ. Association ofHelicobacter PyloriInfection With Gastric Carcinoma: A Meta-Analysis. Official journal of the American College of Gastroenterology \| ACG. 1999;94(9):2373-2379. doi:10.1111/j.1572-0241.1999.01360.x |
| Stomach Male | Hiatal hernia | Binary | 2.26 | Literature | 0.07 | 0.07 | Wu AH, Tseng C-C, Bernstein L. Hiatal hernia, reflux symptoms, body size, and risk of esophageal and gastric adenocarcinoma. Cancer. 2003;98(5):940-948. doi:https://doi.org/10.1002/cncr.11568 |
| Stomach Male | Nonsteroidal anti-inflammatory drug use | Binary | 0.73 | UK Biobank | 0.12 |  | Wu CY, Wu MS, Kuo KN, Wang CB, Chen YJ, Lin JT. Effective reduction of gastric cancer risk with regular use of nonsteroidal anti-inflammatory drugs in Helicobacter pylori-infected patients. J Clin Oncol. Jun 20 2010;28(18):2952-7. doi:10.1200/jco.2009.26.0695 |
| Stomach Male | Pack-years smoking history (fifth-root transformation) | Continuous | 1.26 | UK Biobank | 0.00 |  | Ladeiras-Lopes R, Pereira AK, Nogueira A, Pinheiro-Torres T, Pinto I, Santos-Pereira R, Lunet N. Smoking and gastric cancer: systematic review and meta-analysis of cohort studies. Cancer Causes Control. Sep 2008;19(7):689-701. doi:10.1007/s10552-008-9132-y |
| Stomach Male | Servings of fatty fish per week (square-root transformation) | Continuous | 0.85 | UK Biobank | 1.16 |  | Tsugane S, Sasazuki S. Diet and the risk of gastric cancer: review of epidemiological evidence. Gastric Cancer. 2007;10(2):75-83. doi:10.1007/s10120-007-0420-0 |
| Stomach Male | Servings of processed meat (bacon, ham, sausages, burgers, nuggets, etc.) per week (square-root transformation) | Continuous | 1.02 | UK Biobank | 1.64 |  | Tsugane S, Sasazuki S. Diet and the risk of gastric cancer: review of epidemiological evidence. Gastric Cancer. 2007;10(2):75-83. doi:10.1007/s10120-007-0420-0 |
| Stomach Male | Servings of vegetables per day (square-root transformation) | Continuous | 0.90 | UK Biobank | 1.22 |  | Tsugane S, Sasazuki S. Diet and the risk of gastric cancer: review of epidemiological evidence. Gastric Cancer. 2007;10(2):75-83. doi:10.1007/s10120-007-0420-0 |
| Stomach Male | Smoking (current) | Binary | 1.27 | UK Biobank | 0.13 |  | Ladeiras-Lopes R, Pereira AK, Nogueira A, Pinheiro-Torres T, Pinto I, Santos-Pereira R, Lunet N. Smoking and gastric cancer: systematic review and meta-analysis of cohort studies. Cancer Causes Control. Sep 2008;19(7):689-701. doi:10.1007/s10552-008-9132-y |
| Stomach Male | Statin use | Binary | 0.92 | UK Biobank | 0.23 |  | Su CH, Islam MM, Jia G, Wu CC. Statins and the Risk of Gastric Cancer: A Systematic Review and Meta-Analysis. J Clin Med. Dec 2 2022;11(23)doi:10.3390/jcm11237180 |
| Testicular Male | Cannabis use >50 times in lifetime | Binary | 2.57 | Literature | 0.17 | 0.17 | Callaghan RC, Allebeck P, Akre O, McGlynn KA, Sidorchuk A. Cannabis Use and Incidence of Testicular Cancer: A 42-Year Follow-up of Swedish Men between 1970 and 2011. Cancer Epidemiol Biomarkers Prev. Nov 2017;26(11):1644-1652. doi:10.1158/1055-9965.Epi-17-0428 |
| Testicular Male | Cannabis use at least weekly | Binary | 1.92 | Literature | 0.05 | 0.05 | Gurney J, Shaw C, Stanley J, Signal V, Sarfati D. Cannabis exposure and risk of testicular cancer: a systematic review and meta-analysis. BMC Cancer. Nov 11 2015;15:897. doi:10.1186/s12885-015-1905-6 |
| Testicular Male | Cannabis use currently | Binary | 1.62 | Literature | 0.21 | 0.21 | Gurney J, Shaw C, Stanley J, Signal V, Sarfati D. Cannabis exposure and risk of testicular cancer: a systematic review and meta-analysis. BMC Cancer. Nov 11 2015;15:897. doi:10.1186/s12885-015-1905-6 |
| Testicular Male | Family history of testicular cancer in first degree relative | Binary | 5.50 | Literature | 0.02 | 0.02 | Del Risco Kollerud R, Ruud E, Haugnes HS, et al. Family history of cancer and risk of paediatric and young adult's testicular cancer: A Norwegian cohort study. Br J Cancer. May 2019;120(10):1007-1014. doi:10.1038/s41416-019-0445-2 |
| Testicular Male | Infertility | Binary | 2.80 | Literature | 0.15 | 0.15 | Walsh TJ, Croughan MS, Schembri M, Chan JM, Turek PJ. Increased risk of testicular germ cell cancer among infertile men. Arch Intern Med. Feb 23 2009;169(4):351-6. doi:10.1001/archinternmed.2008.562 |
| Testicular Male | Undescended testis | Binary | 26.89 | UK Biobank | 0.00 |  | Florou M, Tsilidis KK, Siomou E, et al. Orchidopexy for congenital cryptorchidism in childhood and adolescence and testicular cancer in adults: an updated systematic review and meta-analysis of observational studies. Eur J Pediatr. Jun 2023;182(6):2499-2507. doi:10.1007/s00431-023-04947-9 |
| Thyroid Female | Body mass index | Continuous | 1.03 | UK Biobank | 29.80 |  | Kitahara CM, Pfeiffer RM, Sosa JA, Shiels MS. Impact of Overweight and Obesity on US Papillary Thyroid Cancer Incidence Trends (1995-2015). J Natl Cancer Inst. Aug 1 2020;112(8):810-817. doi:10.1093/jnci/djz202 |
| Thyroid Female | Chronic hepatitis C | Binary | 6.28 | UK Biobank | 0.00 |  | Antonelli A, Ferri C, Fallahi P, et al. Thyroid cancer in HCV-related chronic hepatitis patients: a case-control study. Thyroid. May 2007;17(5):447-51. doi:10.1089/thy.2006.0194 |
| Thyroid Female | Family history of thyroid cancer in first degree relative | Binary | 10.30 | Literature | 0.01 | 0.01 | Pal T, Vogl FD, Chappuis PO, et al. Increased risk for nonmedullary thyroid cancer in the first degree relatives of prevalent cases of nonmedullary thyroid cancer: a hospital-based study. J Clin Endocrinol Metab. Nov 2001;86(11):5307-12. doi:10.1210/jcem.86.11.8010 |
| Thyroid Female | Hysterectomy | Binary | 1.14 | Literature | 0.17 | 0.17 | Fabiani R, Rosignoli P, Giacchetta I, Chiavarini M. Hysterectomy and thyroid cancer risk: A systematic review and meta-analysis. Glob Epidemiol. Dec 2023;6:100122. doi:10.1016/j.gloepi.2023.100122 |
| Thyroid Female | Minutes of moderate activity daily (<30 minutes, 30-59 minutes, 60-89 minutes, >90 minutes) | Ordinal | 0.99 | UK Biobank | 1.00 |  | Fiore M, Cristaldi A, Okatyeva V, et al. Physical Activity and Thyroid Cancer Risk: A Case-Control Study in Catania (South Italy). Int J Environ Res Public Health. Apr 22 2019;16(8)doi:10.3390/ijerph16081428 |
| Thyroid Female | Minutes of vigorous activity daily (<30 minutes, 30-59 minutes, 60-89 minutes, >90 minutes) | Ordinal | 0.93 | UK Biobank | 1.00 |  | Fiore M, Cristaldi A, Okatyeva V, et al. Physical Activity and Thyroid Cancer Risk: A Case-Control Study in Catania (South Italy). Int J Environ Res Public Health. Apr 22 2019;16(8)doi:10.3390/ijerph16081428 |
| Thyroid Female | Number of live births | Continuous | 1.16 | UK Biobank | 1.00 |  | Rossing MA, Voigt LF, Wicklund KG, Daling JR. Reproductive factors and risk of papillary thyroid cancer in women. Am J Epidemiol. Apr 15 2000;151(8):765-72. doi:10.1093/oxfordjournals.aje.a010276 |
| Thyroid Male | Body mass index | Continuous | 1.03 | UK Biobank | 29.40 |  | Kitahara CM, Pfeiffer RM, Sosa JA, Shiels MS. Impact of Overweight and Obesity on US Papillary Thyroid Cancer Incidence Trends (1995-2015). J Natl Cancer Inst. Aug 1 2020;112(8):810-817. doi:10.1093/jnci/djz202 |
| Thyroid Male | Chronic hepatitis C | Binary | 11.75 | UK Biobank | 0.00 |  | Antonelli A, Ferri C, Fallahi P, et al. Thyroid cancer in HCV-related chronic hepatitis patients: a case-control study. Thyroid. May 2007;17(5):447-51. doi:10.1089/thy.2006.0194 |
| Thyroid Male | Family history of thyroid cancer in first degree relative | Binary | 10.30 | Literature | 0.01 | 0.01 | Pal T, Vogl FD, Chappuis PO, et al. Increased risk for nonmedullary thyroid cancer in the first degree relatives of prevalent cases of nonmedullary thyroid cancer: a hospital-based study. J Clin Endocrinol Metab. Nov 2001;86(11):5307-12. doi:10.1210/jcem.86.11.8010 |
| Thyroid Male | Minutes of moderate activity daily (<30 minutes, 30-59 minutes, 60-89 minutes, >90 minutes) | Ordinal | 0.96 | UK Biobank | 1.00 |  | Fiore M, Cristaldi A, Okatyeva V, et al. Physical Activity and Thyroid Cancer Risk: A Case-Control Study in Catania (South Italy). Int J Environ Res Public Health. Apr 22 2019;16(8)doi:10.3390/ijerph16081428 |
| Thyroid Male | Minutes of vigorous activity daily (<30 minutes, 30-59 minutes, 60-89 minutes, >90 minutes) | Ordinal | 0.88 | UK Biobank | 1.00 |  | Fiore M, Cristaldi A, Okatyeva V, et al. Physical Activity and Thyroid Cancer Risk: A Case-Control Study in Catania (South Italy). Int J Environ Res Public Health. Apr 22 2019;16(8)doi:10.3390/ijerph16081428 |
| Thyroid Male | Servings of vegetables per day (square-root transformation) | Continuous | 0.94 | UK Biobank | 1.22 |  | Jung SK, Kim K, Tae K, Kong G, Kim MK. The effect of raw vegetable and fruit intake on thyroid cancer risk among women: a case-control study in South Korea. Br J Nutr. Jan 14 2013;109(1):118-28. doi:10.1017/s0007114512000591 |

**eTable 2. Ideal values for modifiable risk factors**

| **Variable** | **Ideal value** | **Reference** |
| --- | --- | --- |
| *Lifestyle-associated risk factors* |  |  |
| Alcohol consumption (drinks/night) | 0 | U.S. Surgeon General. (2025) Alcohol and cancer risk. The U.S. Surgeon General’s Advisory. US: Office of the Surgeon General. |
| Current smoking | no | Freedman ND, Abnet CC, Caporaso NE, et al. Impact of changing US cigarette smoking patterns on incident cancer: risks of 20 smoking-related cancers among the women and men of the NIH-AARP cohort. *Int J Epidemiol*. Jun 2016;45(3):846-56. doi:10.1093/ije/dyv175 |
| Chewing tobacco use | no | Li X, Koskinen AI, Hemminki O, Försti A, Sundquist J, Sundquist K, Hemminki K. Family History of Head and Neck Cancers. *Cancers (Basel)*. Aug 16 2021;13(16)doi:10.3390/cancers13164115 |
| Snuff use | no | Wyss AB, Hashibe M, Lee YA, et al. Smokeless Tobacco Use and the Risk of Head and Neck Cancer: Pooled Analysis of US Studies in the INHANCE Consortium. *Am J Epidemiol*. Nov 15 2016;184(10):703-716. doi:10.1093/aje/kww075 |
| Current cannabis use (any) | no | Gurney J, Shaw C, Stanley J, Signal V, Sarfati D. Cannabis exposure and risk of testicular cancer: a systematic review and meta-analysis. *BMC Cancer*. Nov 11 2015;15:897. doi:10.1186/s12885-015-1905-6 |
| Use of cannabis > once a week | no | Gurney J, Shaw C, Stanley J, Signal V, Sarfati D. Cannabis exposure and risk of testicular cancer: a systematic review and meta-analysis. *BMC Cancer*. Nov 11 2015;15:897. doi:10.1186/s12885-015-1905-6 |
| *Physical activity* |  |  |
| Body mass index | <25 | US Centers for Disease Control and Prevention. “Obesity and Cancer.”. <https://www.cdc.gov/cancer/risk-factors/obesity.html>. Accessed 4/5/2025. |
| Duration of moderate activity daily | ≥30 minutes | Rock CL, Thomson C, Gansler T, et al. American Cancer Society guideline for diet and physical activity for cancer prevention. *CA Cancer J Clin*. Jul 2020;70(4):245-271. doi:10.3322/caac.21591 |
| Duration of vigorous activity daily | ≥30 minutes | Rock CL, Thomson C, Gansler T, et al. American Cancer Society guideline for diet and physical activity for cancer prevention. *CA Cancer J Clin*. Jul 2020;70(4):245-271. doi:10.3322/caac.21591 |
| *Diet* |  |  |
| Servings of fruit per day | ≥2 | Rock CL, Thomson C, Gansler T, et al. American Cancer Society guideline for diet and physical activity for cancer prevention. *CA Cancer J Clin*. Jul 2020;70(4):245-271. doi:10.3322/caac.21591 |
| Servings of vegetables per day | ≥2 | Rock CL, Thomson C, Gansler T, et al. American Cancer Society guideline for diet and physical activity for cancer prevention. *CA Cancer J Clin*. Jul 2020;70(4):245-271. doi:10.3322/caac.21591 |
| Servings of fatty fish per week | ≥3 | American Heart Association. “Fish and Omega-3 Fatty Acids.” https://www.heart.org/en/healthy-living/healthy-eating/eat-smart/fats/fish-and-omega-3-fatty-acids. Accessed 4/5/2025. |
| Servings of processed meat (bacon, ham, sausages, burgers, nuggets, etc.) per week | 0 | Rock CL, Thomson C, Gansler T, et al. American Cancer Society guideline for diet and physical activity for cancer prevention. *CA Cancer J Clin*. Jul 2020;70(4):245-271. doi:10.3322/caac.21591 |
| Servings of red meat (beef, lamb, or pork) per week | 0 | Rock CL, Thomson C, Gansler T, et al. American Cancer Society guideline for diet and physical activity for cancer prevention. *CA Cancer J Clin*. Jul 2020;70(4):245-271. doi:10.3322/caac.21591 |
| At least 1 serving of soy daily | yes | Yan L, Spitznagel EL. Meta-analysis of soy food and risk of prostate cancer in men. *Int J Cancer*. Nov 20 2005;117(4):667-9. doi:10.1002/ijc.21266 |
| Ever add salt to food | Never/rarely | Kronsteiner-Gicevic S, Thompson AS, Gaggl M, Bell W, Cassidy A, Kühn T. Adding salt to food at table as an indicator of gastric cancer risk among adults: a prospective study. *Gastric Cancer*. Jul 2024;27(4):714-721. doi:10.1007/s10120-024-01502-9 |
| Glasses of water per day | ≥9 | Standing Committee on the Scientific Evaluation of Dietary Reference Intakes Panel on Dietary Reference Intakes for Electrolytes Water. *Dietary reference intakes for water, potassium, sodium, chloride, and sulfate*. National Academies Press; 2005. |
| ≥ 1 serving daily of sugar-sweetened beverages | no | Rock CL, Thomson C, Gansler T, et al. American Cancer Society guideline for diet and physical activity for cancer prevention. *CA Cancer J Clin*. Jul 2020;70(4):245-271. doi:10.3322/caac.21591 |
| Cups of tea daily | ≥2 | Song Y, Wang Z, Jin Y, Guo J. Association between tea and coffee consumption and brain cancer risk: an updated meta-analysis. *World J Surg Oncol*. Mar 15 2019;17(1):51. doi:10.1186/s12957-019-1591-y |
| Cups of coffee daily | ≥2 | Song Y, Wang Z, Jin Y, Guo J. Association between tea and coffee consumption and brain cancer risk: an updated meta-analysis. *World J Surg Oncol*. Mar 15 2019;17(1):51. doi:10.1186/s12957-019-1591-y |
| Preference to drink very hot beverages (e.g. when >140 degrees F) | no | Islami F, Poustchi H, Pourshams A, et al. A prospective study of tea drinking temperature and risk of esophageal squamous cell carcinoma. *Int J Cancer*. Jan 1 2020;146(1):18-25. doi:10.1002/ijc.32220 |
| *Oral health* |  |  |
| Brushing teeth daily | yes | Hashim D, Sartori S, Brennan P, et al. The role of oral hygiene in head and neck cancer: results from International Head and Neck Cancer Epidemiology (INHANCE) consortium. *Ann Oncol*. Aug 2016;27(8):1619-25. doi:10.1093/annonc/mdw224 |
| Gum disease | no | Hashim D, Sartori S, Brennan P, et al. The role of oral hygiene in head and neck cancer: results from International Head and Neck Cancer Epidemiology (INHANCE) consortium. *Ann Oncol*. Aug 2016;27(8):1619-25. doi:10.1093/annonc/mdw224 |
| Annual visit to dentist | yes | Hashim D, Sartori S, Brennan P, et al. The role of oral hygiene in head and neck cancer: results from International Head and Neck Cancer Epidemiology (INHANCE) consortium. *Ann Oncol*. Aug 2016;27(8):1619-25. doi:10.1093/annonc/mdw224 |
| Alcohol-based mouthwash use > once a day | no | Hashim D, Sartori S, Brennan P, et al. The role of oral hygiene in head and neck cancer: results from International Head and Neck Cancer Epidemiology (INHANCE) consortium. *Ann Oncol*. Aug 2016;27(8):1619-25. doi:10.1093/annonc/mdw224 |
| *Medications/supplements* |  |  |
| Consumption of high-dose (20-30mg/day) beta carotene supplements | no | World Cancer Research Fund/American Institute for Cancer Research. Continuous Update Project Expert Report 2018. Diet, nutrition, physical activity, and lung cancer. 2017. <https://www.wcrf.org/wp-content/uploads/2021/02/lung-cancer-report.pdf> |
| Calcium supplements (1200mg-2000mg daily) | yes | Shaukat A, Scouras N, Schünemann HJ. Role of supplemental calcium in the recurrence of colorectal adenomas: a metaanalysis of randomized controlled trials. *Am J Gastroenterol*. Feb 2005;100(2):390-4. doi:10.1111/j.1572-0241.2005.41220.x |
| Hypertension treated (if present) | yes | Christakoudi S, Kakourou A, Markozannes G, et al. Blood pressure and risk of cancer in the European Prospective Investigation into Cancer and Nutrition. *Int J Cancer*. May 15 2020;146(10):2680-2693. doi:10.1002/ijc.32576 |
| *Other exposures* |  |  |
| High levels of radon in home | no | Darby S, Hill D, Auvinen A, et al. Radon in homes and risk of lung cancer: collaborative analysis of individual data from 13 European case-control studies. *Bmj*. Jan 29 2005;330(7485):223. doi:10.1136/bmj.38308.477650.63 |
| Daily sunscreen use | yes | Green AC, Williams GM, Logan V, Strutton GM. Reduced melanoma after regular sunscreen use: randomized trial follow-up. *J Clin Oncol*. Jan 20 2011;29(3):257-63. doi:10.1200/jco.2010.28.7078 |

**eTable 3.** Percentiles of lifetime all-cancer risk in the UK Biobank cohort based on reported risk factors and with all modifiable risk factors set to ideal values

|  | **Men reported** | **Men ideal** | **Female reported** | **Female ideal** |
| --- | --- | --- | --- | --- |
| n | 211,861 | 211,861 | 234,934 | 234,934 |
| Mean | 31.72% | 21.20% | 23.42% | 17.90% |
| Standard deviation | 8.59% | 4.58% | 8.72% | 6.12% |
| Median | 29.47% | 20.46% | 20.99% | 16.51% |
| Interquartile range | 8.39% | 3.85% | 8.78% | 4.93% |
| 25th percentile | 26.33% | 18.78% | 17.65% | 14.27% |
| 75th percentile | 34.72% | 22.63% | 26.43% | 19.20% |
| Percentiles |  |  |  |  |
| 10th Percentile | 23.78% | 17.08% | 15.28% | 12.53% |
| 20th Percentile | 25.63% | 18.33% | 16.94% | 13.76% |
| 30th Percentile | 26.96% | 19.17% | 18.31% | 14.74% |
| 40th Percentile | 28.17% | 19.84% | 19.61% | 15.63% |
| 60th Percentile | 31.12% | 21.14% | 22.62% | 17.43% |
| 70th Percentile | 33.30% | 22.04% | 24.85% | 18.53% |
| 80th Percentile | 36.56% | 23.31% | 28.60% | 20.05% |
| 90th Percentile | 42.61% | 25.31% | 35.46% | 24.39% |

**eTable 4.** Percentiles of 10-year all-cancer risk in the UK Biobank cohort by decade

| **Sex** | **Male** | | | **Female** | | |
| --- | --- | --- | --- | --- | --- | --- |
| **Age decade** | **40-<50** | **50-<60** | **60-70** | **40-<50** | **50-<60** | **60-70** |
| Median | 2.55% | 7.47% | 13.88% | 3.95% | 6.27% | 8.88% |
| Interquartile range | 1.54% | 3.68% | 5.02% | 1.78% | 2.78% | 3.75% |
| 25th percentile | 1.88% | 5.73% | 11.80% | 3.23% | 5.21% | 7.55% |
| 75th percentile | 3.42% | 9.41% | 16.82% | 5.01% | 7.99% | 11.30% |
| Percentiles |  |  |  |  |  |  |
| 10th Percentile | 1.51% | 4.61% | 10.33% | 2.73% | 4.49% | 6.67% |
| 20th Percentile | 1.76% | 5.38% | 11.35% | 3.08% | 4.99% | 7.29% |
| 30th Percentile | 2.00% | 6.08% | 12.22% | 3.37% | 5.42% | 7.80% |
| 40th Percentile | 2.26% | 6.76% | 13.04% | 3.66% | 5.83% | 8.32% |
| 60th Percentile | 2.88% | 8.18% | 14.82% | 4.28% | 6.79% | 9.56% |
| 70th Percentile | 3.24% | 8.93% | 16.05% | 4.72% | 7.49% | 10.55% |
| 80th Percentile | 3.64% | 9.98% | 17.81% | 5.41% | 8.68% | 12.39% |
| 90th Percentile | 4.35% | 11.91% | 21.13% | 7.41% | 11.54% | 16.09% |
